# GRAd-COV2, a gorilla adenovirus based candidate vaccine against COVID-19, is safe and immunogenic in young and older adults

**DOI:** 10.1101/2021.04.10.21255202

**Authors:** Simone Lanini, Stefania Capone, Andrea Antinori, Stefano Milleri, Emanuele Nicastri, Roberto Camerini, Chiara Agrati, Concetta Castilletti, Federica Mori, Alessandra Sacchi, Giulia Matusali, Roberta Gagliardini, Virginia Ammendola, Eleonora Cimini, Fabiana Grazioli, Laura Scorzolini, Federico Napolitano, Maria Maddalena Piazzi, Marco Soriani, Aldo De Luca, Simone Battella, Andrea Sommella, Alessandra M. Contino, Federica Barra, Michela Gentile, Angelo Raggioli, Yufang Shi, Enrico Girardi, Markus Maeurer, Maria R. Capobianchi, Francesco Vaia, Mauro Piacentini, Guido Kroemer, Alessandra Vitelli, Stefano Colloca, Antonella Folgori, Giuseppe Ippolito

**Affiliations:** Istituto Nazionale per Le Malattie Infettive Lazzaro Spallanzani IRCCS; Rome, Italy; ReiThera Srl; Rome, Italy; Centro Ricerche Cliniche di Verona srl; Verona, Italy; First Affiliated Hospital of Soochow University; Suzhou, Jiangsu, China; Shanghai Institute of Nutrition and Health, Shanghai Institutes for Biological Sciences, Chinese Academy of Sciences; Shanghai, China; Division of Immunotherapy, ImmunoSurgery, Champalimaud Foundation; Lisboa, Portugal; I Medical Clinic, University of Mainz; Mainz, Germany; Department of Biology, University of Rome “Tor Vergata; Rome, Italy; Sorbonne Université, Institut Universitaire de France; Paris, France; Metabolomics and Cell Biology Platforms, Institut Gustave Roussy; Villejuif, France; Pôle de Biologie, Hôpital Européen Georges Pompidou; Paris, France; Department of Women’s and Children’s Health, Karolinska University Hospital, Stockholm, Sweden

## Abstract

Safe and effective vaccines against coronavirus disease 2019 (COVID-19) are urgently needed to control the ongoing pandemic. Although impressive progress has been made with several COVID-19 vaccines already approved, it is clear that those developed so far cannot meet the global vaccine demand. We have developed a COVID-19 vaccine based on a replication-defective gorilla adenovirus expressing the stabilized pre-fusion SARS-CoV-2 Spike protein, named GRAd-COV2. We aimed to assess the safety and immunogenicity of a single-dose regimen of this vaccine in healthy younger and older adults to select the appropriate dose for each age group. To this purpose, a phase 1, dose-escalation, open-label trial was conducted including 90 healthy subjects, (45 aged 18-55 years and 45 aged 65-85 years), who received a single intramuscular administration of GRAd-CoV2 at three escalating doses. Local and systemic adverse reactions were mostly mild or moderate and of short duration, and no serious AE was reported. Four weeks after vaccination, seroconversion to Spike/RBD was achieved in 43/44 young volunteers and in 45/45 older subjects. Consistently, neutralizing antibodies were detected in 42/44 younger age and 45/45 older age volunteers. In addition, GRAd-COV2 induced a robust and Th1-skewed T cell response against the S antigen in 89/90 subjects from both age groups. Overall, the safety and immunogenicity data from the phase 1 trial support further development of this vaccine.

**One Sentence Summary:** GRAd-COV2, a candidate vaccine for COVID-19 based on a novel gorilla adenovirus, is safe and immunogenic in younger and older adults

## INTRODUCTION

The coronavirus disease 2019 (COVID-19) pandemic is the most significant international public health emergency occurred over a century. COVID-19 is caused by a novel human viral agent, the Severe Acute Respiratory Syndrome Coronavirus 2 (SARS-CoV-2) that emerged in the People Republic of China at the end of 2019. Since its start, COVID-19 pandemic has already caused hundreds million cases of infections with about several million of fatalities worldwide. This unprecedented global health emergency has boosted international efforts to develop a vaccine and a multitude of platforms have been used including genetic vaccines based on mRNA, DNA and viral vectors. As of April 2021, there are 272 vaccine candidates. One hundred and eighty-six are in preclinical development and 86 in clinical development, of which 13 are either authorized in emergency use or approved by different countries (http://www.who.int/publications/m/item/draft-landscape-of-covid-19-candidate-vaccines). However, COVID-19 vaccines rolled out so far cannot meet the global vaccine demand and there is an urgent need to develop additional safe and effective vaccines able to ramp up production to be included in worldwide vaccination campaigns.

We developed GRAd-COV2, a vaccine candidate based on an adenoviral vector derived from a group C Gorilla adenovirus that is made replication defective by deleting the E1 genomic region. GRAd-COV2 vaccine encodes for a full-length SARS-COV-2 Spike (S) protein. The S protein is the primary vaccine target for most COVID-19 vaccines as it is critical for viral cell entry through the human ACE2 receptor *(1)*. The S gene cloned into GRAd-COV2 contains mutations that stabilize the S protein expressed in the host cell in a pre-fusion conformation for better exposure of epitopes important for antibody-mediated neutralization to the immune system *(2, 3)*. GRAd vector was selected based on its immunogenicity and manufacturability profile, in addition to the low frequency of anti-GRAd neutralizing antibodies in human sera compared with human adenovirus type 5 *(4)*. We recently reported that a single dose of GRAd-COV2 elicited strong immune responses, both humoral and cellular, in mice and rhesus macaques *(4)*. These preclinical data supported the start of a first-in-human, dose escalation, phase 1 clinical trial in healthy younger and older adults to evaluate the safety and immunogenicity of GRAd-COV2.

Here we report the trial interim analysis results up to 4 weeks after vaccination.

The vaccine was well tolerated at all three dose levels explored, 5×10^10^ (low dose), 1×10^11^ (intermediate dose) and 2×10^11^ (high dose) viral particles. Importantly humoral and T cell responses were readily induced at similar levels in the two age cohorts. Based on these results, we have initiated a phase 2/3 study that will evaluate the efficacy of either a single GRAd-COV2 administration or a repeated dose regimen (NCT04791423).

## RESULTS

### Volunteers screening and selection

Between August 11 and September 20, 2020, 181 potential volunteers were evaluated for eligibility. Volunteers were screened in excess for speeding up enrollment and to comply with a strict staggering enrollment scheme. Of the 181 subjects screened, 91 (45 in younger adults cohort aged 18-55 and 46 in the older adults cohort, aged 65-85) were vaccinated. Fifty-five volunteers (30 in younger age cohort and 25 in the older age cohort) were excluded as they did not meet inclusion criteria and 35 (25 younger age cohort and 10 in the older age cohort) were not vaccinated as the time window between screening and vaccination was trespassed. Median age, gender distribution and other characteristics were well balanced within study arms both in the younger (Fig. 1A) and in the older adults (Fig. 1B) cohorts.

**Fig. 1.**
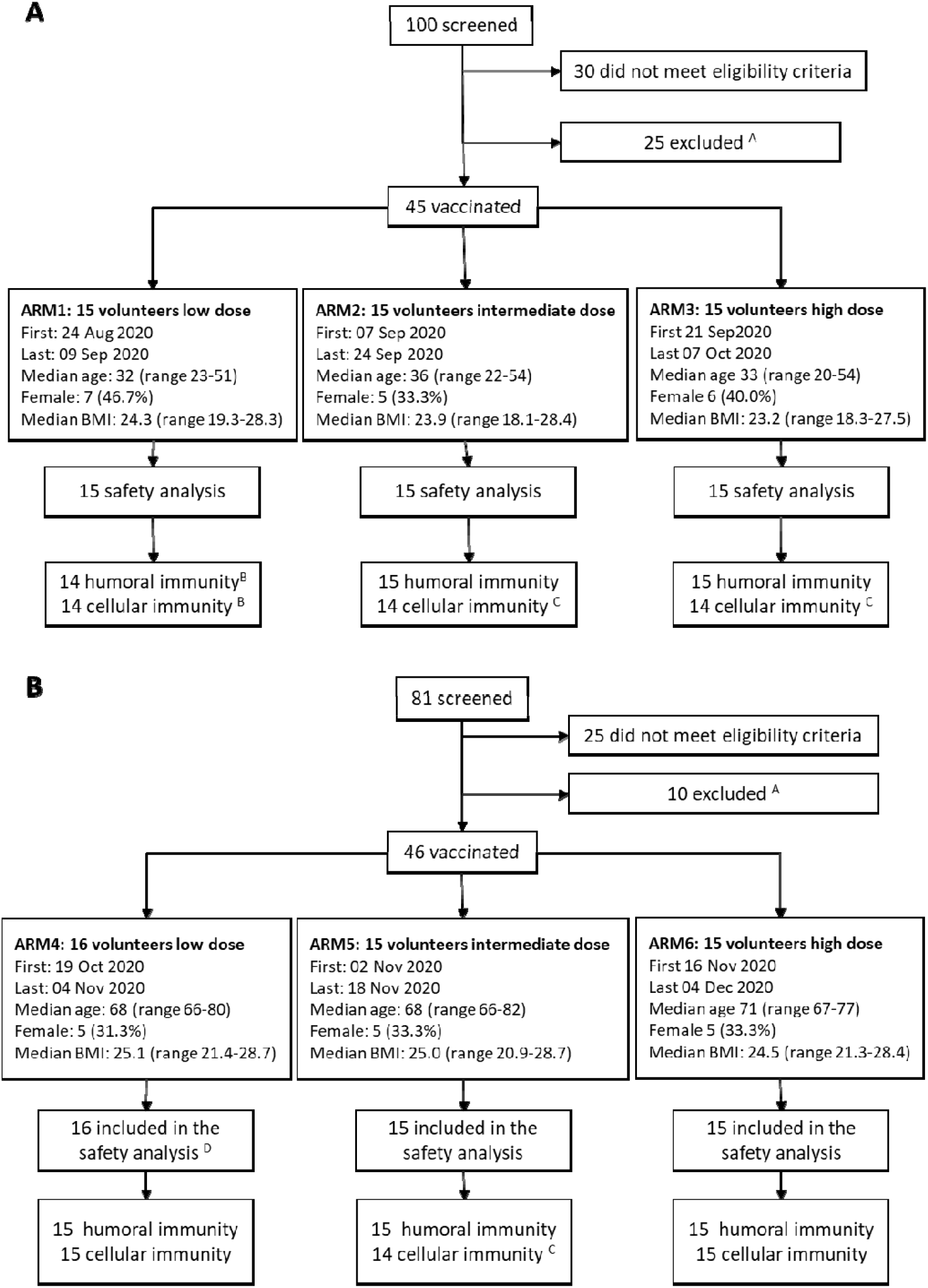
Study sample selection and analysis. Enrollment, study population characteristics, treatment and follow-up of the (**A**) 45 volunteers enrolled in younger age cohort (aged 18-55 years), and of the (**B**) 46 volunteers enrolled in older age cohort (aged 65-85 years). Recorded details include age at enrollment in years (median and range) and the body mass index (BMI). ^A^ volunteers not vaccinated as they lost the time window between screening and vaccination visit; ^B^ 1 volunteer excluded from humoral and cellular immunity analysis due SARS-CoV-2 infection immediately after/before vaccination; ^C^ volunteers excluded from ELISpot analysis only, because of high non-specific IFNγ secretion. ^D^ 1 volunteer excluded from immunogenicity analysis and replaced, due to unspecific reactivity in anti S CLIA assay.

One vaccinated arm 1 volunteer had detectable levels of specific anti-N and S IgG since week 1 after vaccination. As this finding suggested a natural (asymptomatic) infection with SARS-CoV-2, we excluded this subject from the immunogenicity analysis. One vaccinated arm 4 volunteer was found positive to anti-S IgG CLIA assay at baseline. The volunteer did not seroconvert to N, and was found negative when using a different assay (immunofluorescence), suggesting an unspecific reactivity to the CLIA. Being anti-S IgG CLIA, according to protocol, the main serological assay to follow vaccine induced anti-S IgG response, the volunteer was replaced with a subsequent one, and was included in the safety analysis but excluded from the immunogenicity assessment. Moreover, 3 additional volunteers (arms 2, 3 and 5) were excluded from IFNγ ELISpot analysis due to spontaneous IFNγ production above acceptability range (Fig. 1).

### Vaccine safety

Figure 2 reports the frequency of all adverse events by type, cohort and severity.

**Fig. 2.**
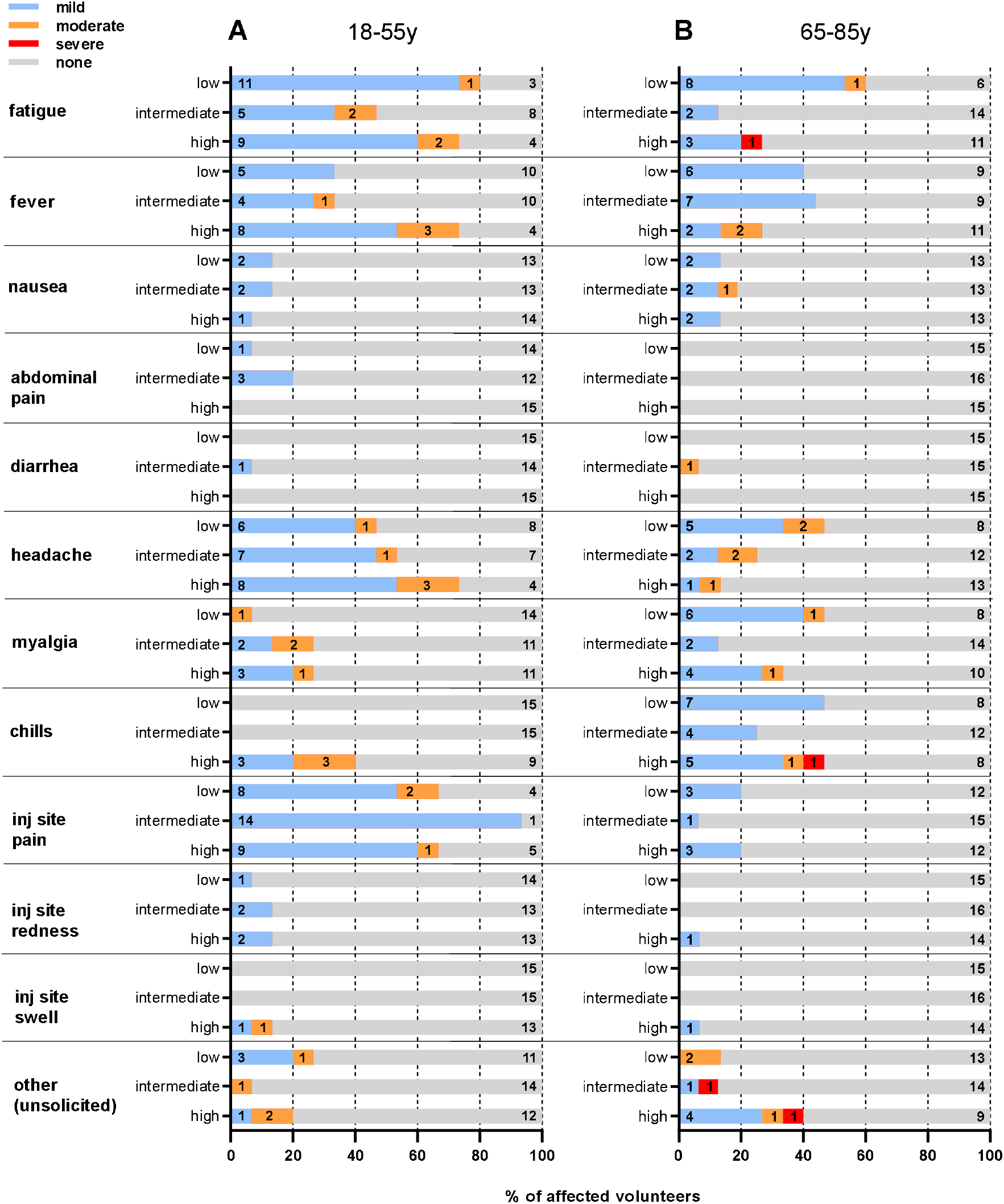
Adverse Event (AE) within 28 days after vaccination. The proportion of volunteers in each study arm experiencing the specific AE, and reported at the maximal severity, is shown. The severity of AE is reported according to intensity scale in the protocol. (**A**) Younger adults cohort. (**B**) Older adult cohort. Low: low-dose arm 5×10^10^ viral particles; Intermediate: intermediate-dose arm 1×10^11^ viral particles; High: high-dose arm 2×10^10^ viral particles.

A single intramuscular administration of GRAd-COV2 was well tolerated at all doses and in both cohorts. Overall, we observed 328 adverse events of any grade; of those, 255 were considered to be related with vaccination. In the younger age cohort we observed 143 solicited AEs (118 mild and 25 moderate) and 8 unsolicited AEs (4 mild and 4 moderate). In the older age cohort (we observed 104 solicited AEs (79 mild, 13 moderate and 2 severe) and 10 unsolicited AEs (5 mild, 3 moderate and 2 severe). Twenty-four volunteers (26%; 7 younger and 17 older) reported no AE, 44 volunteers (48%; 23 younger and 21 older) reported at least one mild AE and 21 volunteers (48%; 15 younger and 6 older) reported at least one moderate AE and 2 (2%; both older age volunteers) reported at least one severe adverse event. No serious AE was reported, and no pre-specified trial-halting rules were met. Nineteen volunteers (15 younger and 4 older) received antipyretics to control AEs. One older age volunteer received inhalator steroids for cough started the day after the vaccination that resolved in 9 days.

Most AEs occurred in the first 48 hours after vaccination and were short-lived (median time to resolution 24 hours, IQR less than 1 day-2 days).

Overall, no clinically significant blood count changes were observed. A transient reduction of neutrophils with concomitant increase in monocytes was detected at day 2 in the majority of volunteers in all age and vaccine dose groups that mostly reverted at pre-dose level by week 1 (Fig S1). Other hematological parameters were mostly unaffected (Fig. S1, lymphocytes and platelets shown for reference), and no vaccine-related trends were noted for biochemistry parameters.

### Binding antibodies to SARS-CoV-2 Spike

Antibody response to GRAd-COV2 vaccination was primarily monitored by a clinically validated CLIA, revealing similar kinetics of anti-S IgG induction in all study groups (Fig. 3A and table S1). Seroconversion to S occurred at week 2, when anti-S IgG became detectable in 23 (52%) out of 44 of the younger volunteers and in 15 (33%) out of 45 of the older age subjects. At week 4 after vaccination, high levels of anti-S IgG were measurable in 40 (91.0%) out of the 44 analyzed younger and in 42 (93.3%) out of the 45 older age volunteers. In the younger age cohort, three volunteers (1 in ID and 2 in HD arm) showed a weak increase of anti-S-IgG (10.8 AU/mL, 11.5 AU/mL and 12.8, respectively), that are below the diagnostic cutoff applied in the clinical practice (15.0 AU/mL). Only one volunteer in the LD arm showed anti-S-protein IgG below the assay limit of detection (LOD; <3.8 AU/mL). Also in the older age cohort, three volunteers (1 in LD, 1 in ID, 1 in HD arm) showed a weak increase of anti-S-IgG (5.0 AU/mL, 8.5 AU/mL and 9.7 respectively) that were below the diagnostic cutoff.

**Fig. 3.**
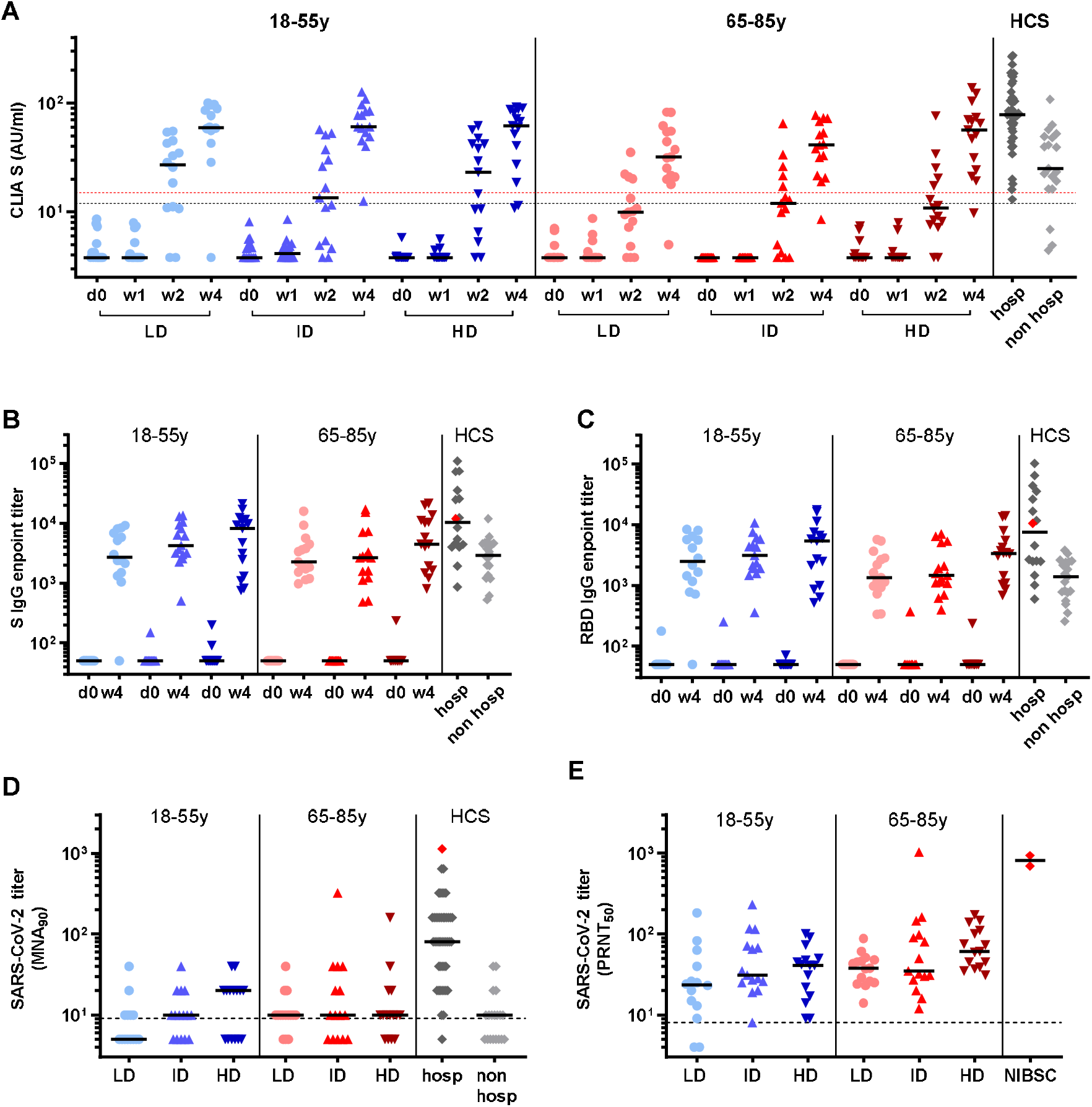
SARS-CoV-2 specific binding and neutralizing antibody responses in GRAd-COV2 vaccinated volunteers. Antibody response to SARS-CoV-2 induced by GRAd-COV2 vaccination at low dose (LD-5×10^10^ vp-circles), intermediate dose (ID-1×10^11^ vp-upright triangles) and high dose (HD-2×10^11^ vp-upside down triangles). Blue and red color shades identify younger and older age cohorts, respectively. Horizontal black lines are set at median across all panels. HCS: human convalescent sera (diamonds), obtained from either previously hospitalized (hosp-dark grey) or from non-hospitalized (non-hosp-light grey) COVID-19 patients, and the NIBSC 20/130 standard plasma (red diamond) are shown for reference. (**A**) IgG binding to S1-S2 measured by CLIA at day of vaccination (d0), and 1-2-4 weeks after vaccination. Data are expressed as arbitrary unit (AU)/ml. Blue and red dashed lines are set at 12 and 15 AU/ml. According to manufacturer, results >15 are clearly positive, between 12 and 15 are equivocal and <12 are negative or may indicate low level of IgG antibodies to the pathogen. (**B-C**) SARS-CoV-2 specific IgG titers in sera collected at d0 and w4 post vaccination measured by ELISA on recombinant full-length Spike (B) or RBD (C). Data are expressed as endpoint titer, and for negative sera where a titer cannot be calculated, an arbitrary value of 50 (or one-half the first serum dilution tested) is assigned. (**D-E**) SARS-CoV-2 neutralizing antibodies at week 4 post vaccination detected by SARS-CoV-2 microneutralization assay (D), or by plaque reduction neutralization test (E). SARS-CoV-2 neutralization titers are expressed as MNA_90_ and PRNT_50_, or the reciprocal of serum dilution achieving 90% or 50% neutralization, respectively. Dashed lines indicate LOD, and negative sera are assigned a value of one/half the LOD.

Antibodies measured by CLIA did not show a dose response relationship in the younger adult cohort (IgG levels at W4 were 59.5, 60.6 and 61.8 AU/mL for LD, ID and HD respectively, P=0.742). A dose response trend was observed in the older age cohort (IgG levels at W4 were 31.8, 41.0 and 56.3 AU/mL for LD, ID and HD respectively, P=0.315). However, despite the absence of a significant dose-response within the cohort, we observed that low and intermediate vaccine doses are associated to significantly lower IgG response in older age volunteers (LD arms median IgG was 59.4 in younger and 31.8 in older adults, P=0.011; ID arms median IgG was 60.6 in younger and 41.0 in older adults, P=0.011). Instead, high vaccine dose provided similar lever of IgG among the two age cohorts (high dose arms median IgG was 61.8 in younger and 56.3 in older adults, P=0.917).

ELISA assay showed that 89 (98.8%) out of 90 volunteers developed detectable level of anti-S IgG (including both antibodies against the whole Spike protein and specific anti-RBD) at 4 weeks after vaccination (Fig 3B-C and table S1).

Median titers to S and RBD showed a dose-response trend both in the younger and older age cohorts. We also observed that median level of IgG either against S or RBD were higher in younger than in older volunteers, although the difference between the two age cohorts were not significant (Fig. 3B-C and table S1).

None of the study volunteers showed seroconversion to N during the 4 weeks following vaccination, suggesting that no SARS-CoV-2 infection occurred.

### Neutralizing antibodies against SARS-CoV-2

Neutralizing antibodies to SARS-CoV-2 were assessed by two different *in vitro* assays, both using SARS-CoV-2 live virus.

Microneutralization assay (MNA_90_) at week 4 after vaccination detected neutralizing antibodies in the serum of 25/44 (56.8%) young volunteers and 33/45 (73.3%) older age volunteers (Fig. 3D). Anti-SARS-CoV-2 neutralizing antibodies were below the limit of detection in 8, 5 and 6 younger, and in 3, 5 and 4 older volunteers in the LD, ID and HD arms, respectively.

Plaque reduction neutralization test (PRNT_50_) revealed that SARS-CoV-2 neutralizing antibodies were detectable in 42/44 (92.5%) younger and in 45/45 (100%) older age volunteers (Fig. 3E). The two younger volunteers with undetectable levels of neutralizing antibody were in the LD arm. We observed a significant increase of the level of neutralizing antibodies across the arms in the older adult cohort (median PRNT_50_ titer 38.0, 35.0 and 61.0 in the LD, ID and HD arms respectively; P=0.048). A similar dose-response trend was also observed across the arms in the younger adults cohort (median PRNT_50_ titer 23.5, 31.0 and 41.0 in the LD, ID and HD arms respectively; P=0.197).

Across all arms, titers of binding and neutralizing antibodies elicited by GRAd-COV2 vaccination were in the range of those measured in subjects recovered from mild COVID-19 (Fig. 3 A-D and table S1).

### T cell response

Quantitative IFNγ ELISpot assay has been used to assess T cell response on freshly isolated PBMCs from volunteers in both cohorts at week 2 after vaccination.

GRAd-COV2 administration at all three doses induced potent S-specific IFNγ producing T cell response in both cohorts (Fig. 4A and table S1). Individual responses ranged between 87 and 10560 IFNγ SFC/million PBMC in younger adults and between 283 and 18877 in older subjects. Notably, 80% of evaluable subjects across the two age cohorts showed a response above 1000 SFC/million PBMC. Only one volunteer in arm1 did not show a detectable anti-S T cell response but still had measurable level of antibodies to S and RBD (ELISA titer of 1494 and 721 respectively).

**Fig. 4.**
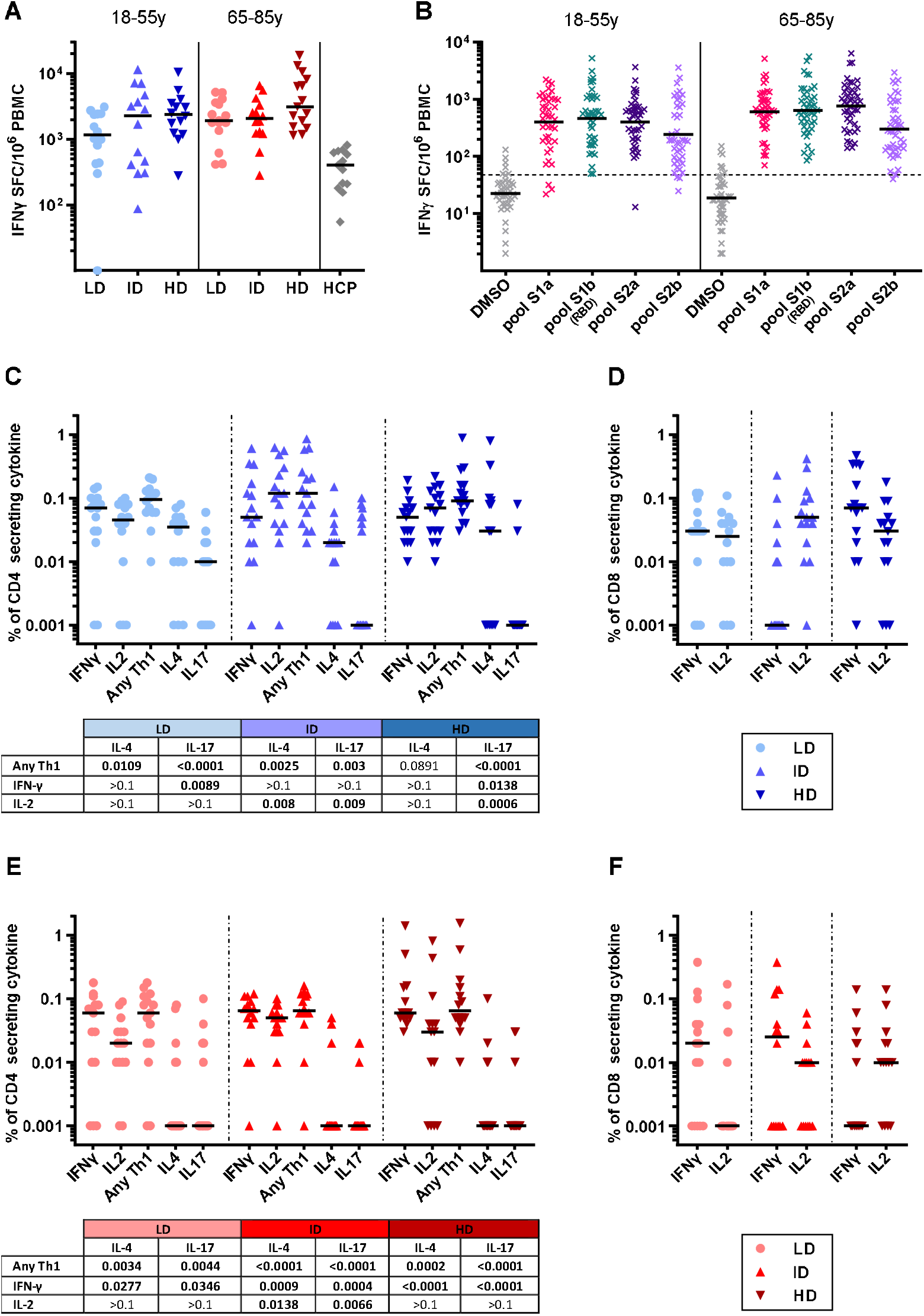
SARS-CoV-2 Spike specific T cell response in GRAd-COV2 vaccinated volunteers. T cell response to Spike peptides, induced by GRAd-COV2 vaccination at low (LD-5×10^10^ vp-circles), intermediate dose (ID-1×10^11^ vp-upright triangles) and high dose (HD-2×10^11^ vp-upside down triangles). Blue and red color shades identify younger and older age cohorts, respectively. Horizontal black lines are set at median across all panels. (**A-B**) IFNγ ELISpot on freshly isolated PBMC at w2. Data are expressed as IFN-γ spot forming cells (SFC)/10^6^ PBMC. In (A) individual data points represent cumulative Spike T cell response, calculated by summing the response to each S1a, S1b, S2a and S2b peptide pools stimulation and correcting for background (DMSO stimulation) in each volunteer. HCP: freshly isolated human convalescent PBMC, obtained from subjects recovering from symptomatic SARS-CoV-2 infection. (B) distribution of IFNγ ELISpot response to individual Spike peptide pools. Dashed line indicates assay positivity cut off (48 SFC/million PBMC). (**C-D-E-F**) IFNγ/IL2/IL4/IL17 intracellular staining and FACS analysis at w2 on fresh PBMC of younger (C-D) and older volunteers (E-F). Data are expressed as the percentage of Spike-specific CD4 (C-E) or CD8 (D-F) secreting each cytokine (or for Any Th1 a combination of cytokines, i.e. sum of CD4 secreting IFNγ alone, IL2 alone, and both IFNγ and IL2), obtained by summing responses to each of the 4 Spike peptide pools and corrected for background (DMSO stimulation). The tables below CD4 graphs shows P values derived by Kruskall-Wallis test comparing Th1/Th2/Th17 profile within each dose group.

Figure 4 shows that an incremental level of T cell response was observed in younger (median response 1162, 2857 and 2272 SFC/per million PBMC in the LD, ID and HD arms, respectively; P=0.154) and older (median response 1917, 2262 and 3142 SFC in the LD, ID and HD arms, respectively, P=0.206) volunteers. However, neither trend was associated to a significant dose-response within the cohorts. Moreover, we did not find significant differences between younger and older study arms receiving the same vaccine dose (P was 0.116, 0984 and 0.152 for LD, ID and HD, respectively).

T cell response induced by vaccination was directed to multiple epitopes, as most of the volunteers in both cohorts had a detectable response against all four S peptide pools analyzed (Fig. S2A and B). Moreover, we found that all regions of the S protein had a similar degree of immunogenicity in both age cohorts (Fig. 4B).

S-specific T cell response was generally higher in GRAd-COV2 vaccinated subjects than in SARS-CoV-2 convalescent controls who were sampled 1-2 months after symptoms onset.

Intracellular staining for cytokine production (ICS) and FACS analysis revealed that vaccine-induced responses involved both S protein specific CD4 and CD8 T lymphocytes in younger and older volunteers (Fig. 4C-D and E-F), with a slightly higher S-specific CD4 than CD8 T cell responses. Importantly, among GRAd-COV2 vaccine induced S-specific CD4, IFNγ production was more prominent than IL4 and IL17 in both age cohorts, indicating that the vaccine induced a predominantly T helper 1 (Th1) response (tables below Fig. 4C and 4E report comparisons).

### Anti-vector antibodies

GRAd neutralization was not set as a screening or a randomization parameter, but was anyway assessed as an exploratory endpoint at baseline and 4 weeks after vaccination. Out of 90 volunteers, we measured neutralization titers >200, a level above which an impact on vaccine immunogenicity has been reported, in only 10 volunteers (11%).

As expected, the vaccination with GRAd-COV2 induced or boosted anti-vector immunity in most volunteers (Fig. 5A), with no significant differences attributable to vaccine dose level or age.

**Fig. 5.**
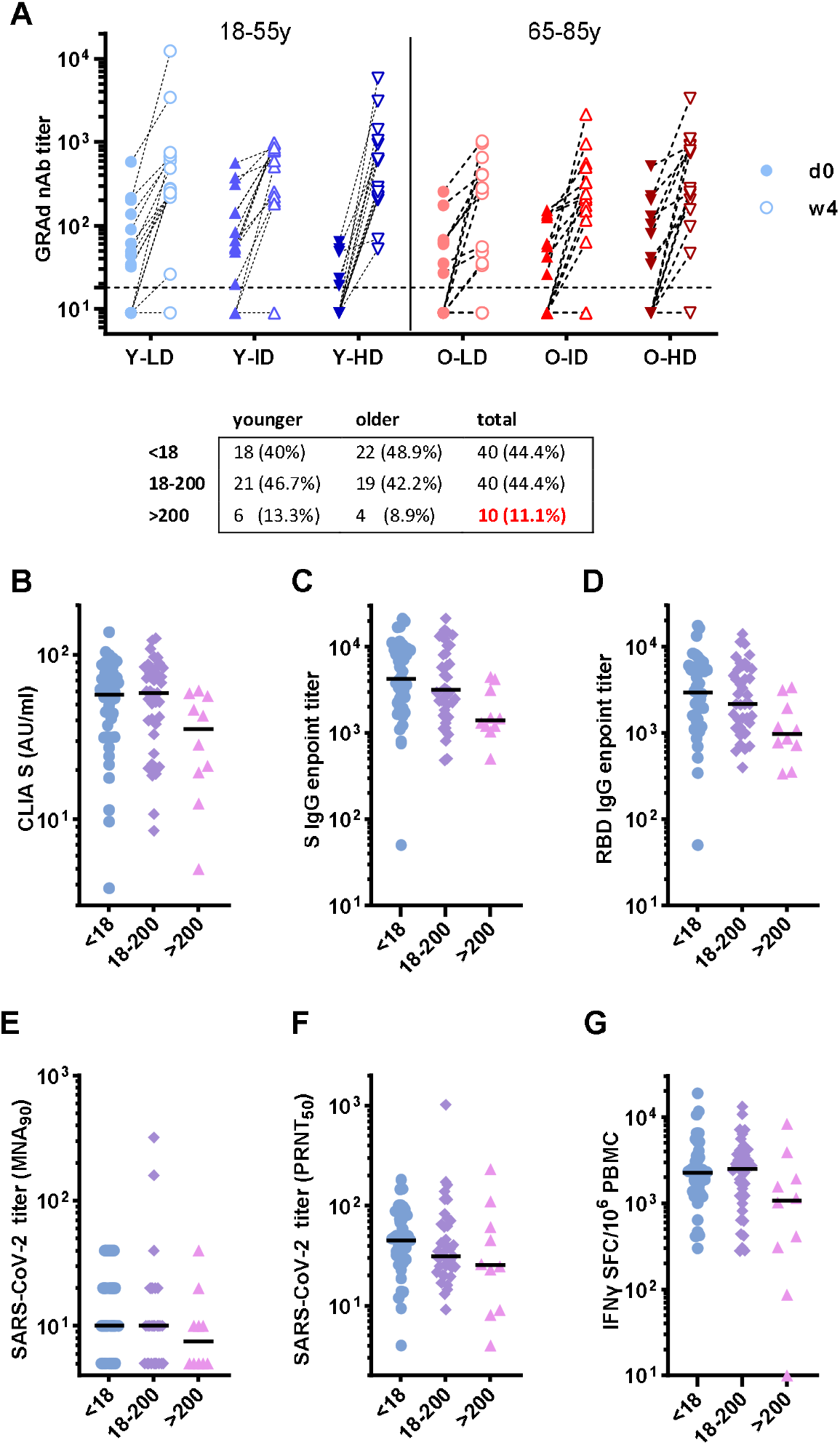
Neutralizing titers to GRAd before and after vaccination and impact on SARS-CoV-2 vaccine induced immune response. (**A**) Neutralizing titers to GRAd vector measured in sera of vaccinated volunteers the day of vaccination (d0-filled symbols) and 4 weeks after (w4-open symbols). The dashed lines set at 18 indicate the assay LOD. The table reports the number (and percentage) of sera with GRAd nAb titer in the indicated range (<18, 18-200 or >=200) in younger, older, and overall across the two age cohorts of volunteers (N=90). (**B-G**) Volunteers are stratified according their GRAd nAb serostatus at baseline irrespective of age cohort and vaccine dose received, and for each stratum the vaccine-induced immune response is presented: the binding antibody response at w4 by CLIA S IgG (B), S ELISA (C), RBD ELISA (D); the SARS-CoV-2 neutralizing Ab response at w4 by MNA_90_ (E), PRNT_50_ (F); the T cell response by IFNγ ELISpot at w2 (G).

To assess the impact of anti-GRAd immunity, pooled immunogenicity data at peak post vaccination (T, binding and neutralizing Ab responses) were stratified according to their baseline GRAd neutralizing titers (Fig. 5B-G). A trend for reduced S binding antibody response to the vaccine antigen was noted in volunteers with GRAd nAb >200 at baseline, with IgG titers measured by ELISA significantly lower (P=0.018 on full length S and P=0.0145 on RBD).

Similar trend was noted for T cell response and SARS-CoV-2 neutralizing Ab. Conversely, vaccine-induced immune responses in individuals with GRAd nAb titers in the 18-200 range (i.e. low but detectable) at baseline were undistinguishable or similar to those of volunteers with no anti-vector immunity.

### Correlative analysis of the immune response

A correlation matrix was computed to define correlations between the immunological readouts of this study (Fig.6 and associated table). A strong positive correlation was observed among all serological tests (neutralization, CLIA, anti-S and anti-RBD ELISA). Correlation was strongest amongst assays detecting binding Abs (CLIA and ELISA to S and RBD, r values between 0.89 and 0.94), and within the two neutralization assays (r=0.8). Correlation was significant but less strong within the two serology assay blocks (binding vs neutralizing, r between 0.5 and 0.66). Moreover, the T cell immunity significantly directly correlated with the serological assays (r=0.38 to 0.47), suggesting that GRAd-COV2 vaccination induces well-coordinated Ab and T cell responses to the encoded Spike. Of note and confirming the observation in previous paragraph, all vaccine induced immunological readouts trended to inversely correlate with the anti-vector antibody at baseline, reaching significance only for Ab responses as measured by ELISA (r= -0.24, P=0.026 for S, and r= -0.24 and P=0.024 for RBD).

**Fig. 6.**
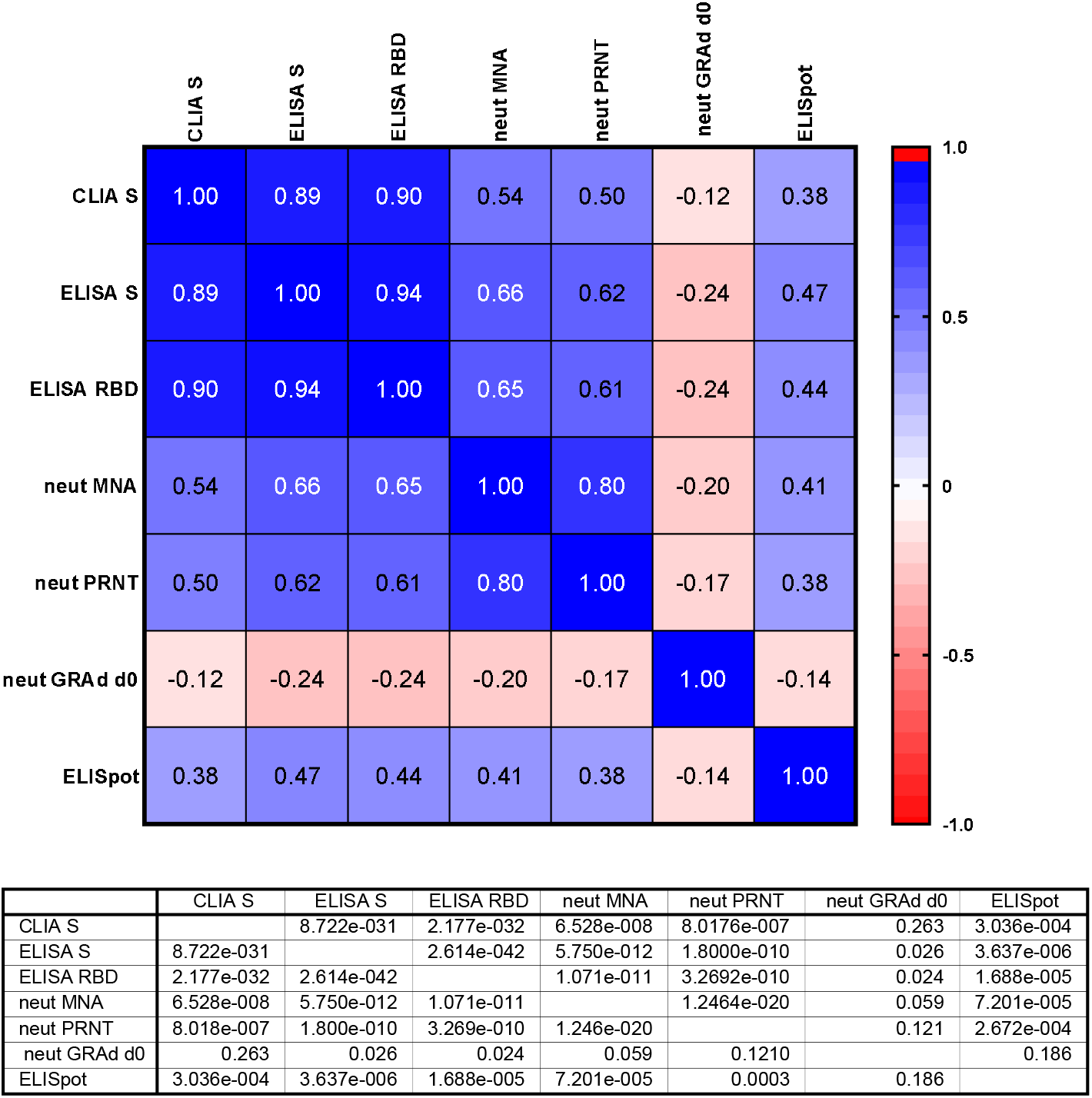
Correlation matrix of GRAd-COV2 immune response. A correlation (non parametric Spearman, two-tailed) matrix was computed for the full set of vaccinated volunteers irrespective of and age cohort vaccine dose received, comparing the measured anti-SARS-CoV-2 or anti vector immune response to evaluate how the different immune parameters correlate each other. The analysis was run on 87/90 volunteers for which data for all assays were available. Spearman r values are shown in the heatmap, and the table reports P values for each pair of variables.

## DISCUSSION

Here we report the first-in-human data on the safety and immunogenicity of a single administration of GRAd-COV2 given at different doses in healthy younger adults aged 18-55 and in healthy older adults aged 65-85 years. GRAd-COV2 is a COVID-19 candidate vaccine based on a replication-defective gorilla adenovirus vector. Our analysis provides evidence that GRAd-COV2 is well tolerated in both younger and older age cohorts at all three doses assessed. Indeed at 4 week of follow-up most of the observed adverse events were short lived and mild or moderate in intensity. Only 2 volunteers reported at least one severe adverse event and none reported serious adverse events. Solicited adverse events were less common in older age volunteers, similarly to what described for other COVID-19 vaccines *(5-7)*. In general, the adverse event profile did not differ from those reported in published work for other vector-based vaccines *(7-12)* while they were milder than those reported for mRNA formulated in liponanoparticles (LNP) and for adjuvanted vaccines *(2, 13, 14)*.

The vaccine was immunogenic at all doses, inducing seroconversion in the majority of participants of both age groups. Onset of immune responses was rapid with T cell responses already detectable 14 days after vaccination and antibodies titers increasing to day 28. We used two assays to measure neutralizing antibodies against live SARS-CoV2 in vaccinated volunteers, which exploit different virus strains and readouts. A microneutralization assay based on cytopathic effect (CPE) assessment provided evidence that 65% of the volunteers had measurable MNA_90_ titers, while a more sensitive plaque reduction assay could determine PRNT_50_ titers a in 98% of vaccinated volunteers. In general, titers of neutralizing antibodies to SARS-CoV-2 were lower than those reported in studies of vaccines based on mRNA *(13, 15, 16)* and on other viral vectors like Ad26 and ChAdOx1*(7, 9)*. However, the lack of standards and use of different assays complicate the comparison of performance of the various COVID-19 vaccines that are currently in use or in development. A comparison with convalescent serum panels stratified according to COVID-19 severity of the donors, showed that volunteers who received the GRAd-COV2 vaccine elicited similar titers of SARS-CoV-2 binding and neutralizing antibodies as did non-hospitalized people who had recovered from symptomatic COVID-19. In addition, the proportion of volunteers with detectable levels of neutralizing antibodies 28 days after vaccination seems not inferior to that reported in clinical studies after a single administration of other vaccines, including BNT162b2 *(5, 16)*, mRNA-1273 *(13, 17)*, Gam-COVID-Vac *(10)*, ChAdOx-1 *(8, 9)*, Ad-26 *(7)* and Ad5-vectored COVID-19 *(11, 12)*, suggesting that the measured lower nAb titers could result from different assay sensitivities.

There is a lack of knowledge on reliable immune correlates that would predict the efficacy of COVID-19 vaccines *(18-20)*. However, preliminary evidence from a non-human primate model *(21)*, clinical observation of the natural infection *(22)* as well as evidence of protective efficacy after first dose for several authorized vaccines *(5, 17, 23)* suggested that even low titer of neutralizing titers may be protective. The single administration of GRAd-COV2 elicits robust and Th1-skewed immune response, similar to that induced by a single dose of other vectored COVID-19 vaccines such as ChAdOx1 nCoV-19 *(8, 9)*, Ad5 *(11, 12)* and Ad26 *(7)*, and by two doses of mRNA-1273 *(13, 15)*. Indeed, a single administration of GRAd-COV2 elicits, within 2 weeks, a Th1 immune response that is broadly directed to multiple S protein epitopes. Importantly, a robust T cell response, such as that elicited by GRAd-COV-2, likely determines the duration of anti-COVID-19 immunity *(24, 25)* and prevents unintended immune reactions such as the vaccine-associated enhanced respiratory disease (VAERD) *(26, 27)*. Moreover, S protein-specific T cell responses directly correlated with binding and neutralizing antibodies, suggesting that GRAd-COV2 vaccine is able to shape a well-balanced and coordinated cellular and humoral specific immune response, as this naturally occurs in mild COVID-19 *(28, 29)*.

New SARS-CoV-2 variants are becoming predominant through a process of evolutionary selection that is not well understood. Among others, a variant of concern is the B.1.351 strain which is less effectively neutralized by convalescent plasma from COVID-19 patients and by sera from those vaccinated with several vaccines in development *(30-32)*. The decrement in neutralization can be more than 10-fold with convalescent plasma and averages 5-to 6-fold less with sera from vaccinated individuals. The interim data from a randomized placebo-controlled vaccine study, of rAd26 from Janssen in the US, Brazil, and South Africa showed efficacy against COVID-19 at 72%, 66%, and 57%, respectively (https://www.jnj.com/johnson-johnson-announcessingle-shot-janssen-covid-19-vaccine-candidatemet-primary-endpoints-in-interim-analysis-of-itsphase-3-ensemble-trial). Viral sequence data from infected patients showed that the B.1.351 strain was responsible for the majority of infections in South Africa. Lower vaccine efficacy in the South Africa cohort could be related to antigenic variation or to geographic or population differences. Interestingly, despite the reduced efficacy, the single dose of rAd26 vaccine was 85% effective overall in preventing severe COVID-19, and protection was similar in all regions. Given the important reduction of neutralizing activity on the B.1.351 variant, it is tempting to speculate that the vaccine-induced T cell responses might be predominant in protecting from severe disease.

As for other candidate vaccines using viral vectors, pre-existing immune responses against the vector can compromise the induction of an immune response against the target antigen *(33, 34)* and indeed this was shown also for a candidate COVID-19 vaccine based on the common human Ad5 serotype *(11)*. In our study, high levels of neutralizing antibodies against GRAd-COV2 were associated with a reduced median T cell response and antibody levels against S protein. Albeit significant, the presence of pre-existing anti-GRAd antibodies did not abrogate the immune response in most of the volunteers. These findings highlight the potential utility of vectors such as GRAd, for which pre-existing immunity is infrequent in the human population.

Our interim analysis indicates that vaccine candidate GRAd-COV2 is safe and immunogenic in both younger and older adults. This finding has supported and guided our decision to proceed with an ongoing phase 2/3 trial (NCT04791423) to evaluate the efficacy of either a single dose or of a two-dose regimen of GRAd-COV2.

Indeed, experience from other ongoing clinical trials on adenovirus-based vaccines provided evidence that a second administration of vaccine is safe and can significantly improve the production of neutralizing antibodies *(9, 10)*. The observed increase of anti-GRAd neutralizing antibody titers post vaccination is in line with that observed with other adenovirus based COVID-19 vaccines *(6, 7)* which did not prevent boosting of anti-S antibodies upon a second administration. It is worth noticing that most of the anti-COVID-19 vaccines need a prime boost scheme, including non-replicating viral vectored vaccines, mRNA and adjuvanted subunit vaccines.

This study has some limitations due to the low number of volunteers per arm (N=15), and to the lack of subject randomization amongst study arms on the basis of GRAd neutralizing titer at baseline. Both these aspects could have had an impact in the poor vaccine dose effect observed. However, the explored dose range (4-fold between low and high dose) was limited, and no major dose effect was expected. Nevertheless, for phase 2/3 single-dose regimen we selected the 2×10^11^ vp (high) dose, due to higher and more consistent immunogenicity especially in the elderly cohort. As for the two-dose regimen, the intermediate dose of 1×10^11^ vp was selected which represents the best compromise between tolerability and immunogenicity. Finally, data beyond w4 post vaccination on persistence of humoral and cellular responses will be part of a future report once the study follow up will be completed.

## MATERIALS AND METHODS

### Study design

This study is a phase 1, dose-escalation, open label clinical trial designed to determine the safety and immunogenicity of GRAd-COV2. The study included two age cohorts, of either younger (18-55) or older (65-85) adults. Each cohort consisted of 3 arms of 15 volunteers each, for assessing a single administration at three different dose levels of GRAd-COV2: low dose (LD) 5×10^10^; intermediate dose (ID) 1×10^11^ and high dose (HD) 2×10^11^ viral particles (vp). We report here the safety and immunogenicity endpoints collected in the first 4 weeks after vaccination for the volunteers enrolled in both age cohorts, as foreseen in the study Protocol (Interim analyses 1 and 2). The full study protocol is provided in Supplementary material.

Volunteers and settings: eligible participants were healthy adults aged 18 to 55 years with no history of COVID-19, no laboratory findings suggestive of current or previous infection with SARS-COV-2 and who have attended the screening visit no more than 21 days before vaccination (appendix 1 lists all eligibility criteria).

Trial procedures: GRAd-COV2 was manufactured by ReiThera under good manufacturing practice conditions, suspended in formulation buffer at a concentration of 2 × 10^11^ vp/mL. Volunteers received a single intramuscular injection in the deltoid. For administration of the HD, 1ml of GRAd-COV2 was injected without dilution. For ID and LD, the vaccine was diluted in sterile saline solution to reach a final 1ml injection volume.

The dose-escalation staggered enrollment included 3 sentinel volunteers in the LD arm followed by enrollment of the full LD arm. ID and HD sentinel volunteers were enrolled 7 days after that safety data of LD or ID arm were available, respectively. All the enrollment, stages were supervised by an independent data safety monitoring board. Volunteers recorded local and systemic reactions on a diary card for 28 days. The severity and relatedness with vaccination or adverse events were assessed by the medical team in each center.

### Specimens from SARS-COV-2 convalescent patients

As comparator for immunogenicity analysis we used three independent sets of anonymized specimens (sera and PBMC) from COVID-19 patients either hospitalized or recovering from mild symptomatic disease, collected 20 to 60 days after symptom onset. The Research reagent for anti-SARS-CoV-2 Ab (NIBSC code 20/130), a human plasma from a donor recovered from COVID-19, was included as a positive control.

### Ethical statement

All participants provided written informed consent before enrolment. The trial was conducted at the National Institute for Infectious Diseases Lazzaro Spallanzani (INMI) in Rome and at Centro Ricerche Cliniche in Verona (CRC-Verona). The study was conducted according to the Declaration of Helsinki, and approved by the Italian Regulatory Drug Agency (AIFA) and the Italian National Ethical Committee for COVID-19 clinical studies (ClinicalTrials.gov NCT04528641; EudraCT 2020-002835-31).

Sera from convalescent patients who resolved SARS-CoV-2 infections came from residual specimens used for diagnostic purposes and were utilized according to INMI protocols for observational studies approved form internal ethical committee.

### SARS-CoV-2 anti-Spike and anti-Nucleoprotein IgG high throughput Chemiluminescence Immunoassay

Two commercial Ig assays were used according to manufacturer’s protocols: 1) DiaSorin LIAISON® SARS-CoV-2 S1/S2 IgG test on LIAISON® XL analyzers, a chemiluminescence immunoassays (CLIA) detecting anti-S1/S2 IgG (IgG antibody concentrations expressed as arbitrary units, AU/mL >= 15 are considered positive, DiaSorin, Italy); 2) Abbott SARS-CoV-2 assay on Abbott ARCHITECT® i2000sr, a chemiluminescence microparticle assay (CMIA) detecting anti-N IgG (index value Sample/Cut-off >= 1.4 is considered positive; Abbott Diagnostics, Chicago, IL, USA). The laboratory turnaround/determination is of 30 minutes for both CLIA and CMIA.

### SARS-CoV-2 Spike and RBD ELISA

SARS-CoV-2 ELISA was developed using as coated antigens either SARS-CoV-2 full length soluble prefusion stabilized Spike protein (expressed in Expi293 cells at ReiThera) or a recombinant RBD (expressed in HEK293 cells, ACROBiosystems). Proteins were coated on NUNC Maxisorp plates (ThermoFisher Scientific) at optimized concentration (5 μg/ml for R121, 2,5 μg/ml for RBD) in PBS overnight at 4°C. The following day, plates were washed with PBS 0,05% Tween-20 (PBS-T), then blocked with PBS-T+3% non-fat dry milk for 1,5h at 25°C in shaking. After wash, serum dilution curves (six 3-fold serial dilution of human sera from 1:100 to 1:24,300) prepared in PBS-T+1% non-fat dried milk were plated and incubated for 2h at 25°C in shaking. Plates were then washed and incubated with 1:2000 diluted anti-human IgG (Fc-specific, Sigma) for 1h at 25°C in shaking. Plates were then washed one last time and incubated with alkaline phosphatase substrate SigmaFast (Sigma) at 25°C. Absorbance was read at 405 and 620 nm using EnSight multiple plate reader (PerkinElmer), at 10, 20 and 30 minutes; the read with the best R-squared value was used to calculate endpoint titers. The endpoint titer was defined as the highest serum dilution that resulted in an absorbance value (OD-optical density) just above the calculated background of 4-fold the OD from secondary antibody alone. A COVID-19 convalescent patient’s serum at 1:200 and 1:6400 dilution was included as positive control in each plate to ensure inter-assay reproducibility. The research reagent for anti-SARS-CoV-2 Ab NIBSC code 20/130 was used as standard reference in each experiment.

### SARS-CoV-2 microneutralization assay (MNA)

Sera collected from vaccinated volunteer or convalescent COVID-19 patients were heat-inactivated at 56°C for 30 minutes (min), and titrated in duplicate in 7 two-fold serial dilutions (starting dilution 1:10). Equal volumes of 100 TCID_50_ SARS-CoV-2 (strain 2019-nCoV/Italy-INMI1; GISAID accession ID: EPI_ISL_412974) and serum dilutions, were mixed and incubated at 37 °C 5% CO_2_ for 30 min. Subsequently, 96-well tissue culture plates with sub-confluent Vero E6 cell monolayers were infected with 100 μl/well of virus-serum mixtures and incubated at 37 °C and 5% CO_2_. To standardize inter-assay procedures, positive control samples showing high (1:160) and low (1:40) neutralizing activity were included in each MNA session. After 48 hours, microplates were observed by light microscope for the presence of CPE. The supernatant of each plate was carefully discarded and 120 μl of a Crystal Violet solution containing 2% Formaldehyde was added to each well. After 30 min, the fixing solution was removed by washing with tap water and cell viability measured by photometer at 595 nm (Synergy(tm) HTX Multi-Mode Microplate Reader, Biotek). The highest serum dilution inhibiting at least 90% of the CPE was indicated as the neutralization titre and expressed as the reciprocal of serum dilution (MNA_90_). Serum from the National Institute for Biological Standards and Control, UK (NIBSC) with known neutralization titer was used as reference in MNA (Research reagent for anti-SARS-CoV-2 Ab NIBSC code 20/130).

### SARS-CoV-2 plaque reduction neutralization test (PRNT)

As an exploratory endpoint, the SARS-CoV-2 PRNT_50_ titers of vaccinated volunteers’ sera was determined by means of a plaque reduction neutralization test (PRNT) developed and run at Viroclinics Biosciences. Briefly, a standard number of SARS-CoV-2 (Bav/Pat1/2020 strain) infectious units were incubated with eight two-fold serial dilutions of heat inactivated sera, starting from 1:8 and up to 1:1024. After 1h pre-incubation, the virus/serum mixtures were inoculated on VeroE6 cells for 1 hour, than washed, replaced with infection medium and the cells were left overnight. After 16-24h the cells were formalin-fixed, permeabilized with ethanol, and incubated with primary anti SARS-CoV-2 Nucleocapsid monoclonal antibody followed by a secondary anti-mouse IgG peroxidase conjugate and TrueBlue substrate, which forms a blue precipitate on positive cells. Images of all wells were acquired by an ImmunoSpot analyzer (Cellular Technology Limited-CTL), equipped with software capable to accurately count the virus positive cells. The 50% neutralization titers were calculated according to a method described earlier *(35)*. The NIBSC standard 20/130 was tested as reference in the same run than the RT-COV-2 trial serum samples, with resulting PRNT_50_ of 697 and 934 for younger and older adults assay runs, respectively.

### GRAd Neutralizing Antibody Assay

Neutralizing antibody (nAb) titers in human sera were assayed as previously described *(36)*. Briefly, 8×10^4^ HEK293 cells per well were seeded in 96 well plates the day before the assay. GRAd vector encoding for the reporter gene secreted alkaline phosphatase (SEAP) at a pre-optimized multiplicity of infection (MOI) was preincubated for 1h at 37°C alone or with serial dilutions of control or test serum samples and then added to the 80-90% confluent HEK293 cells. After incubation for 1h at 37°C, the serum/infection mix was removed and replaced with 10% FBS in DMEM. SEAP expression was measured 24 hours later in cell supernatant by means of the chemiluminescent substrate from the Phospha-Light(tm) kit (Applied Biosystems). Neutralization titers were defined as the dilution at which a 50% reduction of SEAP activity from serum sample was observed relative to SEAP activity from virus alone.

### PBMC isolation and stimulation

Peripheral venous blood (40 ml) was collected in 7 ml lithium-heparin Vacutest blood collection tubes (Kima). Peripheral blood mononuclear cells (PBMC) were isolated from peripheral blood by density gradient centrifugation (Histopaque 1077, Sigma Aldrich). After separation, PBMC were suspended in RPMI 1640 (Sigma Aldrich) supplemented with 10% heat-inactivated highly defined fetal bovine serum (FBS) (HyClone), 2 mmol/L L-glutamine, 10 mmol/L HEPES buffer (N-2-hydroxyethylpiperazine-N-2-ethane sulfonic acid, Sigma Aldrich), and 100 U/ml penicillin, and 100 µg/mL streptomycin (Gibco), hereafter termed R10. PBMC count and viability were performed by using Guava Muse (Luminex). A set of 316 15mer peptides overlapping by 11 amino acids (synthetized by Elabscience Biotech Inc, and distributed by TEMA RICERCA), designed to cover the full-length S protein, was arranged into 4 pools (S1a, S1b-including RBD domain, S2a and S2b). PBMC (2×10^6^/ml) were stimulated with the four Spike peptides pools for 18 hours (3μg/ml each peptide final concentration). For flow-cytometry experiments, brefeldin A (10ug/ml, Sigma Aldrich) was added to block endoplasmic reticulum and Golgi apparatus.

### IFNγ ELISpot assay

The frequency of IFNγ-producing T cells was assessed by enzyme-linked immunosorbent spot-forming cell assay (ELISpot) after specific stimulation. PBMCs were resuspended in R10, stimulated with peptides pools, as described above, and plated at 2 × 10^5^ cells/well in ELISpot plates (Human IFN-γ ELISpot plus kit; Mabtech). PBMCs were incubated for 18-20 hours with 5% of CO_2_. At the end of incubation, the ELISpot assay was developed according to manufacturer’s instructions. Spontaneous cytokine production (background) was assessed by incubating PBMC with DMSO, the peptides diluent (Sigma,). Results are expressed as spot forming cells (SFC)/10^6^ PBMCs in stimulating cultures after subtracting spontaneous background. A result was considered positive if matching two criteria: i) higher than >48 SFC/10^6^ PBMC and ii) higher than 3 times Background. Data from three volunteers were excluded from all ELISpot analyses since their spontaneous IFNγ secretion in DMSO wells (yLD-101027, 450 SFC; yID-101034, 465 SFC; oID-101072, 233 SFC on DMSO) was above the mean+2standard deviation of the study population (mean=42 SFC, SD=72.27, mean+2SD=186 SFC/million PBMC), leading to unreliable quantitative analysis.

### Intracellular staining and flow-cytometry

Intracellular flow cytometry was performed by using ViaKrome 808 Fixable Viability dye, anti-IL17a Alexa Fluor 700, anti-CD45 Krome Orange, (Beckman Coulter), anti-CD3 BUV661, anti-CD8 PercP, anti-CD4 V450, anti-IL2 FITC, anti-IL4 PE (BD Biosciences), and anti-IFN-γ PE-Vio770 (Milteny Biotec). Briefly, PBMC were washed in DPBS (Corning) and stained with ViaKrome 808 Fixable Viability dye, anti-CD45, anti-CD3, anti-CD8, anti-CD4 for 15 minutes at 4°C. After washing cells were fixed with PFA 1% (Electron Microscopy Science), then stained with anti-IL17a, anti-IL2, anti-IL4 and anti-IFN-γ in PBS (Corning) BSA 0.5% (Sigma Aldrich) and Sodium Azide 0.01% (Serva Serving Scientists), saponin 0.5% (Sigma Aldrich) for 30 minutes at room temperature. Acquisition of 200,000 events was performed in the CD3+ gated population on CytoFLEX LX flow-cytometer, and analyzed with CytExpert software (Beckman Coulter). Data were analyzed with CytExpert software (Beckman Coulter: doublets were excluded in the FSC-H/FSC-A dot plot. Live cells were selected as Viakrome neg cells. In the immunological plot (SideScatter (SSC)/CD45), the CD45+ cells were gated, followed by gating CD3+ lymphocytes. Among the CD3+ cells, CD4+ or CD8+ cells were selected and the percentage CD4 or CD8 cells producing cytokines were evaluated. Spontaneous cytokines production (DMSO stimulation) was subtracted.

### Statistical analysis

This is a descriptive phase I first in human trial to assess safety and immunogenicity of three different vaccine doses and no formal sample size calculation is carried out. Categorical variables including occurrence of adverse events (AE) and detectable levels of binding and neutralizing antibody against SARS-CoV-2 and GRAd were reported as proportions. Continuous variables including results of CLIA anti-S IgG, MNA_90_ titers, PNRT_50_ titers and ELISpot values for IFNγ secretion were reported as median and interquartile range. Comparison of medians across arms and the impact of pre-existing immunity against GRAd were evaluated by two-tailed Kruskal–Wallis one-way variance analysis. Association between categorical variable was carried out by Fisher exact test. Correlations between immunogenicity assays were assessed by non-parametric Spearman’s rank tests.

## Data Availability

Full dataset can be required and will be availbale on request by 1 year after the end of the study

## Supplementary Materials

Fig. S1. Hematological parameters over time.

Fig. S2. Breadth of GRAd-COV-2 induced T cell response.

Table S1. Median and Interquartile range of main immunological measures.

RT-CoV-2 study Protocol

## GRAd study group

Sandrine Ottou (INMI), Serena Vita (INMI), Alessandra Vergori (INMI), Alessandra D’Abramo (INMI), Antonella Petrecchia (INMI), Chiara Montaldo (INMI), Emilio Scalise (INMI), Germana Grassi (INMI), Rita Casetti (INMI), Veronica Bordoni (INMI), Stefania Notari (INMI), Francesca Colavita (INMI), Silvia Meschi (INMI), Daniele Lapa (INMI), Licia Bordi (INMI), Silvia Murachelli (INMI), Teresa Tambasco (INMI), Alessandra Grillo (INMI), Erminia Masone (INMI), Elisa Marchioni (INMI), Dorian Bardhi (INMI), Ottavia Porzio (Ospedale Pediatrico Bambino Gesù), Francesca Cocca (Ospedale Pediatrico Bambino GesuSilvia Murachelli (INMI), Irene Turrini (CRC-Verona), Feliciana Malescio (CRC-Verona), Luigi Ziviani (CRC-Verona), Rita Lawlor (CRC-Verona), Federica Poli (CRC-Verona), Federica Martire (CRC-Verona), Daniela Zamboni (CRC-Verona), Flavia Mazzaferri (CRC-Verona).

## Acknowledgments

This Phase 1 study was through the participation of clinical trial volunteers. We are profoundly grateful to the volunteers for their effort.

We are grateful to the members of the Phase 1 Committee at the National Institute of health (ISS), of the technic-scientific committee of the Italian Drug Regulatory Agency (AIFA) and to the National Ethical Board for COVID-19.

We thank the members of the Data Safety and Monitoring Board for the priceless efforts spent for supporting the investigators on assessing information and for advising on trial continuation: Franco Locatelli (President), Paolo Antonio Grossi (Member), Sergio Bonini (Member) and Paolo Bruzzi (Member).

We thank the Prof. Rino Rappuoli for the support for interpretation of the interim analysis results.

We finally thank the ReiThera GMP and PD staff.

## Author contributions

Conceptualization: SL, SCa, AF, GI. Methodology: SL, SCa, AA, SM, EN, RC, CA, CC, SB, EG, GI, AF, GK, AV, SCo, MM, YS. Investigation: SL, SCa, AA, SM, EN, CA, CC, FG, SB, ASo, AMC, FB, MG, AR, ASa, GM, RG, EC, LS, MMP, ADL, EG, GK, MP. Visualization: SL, SCa

Funding acquisition: GI, EG, SCo, AF, MP. Project administration: FM, MMP, EG, FV, MS. Supervision: GI, FV, VA, FN, AV, AF, SCo. MRC. Writing – original draft: SL, SCa. Writing – review & editing: CA, CC, GK, MM, AV, GI, AF.

## Data and materials availability

All data are available in the main text or the supplementary materials. This trial is registered with ClinicaTrials.gov, NCT04528641.

## References and Notes

1. L. Dai, G. F. Gao, Viral targets for vaccines against COVID-19. Nat Rev Immunol 21, 73–82 (2021).

2. J. Pallesen, N. Wang, K. S. Corbett, D. Wrapp, R. N. Kirchdoerfer, H. L. Turner, C. A. Cottrell, M. M. Becker, L. Wang, W. Shi, W. P. Kong, E. L. Andres, A. N. Kettenbach, M. R. Denison, J. D. Chappell, B. S. Graham, A. B. Ward, J. S. McLellan, Immunogenicity and structures of a rationally designed prefusion MERS-CoV spike antigen. Proc Natl Acad Sci U S A 114, E7348–E7357 (2017).

3. D. Wrapp, N. Wang, K. S. Corbett, J. A. Goldsmith, C. L. Hsieh, O. Abiona, B. S. Graham, J. S. McLellan, Cryo-EM structure of the 2019-nCoV spike in the prefusion conformation. Science 367, 1260–1263 (2020).

4. S. Capone, A. Raggioli, M. Gentile, S. Battella, A. Lahm, A. Sommella, A. M. Contino, R. A. Urbanowicz, R. Scala, F. Barra, A. Leuzzi, E. Lilli, G. Miselli, A. Noto, M. Ferraiuolo, F. Talotta, T. Tsoleridis, C. Castilletti, G. Matusali, F. Colavita, D. Lapa, S. Meschi, M. Capobianchi, M. Soriani, A. Folgori, J. K. Ball, S. Colloca, A. Vitelli, Immunogenicity of a new gorilla adenovirus vaccine candidate for COVID-19. bioRxiv, 2020.2010.2022.349951 (2020).

5. F. P. Polack, S. J. Thomas, N. Kitchin, J. Absalon, A. Gurtman, S. Lockhart, J. L. Perez,G. Perez Marc, E. D. Moreira, C. Zerbini, R. Bailey, K. A. Swanson, S. Roychoudhury,K. Koury, P. Li, W. V. Kalina, D. Cooper, R. W. Frenck, Jr., L. L. Hammitt, O. Tureci,H. Nell, A. Schaefer, S. Unal, D. B. Tresnan, S. Mather, P. R. Dormitzer, U. Sahin, K. U. Jansen, W. C. Gruber, C. C. T. Group, Safety and Efficacy of the BNT162b2 mRNA Covid-19 Vaccine. N Engl J Med 383, 2603–2615 (2020).

6. M. N. Ramasamy, A. M. Minassian, K. J. Ewer, A. L. Flaxman, P. M. Folegatti, D. R. Owens, M. Voysey, P. K. Aley, B. Angus, G. Babbage, S. Belij-Rammerstorfer, L. Berry,S. Bibi, M. Bittaye, K. Cathie, H. Chappell, S. Charlton, P. Cicconi, E. A. Clutterbuck, R. Colin-Jones, C. Dold, K. R. W. Emary, S. Fedosyuk, M. Fuskova, D. Gbesemete, C. Green, B. Hallis, M. M. Hou, D. Jenkin, C. C. D. Joe, E. J. Kelly, S. Kerridge, A. M. Lawrie, A. Lelliott, M. N. Lwin, R. Makinson, N. G. Marchevsky, Y. Mujadidi, A. P. S. Munro, M. Pacurar, E. Plested, J. Rand, T. Rawlinson, S. Rhead, H. Robinson, A. J. Ritchie, A. L. Ross-Russell, S. Saich, N. Singh, C. C. Smith, M. D. Snape, R. Song, R. Tarrant, Y. Themistocleous, K. M. Thomas, T. L. Villafana, S. C. Warren, M. E. E. Watson, A. D. Douglas, A. V. S. Hill, T. Lambe, S. C. Gilbert, S. N. Faust, A. J. Pollard,C. V. T. G. Oxford, Safety and immunogenicity of ChAdOx1 nCoV-19 vaccine administered in a prime-boost regimen in young and old adults (COV002): a single-blind, randomised, controlled, phase 2/3 trial. Lancet 396, 1979–1993 (2021).

7. J. Sadoff, M. Le Gars, G. Shukarev, D. Heerwegh, C. Truyers, A. M. de Groot, J. Stoop, S. Tete, W. Van Damme, I. Leroux-Roels, P. J. Berghmans, M. Kimmel, P. Van Damme, J. de Hoon, W. Smith, K. E. Stephenson, S. C. De Rosa, K. W. Cohen, M. J. McElrath, E. Cormier, G. Scheper, D. H. Barouch, J. Hendriks, F. Struyf, M. Douoguih, J. Van Hoof, H. Schuitemaker, Interim Results of a Phase 1-2a Trial of Ad26.COV2.S Covid-19 Vaccine. N Engl J Med, (2021).

8. P. M. Folegatti, M. Bittaye, A. Flaxman, F. R. Lopez, D. Bellamy, A. Kupke, C. Mair, R. Makinson, J. Sheridan, C. Rohde, S. Halwe, Y. Jeong, Y. S. Park, J. O. Kim, M. Song, A. Boyd, N. Tran, D. Silman, I. Poulton, M. Datoo, J. Marshall, Y. Themistocleous, A. Lawrie, R. Roberts, E. Berrie, S. Becker, T. Lambe, A. Hill, K. Ewer, S. Gilbert, Safety and immunogenicity of a candidate Middle East respiratory syndrome coronavirus viral-vectored vaccine: a dose-escalation, open-label, non-randomised, uncontrolled, phase 1 trial. Lancet Infect Dis 20, 816–826 (2020).

9. P. M. Folegatti, K. J. Ewer, P. K. Aley, B. Angus, S. Becker, S. Belij-Rammerstorfer, D. Bellamy, S. Bibi, M. Bittaye, E. A. Clutterbuck, C. Dold, S. N. Faust, A. Finn, A. L. Flaxman, B. Hallis, P. Heath, D. Jenkin, R. Lazarus, R. Makinson, A. M. Minassian, K. M. Pollock, M. Ramasamy, H. Robinson, M. Snape, R. Tarrant, M. Voysey, C. Green, A. D. Douglas, A. V. S. Hill, T. Lambe, S. C. Gilbert, A. J. Pollard, C. V. T. G. Oxford, Safety and immunogenicity of the ChAdOx1 nCoV-19 vaccine against SARS-CoV-2: a preliminary report of a phase 1/2, single-blind, randomised controlled trial. Lancet 396, 467–478 (2020).

10. D. Y. Logunov, I. V. Dolzhikova, O. V. Zubkova, A. I. Tukhvatulin, D. V. Shcheblyakov, A. S. Dzharullaeva, D. M. Grousova, A. S. Erokhova, A. V. Kovyrshina, G. Botikov, F. M. Izhaeva, O. Popova, T. A. Ozharovskaya, I. B. Esmagambetov, I. A. Favorskaya, D. I. Zrelkin, D. V. Voronina, D. N. Shcherbinin, A. S. Semikhin, Y. V. Simakova, E. A. Tokarskaya, N. L. Lubenets, D. A. Egorova, M. M. Shmarov, N. A. Nikitenko, L. F. Morozova, E. A. Smolyarchuk, E. V. Kryukov, V. F. Babira, S. V. Borisevich, B. S. Naroditsky, A. L. Gintsburg, Safety and immunogenicity of an rAd26 and rAd5 vector-based heterologous prime-boost COVID-19 vaccine in two formulations: two open, non-randomised phase 1/2 studies from Russia. Lancet 396, 887–897 (2020).

11. F. C. Zhu, X. H. Guan, Y. H. Li, J. Y. Huang, T. Jiang, L. H. Hou, J. X. Li, B. F. Yang, L. Wang, W. J. Wang, S. P. Wu, Z. Wang, X. H. Wu, J. J. Xu, Z. Zhang, S. Y. Jia, B. S. Wang, Y. Hu, J. J. Liu, J. Zhang, X. A. Qian, Q. Li, H. X. Pan, H. D. Jiang, P. Deng, J. B. Gou, X. W. Wang, X. H. Wang, W. Chen, Immunogenicity and safety of a recombinant adenovirus type-5-vectored COVID-19 vaccine in healthy adults aged 18 years or older: a randomised, double-blind, placebo-controlled, phase 2 trial. Lancet 396, 479–488 (2020).

12. F. C. Zhu, Y. H. Li, X. H. Guan, L. H. Hou, W. J. Wang, J. X. Li, S. P. Wu, B. S. Wang, Z. Wang, L. Wang, S. Y. Jia, H. D. Jiang, L. Wang, T. Jiang, Y. Hu, J. B. Gou, S. B. Xu, J. J. Xu, X. W. Wang, W. Wang, W. Chen, Safety, tolerability, and immunogenicity of a recombinant adenovirus type-5 vectored COVID-19 vaccine: a dose-escalation, open-label, non-randomised, first-in-human trial. Lancet 395, 1845–1854 (2020).

13. L. A. Jackson, E. J. Anderson, N. G. Rouphael, P. C. Roberts, M. Makhene, R. N. Coler, M. P. McCullough, J. D. Chappell, M. R. Denison, L. J. Stevens, A. J. Pruijssers, A. McDermott, B. Flach, N. A. Doria-Rose, K. S. Corbett, K. M. Morabito, S. O’Dell, S. D. Schmidt, P. A. Swanson, 2nd, M. Padilla, J. R. Mascola, K. M. Neuzil, H. Bennett, W. Sun, E. Peters, M. Makowski, J. Albert, K. Cross, W. Buchanan, R. Pikaart-Tautges, J. E. Ledgerwood, B. S. Graham, J. H. Beigel, R. N. A. S. G. m, An mRNA Vaccine against SARS-CoV-2 - Preliminary Report. N Engl J Med 383, 1920–1931 (2020).

14. C. Keech, G. Albert, I. Cho, A. Robertson, P. Reed, S. Neal, J. S. Plested, M. Zhu, S. Cloney-Clark, H. Zhou, G. Smith, N. Patel, M. B. Frieman, R. E. Haupt, J. Logue, M. McGrath, S. Weston, P. A. Piedra, C. Desai, K. Callahan, M. Lewis, P. Price-Abbott, N. Formica, V. Shinde, L. Fries, J. D. Lickliter, P. Griffin, B. Wilkinson, G. M. Glenn, Phase 1-2 Trial of a SARS-CoV-2 Recombinant Spike Protein Nanoparticle Vaccine. N Engl J Med 383, 2320–2332 (2020).

15. E. J. Anderson, N. G. Rouphael, A. T. Widge, L. A. Jackson, P. C. Roberts, M. Makhene, J. D. Chappell, M. R. Denison, L. J. Stevens, A. J. Pruijssers, A. B. McDermott, B. Flach, B. C. Lin, N. A. Doria-Rose, S. O’Dell, S. D. Schmidt, K. S. Corbett, P. A. Swanson, 2nd, M. Padilla, K. M. Neuzil, H. Bennett, B. Leav, M. Makowski, J. Albert, K. Cross, V. V. Edara, K. Floyd, M. S. Suthar, D. R. Martinez, R. Baric, W. Buchanan, C. J. Luke, V. K. Phadke, C. A. Rostad, J. E. Ledgerwood, B. S. Graham, J. H. Beigel, R. N. A. S. G. m, Safety and Immunogenicity of SARS-CoV-2 mRNA-1273 Vaccine in Older Adults. N Engl J Med 383, 2427–2438 (2020).

16. E. E. Walsh, R. W. Frenck, Jr., A. R. Falsey, N. Kitchin, J. Absalon, A. Gurtman, S. Lockhart, K. Neuzil, M. J. Mulligan, R. Bailey, K. A. Swanson, P. Li, K. Koury, W. Kalina, D. Cooper, C. Fontes-Garfias, P. Y. Shi, O. Tureci, K. R. Tompkins, K. E. Lyke, V. Raabe, P. R. Dormitzer, K. U. Jansen, U. Sahin, W. C. Gruber, Safety and Immunogenicity of Two RNA-Based Covid-19 Vaccine Candidates. N Engl J Med 383, 2439–2450 (2020).

17. L. R. Baden, H. M. El Sahly, B. Essink, K. Kotloff, S. Frey, R. Novak, D. Diemert, S. A. Spector, N. Rouphael, C. B. Creech, J. McGettigan, S. Khetan, N. Segall, J. Solis, A. Brosz, C. Fierro, H. Schwartz, K. Neuzil, L. Corey, P. Gilbert, H. Janes, D. Follmann, M. Marovich, J. Mascola, L. Polakowski, J. Ledgerwood, B. S. Graham, H. Bennett, R. Pajon, C. Knightly, B. Leav, W. Deng, H. Zhou, S. Han, M. Ivarsson, J. Miller, T. Zaks, C. S. Group, Efficacy and Safety of the mRNA-1273 SARS-CoV-2 Vaccine. N Engl J Med 384, 403–416 (2021).

18. S. H. Hodgson, K. Mansatta, G. Mallett, V. Harris, K. R. W. Emary, A. J. Pollard, What defines an efficacious COVID-19 vaccine? A review of the challenges assessing the clinical efficacy of vaccines against SARS-CoV-2. Lancet Infect Dis 21, e26–e35 (2021).

19. P. Jin, J. Li, H. Pan, Y. Wu, F. Zhu, Immunological surrogate endpoints of COVID-2019 vaccines: the evidence we have versus the evidence we need. Signal Transduct Target Ther 6, 48 (2021).

20. J. H. Kim, F. Marks, J. D. Clemens, Looking beyond COVID-19 vaccine phase 3 trials. Nat Med 27, 205–211 (2021).

21. K. McMahan, J. Yu, N. B. Mercado, C. Loos, L. H. Tostanoski, A. Chandrashekar, J. Liu, L. Peter, C. Atyeo, A. Zhu, E. A. Bondzie, G. Dagotto, M. S. Gebre, C. Jacob-Dolan, Z. Li, F. Nampanya, S. Patel, L. Pessaint, A. Van Ry, K. Blade, J. Yalley-Ogunro, M. Cabus, R. Brown, A. Cook, E. Teow, H. Andersen, M. G. Lewis, D. A. Lauffenburger, G. Alter, D. H. Barouch, Correlates of protection against SARS-CoV-2 in rhesus macaques. Nature 590, 630–634 (2021).

22. Addetia, K. H. D. Crawford, A. Dingens, H. Zhu, P. Roychoudhury, M. L. Huang, K. R. Jerome, J. D. Bloom, A. L. Greninger, Neutralizing Antibodies Correlate with Protection from SARS-CoV-2 in Humans during a Fishery Vessel Outbreak with a High Attack Rate. J Clin Microbiol 58, (2020).

23. M. Voysey, S. A. Costa Clemens, S. A. Madhi, L. Y. Weckx, P. M. Folegatti, P. K. Aley, B. Angus, V. L. Baillie, S. L. Barnabas, Q. E. Bhorat, S. Bibi, C. Briner, P. Cicconi, E. A. Clutterbuck, A. M. Collins, C. L. Cutland, T. C. Darton, K. Dheda, C. Dold, C. J. A. Duncan, K. R. W. Emary, K. J. Ewer, A. Flaxman, L. Fairlie, S. N. Faust, S. Feng, D. M. Ferreira, A. Finn, E. Galiza, A. L. Goodman, C. M. Green, C. A. Green, M. Greenland, C. Hill, H. C. Hill, I. Hirsch, A. Izu, D. Jenkin, C. C. D. Joe, S. Kerridge, A. Koen, G. Kwatra, R. Lazarus, V. Libri, P. J. Lillie, N. G. Marchevsky, R. P. Marshall, A. V. A. Mendes, E. P. Milan, A. M. Minassian, A. McGregor, Y. F. Mujadidi, A. Nana, S. D. Padayachee, D. J. Phillips, A. Pittella, E. Plested, K. M. Pollock, M. N. Ramasamy, A. J. Ritchie, H. Robinson, A. V. Schwarzbold, A. Smith, R. Song, M. D. Snape, E. Sprinz, R. K. Sutherland, E. C. Thomson, M. E. Torok, M. Toshner, D. P. J. Turner, J. Vekemans, T. L. Villafana, T. White, C. J. Williams, A. D. Douglas, A. V. S. Hill, T. Lambe, S. C. Gilbert, A. J. Pollard, C. V. T. G. Oxford, Single-dose administration and the influence of the timing of the booster dose on immunogenicity and efficacy of ChAdOx1 nCoV-19 (AZD1222) vaccine: a pooled analysis of four randomised trials. Lancet 397, 881–891 (2021).

24. W. F. Garcia-Beltran, E. C. Lam, M. G. Astudillo, D. Yang, T. E. Miller, J. Feldman, B. M. Hauser, T. M. Caradonna, K. L. Clayton, A. D. Nitido, M. R. Murali, G. Alter, R. C. Charles, A. Dighe, J. A. Branda, J. K. Lennerz, D. Lingwood, A. G. Schmidt, A. J. Iafrate, A. B. Balazs, COVID-19-neutralizing antibodies predict disease severity and survival. Cell 184, 476–488 e411 (2021).

25. H. F. Sewell, R. M. Agius, D. Kendrick, M. Stewart, Covid-19 vaccines: delivering protective immunity. BMJ 371, m4838 (2020).

26. B. S. Graham, Rapid COVID-19 vaccine development. Science 368, 945–946 (2020).

27. B. F. Haynes, L. Corey, P. Fernandes, P. B. Gilbert, P. J. Hotez, S. Rao, M. R. Santos, H. Schuitemaker, M. Watson, A. Arvin, Prospects for a safe COVID-19 vaccine. Sci Transl Med 12, (2020).

28. Z. Chen, E. John Wherry, T cell responses in patients with COVID-19. Nat Rev Immunol 20, 529–536 (2020).

29. N. Vabret, G. J. Britton, C. Gruber, S. Hegde, J. Kim, M. Kuksin, R. Levantovsky, L. Malle, A. Moreira, M. D. Park, L. Pia, E. Risson, M. Saffern, B. Salome, M. Esai Selvan, M. P. Spindler, J. Tan, V. van der Heide, J. K. Gregory, K. Alexandropoulos, N. Bhardwaj, B. D. Brown, B. Greenbaum, Z. H. Gumus, D. Homann, A. Horowitz, A. O. Kamphorst, M. A. Curotto de Lafaille, S. Mehandru, M. Merad, R. M. Samstein, P. Sinai Immunology Review, Immunology of COVID-19: Current State of the Science. Immunity 52, 910–941 (2020).

30. D. Ho, P. Wang, L. Liu, S. Iketani, Y. Luo, Y. Guo, M. Wang, J. Yu, B. Zhang, P. Kwong, B. Graham, J. Mascola, J. Chang, M. Yin, M. Sobieszczyk, C. Kyratsous, L. Shapiro, Z. Sheng, M. Nair, Y. Huang, Increased Resistance of SARS-CoV-2 Variants B.1.351 and B.1.1.7 to Antibody Neutralization. Res Sq, (2021).

31. C. K. Wibmer, F. Ayres, T. Hermanus, M. Madzivhandila, P. Kgagudi, B. Oosthuysen, B. E. Lambson, T. de Oliveira, M. Vermeulen, K. van der Berg, T. Rossouw, M. Boswell, V. Ueckermann, S. Meiring, A. von Gottberg, C. Cohen, L. Morris, J. N. Bhiman, P. L. Moore, SARS-CoV-2 501Y.V2 escapes neutralization by South African COVID-19 donor plasma. Nat Med, (2021).

32. K. Wu, A. P. Werner, J. I. Moliva, M. Koch, A. Choi, G. B. E. Stewart-Jones, H. Bennett, S. Boyoglu-Barnum, W. Shi, B. S. Graham, A. Carfi, K. S. Corbett, R. A. Seder, D. K. Edwards, mRNA-1273 vaccine induces neutralizing antibodies against spike mutants from global SARS-CoV-2 variants. bioRxiv, (2021).

33. S. P. Buchbinder, D. V. Mehrotra, A. Duerr, D. W. Fitzgerald, R. Mogg, D. Li, P. B. Gilbert, J. R. Lama, M. Marmor, C. Del Rio, M. J. McElrath, D. R. Casimiro, K. M. Gottesdiener, J. A. Chodakewitz, L. Corey, M. N. Robertson, T. Step Study Protocol, Efficacy assessment of a cell-mediated immunity HIV-1 vaccine (the Step Study): a double-blind, randomised, placebo-controlled, test-of-concept trial. Lancet 372, 1881–1893 (2008).

34. S. Zhang, W. Huang, X. Zhou, Q. Zhao, Q. Wang, B. Jia, Seroprevalence of neutralizing antibodies to human adenoviruses type-5 and type-26 and chimpanzee adenovirus type-68 in healthy Chinese adults. J Med Virol 85, 1077–1084 (2013).

35. E. Zielinska, D. Liu, H. Y. Wu, J. Quiroz, R. Rappaport, D. P. Yang, Development of an improved microneutralization assay for respiratory syncytial virus by automated plaque counting using imaging analysis. Virol J 2, 84 (2005).

36. M. Aste-Amezaga, A. J. Bett, F. Wang, D. R. Casimiro, J. M. Antonello, D. K. Patel, E. C. Dell, L. L. Franlin, N. M. Dougherty, P. S. Bennett, H. C. Perry, M. E. Davies, J. W. Shiver, P. M. Keller, M. D. Yeager, Quantitative adenovirus neutralization assays based on the secreted alkaline phosphatase reporter gene: application in epidemiologic studies and in the design of adenovector vaccines. Hum Gene Ther 15, 293–304 (2004).

