## Supplemental file 1 for "GRAd-COV2, a gorilla adenovirus based candidate vaccine against COVID-19, is safe and immunogenic in young and older adults"

**Lanini S. et al Supplementary Materials**

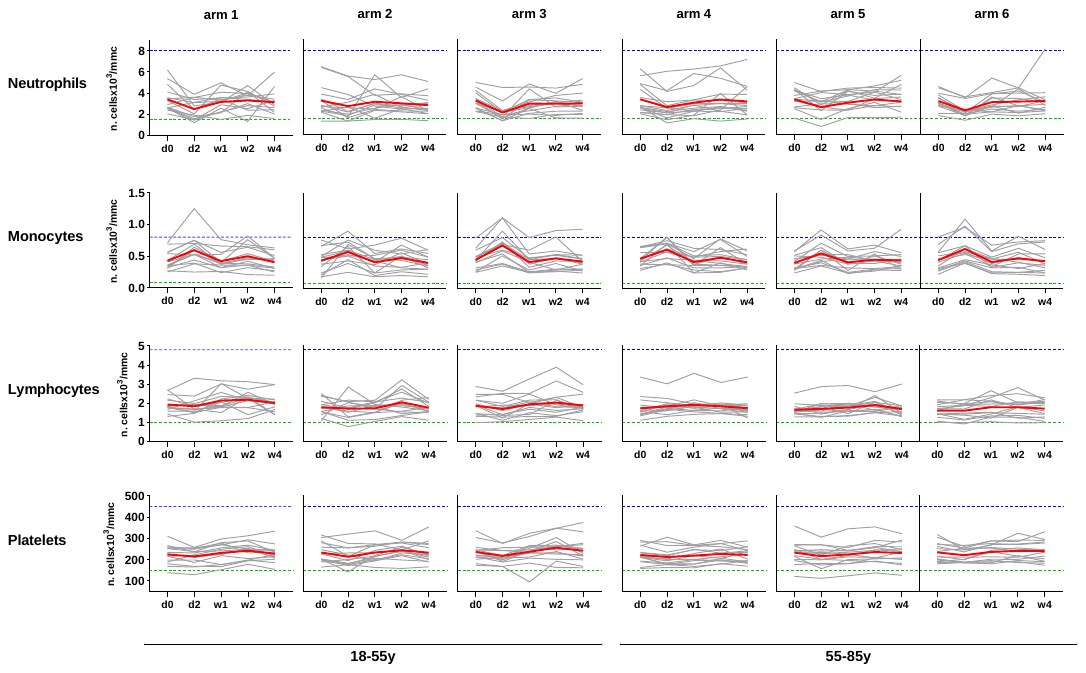

**Fig. S1**. **Hematological parameters over time.** Cell counts are shown for selected hematological parameters (Neutrophils, Monocytes, Lymphocytes and platelets) in each study arm at the following study visits: day of vaccination (d0), 48 hours after vaccination (d2), and 1, 2 and 4 weeks post vaccination (w1-w2-w4). Grey and red lines represent the cell count kinetics in individual volunteers and the group mean, respectively. Data are expressed as n. cells (x10^3^) per cubic millimeter of blood. Green and blue dashed lines denotes normal range for each parameter, namely 1.5-8 for Neutrophils, 0.08-0.8 for Monocytes, 1.5-4.8 for Lymphocytes and 150-450 for platelets.

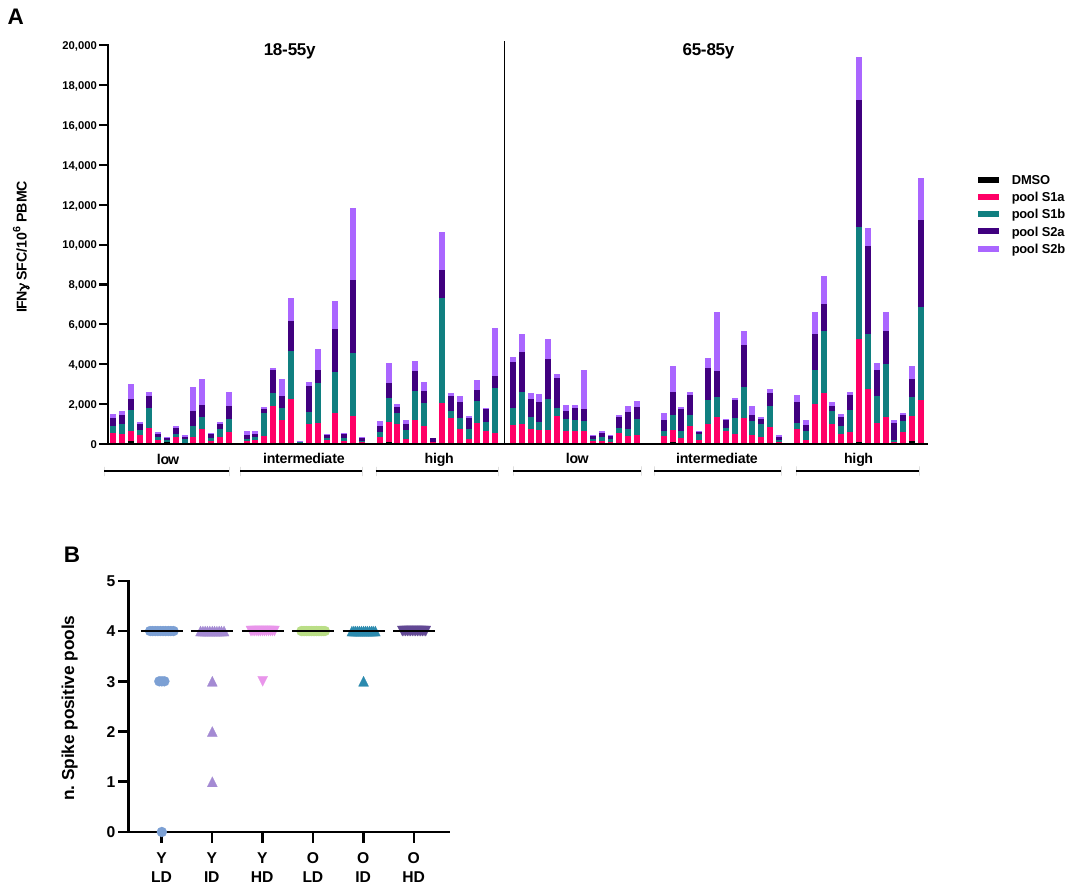

**Fig. S2. Breadth of GRAd-COV-2 induced T cell response.** A) IFNγ ELISpot response to individual S1a, S1b, S2a and S2b peptide pools and to the DMSO diluent (negative control) in freshly isolated PBMC at w2, expressed as IFNγ SFC/million PBMC. Each bar shows the response in an individual volunteer. Age cohort is indicated on top, and vaccine dose received is indicated at the bottom of the graph. B) The number of Spike peptide pools recognized in each volunteer is shown for each vaccine dose arm. Definition of a positive response is based on two criteria: response on the individual peptide pool >48 SFC/million PBMC AND response at least 3-fold the DMSO (background) response in that individual. Horizontal bar is set at median. Y=younger adults; O=older adults. LD=low dose; ID=intermediate dose; HD=high dose.

|  |  | **Y-LD** | **Y-ID** | **Y-HD** | **O-LD** | **O-ID** | **O-HD** | **HC hosp** | **HC non hosp** |
| --- | --- | --- | --- | --- | --- | --- | --- | --- | --- |
| **CLIA w4** | median | 59,45 | 60,6 | 61,8 | 31,8 | 41 | 56,3 | 78 | 24.9 |
|  | IQR | 56,1-89 | 49,1-84,4 | 27,3-87,2 | 20,8-55,3 | 21,5-67,9 | 24,2-73,8 | 56,5-111 | 16,2-49,3 |
| **ELISA**  **S** | median | 2672 | 4230 | 8120 | 2251 | 2652 | 4463 | 10216 | 2887 |
|  | IQR | 1494-6836 | 2994-9478 | 1308-11525 | 1527-4468 | 1222-7143 | 1651-11111 | 4106-32769 | 1352-4618 |
| **ELISA RBD** | median | 2490 | 3132 | 5337 | 1361 | 1478 | 3374 | 7530 | 1384 |
|  | IQR | 1166-5745 | 1925-5791 | 997-7607 | 936-2846 | 1088-5160 | 1083-5525 | 2481-33268 | 770-2459 |
| **MNA_90_** | median | <10 | 10 | 20 | 10 | 10 | 10 | 80 | 10 |
|  | IQR | <10-10 | <10-20 | <10-20 | 10-10 | <10-40 | <10-20 | 40-160 | <10-10 |
| **PNRT_50_** | median | 23,5 | 31 | 41 | 38 | 35 | 61 | na | na |
|  | IQR | 12-45,75 | 25-71 | 17-50 | 25-48 | 27-97 | 39-111 | na | na |
| **ELISpot** | median | 1162 | 2278 | 2391 | 1917 | 2073 | 3142 | na | 405 |
|  | IQR | 435-2434 | 386-5303 | 1231-3693 | 1353-3613 | 1226-3528 | 1555-8265 | na | 195-645 |

**Table S1. Median and Interquartile range of main immunological measures.** Y=younger adults; O=older adults. LD=low dose; ID=intermediate dose; HD=high dose. HC: human COVID-19 convalescent serum or PBMC

|  | A Phase 1, Dose-Escalation Study to assess the Safety and Immunogenicity of a COVID-19 Vaccine GRAd-COV2 in Healthy Adults and Elderly Subjects |
| --- | --- |
| **Short Title** | GRAd-COV2 vaccine against COVID-19 |
| **Trial ID** | RT-CoV-2 |
| **Registration number** | 2020-002835-31 |
| **Primary study Vaccine(s)** | GRAd-COV2 |
| **Version** | 3.0 final |
| **Date** | 14 December 2020 |
| **Sponsor(s)** | ReiThera Srl |
| **Manufacturer** | ReiThera Srl |
| **Principal Investigator** | Simone Lanini |
| **Conducted by** | Istituto Nazionale Malattie Infettive, Lazzaro Spallanzani |
| **Main Co-Investigators** | Stefano Milleri, Emanuele Nicastri, Andrea Antinori |
| **Version log** | Log of specific amendments by version |
| **Confidentiality statement** | Statement outlining the distribution of the document |

**SIGNATURE PAGE**

The signature below provides the necessary assurance that this trial will be conducted according to all stipulations of this protocol, including all statements regarding confidentiality, and according to local legal and regulatory requirements and applicable ICH E6 GCP guidelines.

I agree to conduct this trial in compliance with the International Conference on Harmonization Tripartite Guideline on Good Clinical Practice (E6R2) and applicable regulatory requirements.

I agree to conduct this trial in accordance with the current protocol and will not make changes to this protocol without obtaining the sponsor’s approval and National Ethical committee for COVID approval, except when necessary to protect the safety, rights, or welfare of subjects.

|  | Details | Signature |
| --- | --- | --- |
| Protocol Author/Principal Investigator | Simone Lanini  Dirigente Medico INMI Lazzaro Spallanzani IRCCS  Via Portuense 292, Roma - Italy   phone: +39 6 551701 | [DATE] |
| Investigator | Stefano Milleri  Medical and Scientific Director  Centro Ricerche Cliniche di Verona Srl  C/o Policlinico G.B. Rossi  P.le L.A. Scuro 10  37134 Verona - Italy   mobile phone: +39 045 8126618 | [DATE] |
| Sponsor | Antonella Folgori  Chief Executive Officer  ReiThera Srl  Via di Castel Romano 100  00128 Rome. Italy | [DATE] |

**Version History**

| **Protocol version N°** | **Date** | **Authors** |
| --- | --- | --- |
| 1.0 | 30 July 2020 | **Coordinating Authors: Simone Lanini**  **Contributing Authors:** Stefano Milleri, Stefania Capone, Federica Mori |
| 2.0 (Current) | 08 September 2020 | **Authors:** Simone Lanini |

**Modifications made since version 1.0**

| **Version** | **Sections** | **Modifications** |
| --- | --- | --- |
| 2.0 | Throughout | Editing, correction of mistakes and omissions |
|  | First page, footers throughout the document | Version Update |
|  | Signature page | Investigator and sponsor signatures added |
|  | 1 List of tables and Figures | List of tables and Figures has been included |
|  | Abbreviation list and throughout the document | “Participant” has been replaced by the word “Volunteer” |
|  | 5.2.2 Secondary Endpoints | Added the timepoints accidentally omitted from previous version: two timepoints (T0 and week 1) for the quantification of antibodies SAR-CoV-2 S- protein and for the detection of antibodies SAR-CoV-2 N- protein and one timepoint (T0) for the Ex vivo IFNγ ELISpot responses to SARS-CoV-2 S-protein assay. |
|  | 5.2.5 Deliverables | Deletion of fixed dates for the delivery of reports. |
|  | 7.4 Inclusion criteria | Updated of Inclusion criteria.  Inclusion criterion 5 Rephrased the sentence as a word was missing "refrain from blood DONATION"  Inclusion criteria 7 Deleted  Inclusion criterion 8 Addition of double contraceptive methods for female volunteers and criteria for male volunteers.  Inclusion criterion 11 Systolic specification removed toward Blood Pressure indication |
|  | 7.5 Exclusion criteria | Exclusion criterion 17 the volunteer must not participate in another INTERVENTIONAL study  Exclusion criteria 20 Blood dyscrasias has been deleted  Exclusion criteria 24 Added more flexible restriction for influenza and pneumococcal vaccination |
|  | 7.6 lost of follow-up and withdrawal | Updated information about lost of follow-up and withdrawal of volunteers |
|  | 7.8 Pregnancy | Updated information about pregnancy reporting and acceptable forms of contraception |
|  | 8.1 Vaccination procedure | Updated information about vaccine administration in case of any arm abnormality or permanent body art |
|  | 8.2 Medical visit, schedule of events (table 9), 8.6 Enrolment visit (T0), 8.8 Visit (D2, W1, W2, W4, W8, W12 and W24) | Updating of the information that will be collected during the study visits.  Updating of the information about diary card. |
|  | 8.3 Blood test and other analysis for monitoring safety, schedule of events (table 9) | Clarification of laboratory analysis |
|  | 8.4. Immunogenicity analysis | Updated information about HLA typing. |
|  | 8.5 Screening | Increased the duration of the screening phase. Updating information about re-screening. |
|  | 9.2 Supply and Storage | Updating information about IMP temperature excursions. |
|  | 9.3. Administration of Investigational Medicinal Products | Clarified that only one shot will be administered to each volunteer |
|  | 9.4. Accountability and Final Disposition for the Investigational Product | Included information on general process of IMP accountability and return process. |
|  | 10.1. Definitions | Updated information on Serious adverse event (SAE) and Adverse events of special interest (AESI). |
|  | 10.2. Solicited Adverse Reactions | Updated information on the classification of solicited adverse reactions. |
|  | 10.5. Reporting procedure for all AE | Updated information about reporting procedure for all AE. |
|  | 10.7. Management of volunteers with AE, Table 1 Adverse event severity scale | Updated information about AEs. |
|  | 14.1. Data safety and monitory board (DSMB) | Updated information about DSMB. |
|  | 15.1. Serology, 15.2. T cell Immunology | Updated information about exploratory studies. |
|  | 17.3. Volunteers compensation | Updated information about volunteers compensation. |
| 3.0 | 10.2. Solicited Adverse Reactions | List of Solicited Adverse Reactions has been updated based on the list of Adverse Reactions in the patient diary |

**Summary**

### 1. List of tables and Figures

### 2. Synopsis

| Trial Title | A Phase 1, Dose-Escalation Study to assess the Safety and Immunogenicity of a COVID-19 Vaccine GRAd-COV2 in Healthy Adults and Elderly Subjects | |
| --- | --- | --- |
| Internal ref. no. | RT-CoV-2 | |
| Clinical Phase | Phase I | |
| Trial Centre | Istituto per Malattie Infettive (INMI) L. Spallanzani, Via Portuense, 292, Rome; Coordinator Clinical Site  Centro Ricerche Cliniche Verona srl c/o Policlinico G.B. Rossi P.le Scuro, 10 37134 Verona | |
| Trial Design | Open label study | |
| Trial Volunteers | (i) Healthy adult volunteers (aged 18-55) including men and non-pregnant women  (ii) Elderly volunteers (aged 65-85) without significant clinical conditions | |
| Planned Sample Size | Arm 1: 15 Healthy younger adults volunteers (aged 18-55); 1 dose GRAd-COV2 (5 x 10^10^ vp)  Arm 2: 15 Healthy younger adults volunteers (aged 18-55); 1 dose GRAd-COV2 (1 x 10^11^ vp)  Arm 3: 15 Healthy younger adults volunteers (aged 18-55); 1 dose GRAd-COV2 (2 x 10^11^ vp)  Arm 4: 15 Healthy older adults volunteers (aged 65-85); 1 dose GRAd-COV2 (5 x 10^10^ vp)  Arm 5: 15 Healthy older adults volunteers (aged 65-85); 1 dose GRAd-COV2 (1 x 10^11^ vp)  Arm 6: 15 Healthy older adults volunteers (aged 65-85); 1 dose GRAd-COV2 (2 x 10^11^ vp)  Total = 90 volunteers | |
| Study duration | 12 months | |
| Follow up duration | Patients will be followed up for 24 weeks after vaccination. Follow-up will include medical visit and test at day 2, day 7, week 2, week 4 week 8, week 12 and week 24 | |
| Planned Trial Period | 10 months (from screening of first volunteer to 6 months after final vaccination of last volunteer) | |
|  | Objectives | Endpoints |
| Primary | To evaluate the safety of administering GRAd-COV2, intramuscularly in healthy (younger and older adults) volunteers | A. Occurrence of solicited local AE signs and symptoms for 7 days following the vaccination  B. Occurrence of solicited systemic AE signs and symptoms for 7 days following the vaccination  C. Occurrence of unsolicited AE for 28 days following the vaccination  D. Change from baseline for safety laboratory measures at different time point  E. Occurrence of severe AE during the whole study duration |
| Secondary | To assess the cellular and humoral immune response to SARS-CoV2 elicited by the vaccine | A. CLIA to quantify antibodies to SARS-CoV-2 S-protein (anti-S-Ab)  B. CLIA to detect antibodies to SARS-CoV-2 nucleocapsid protein (anti-N-Ab)  C. SARS-Cov-2 micro-neutralization assay to quantify neutralizing antibody activity to the virus  D. Ex vivo IFN-gamma ELISpot responses to SARS-CoV-2 S-protein |
| Investigational Medicinal Product | Replication-defective gorilla adenovirus (GRAd) encoding prefusion-stabilized SARS-CoV-2 S-protein protein, GRAd-COV2 | |
| Formulation, Dose, Route of Administration | Liquid, intramuscular needle injection | |

### 3. Abbreviation

| Ad | Adenovirus |
| --- | --- |
| AE | Adverse event |
| AR | Adverse reaction |
| ChAd | Chimpanzee Adenovirus |
| CI | Chief Investigator |
| CLIA | Chemiluminescent immunoassay |
| CMV | Cytomegalovirus |
| COVID-19 | Coronavirus disease which is caused by SARS-CoV-2 |
| CRF | Case Report Form |
| CT | Clinical Trials |
| CTA | Clinical Trials Authorisation |
| DNA | Deoxyribonucleic acid |
| DSMB | Data Safety Monitoring Board |
| DSUR | Development Safety Update Report |
| EC | Ethics Committee |
| ECG | Electrocardiogram |
| ELISpot | Enzyme-linked immunosorbent spot |
| GCP | Good Clinical Practice |
| GMO | Genetically Modified Organism |
| GP | General Practitioner (Medico di Medicina generale) |
| GRAd | Gorilla Adenovirus |
| HLA | Human Leukocyte Antigen |
| IB | Investigators Brochure |
| ICF | Informed Consent Form |
| ICH | International Conference on Harmonisation |
| IMP | Investigational Medicinal Product |
| IFN-γ | Interferon Gamma |
| N-protein | Nucleocapside protein |
| PBMC | Peripheral Blood Mononuclear Cell |
| VIL | Volunteer Information Leaflet |
| RNA | Ribonucleic acid |
| S-protein | Spike protein |
| SAE | Serious Adverse Event |
| SAR | Serious Adverse Reaction |
| SARS-CoV-2 | Severe acute respiratory syndrome coronavirus 2 |
| SFC | Spot-Forming Cells |
| SOP | Standard Operating Procedure |
| SUSAR | Suspected Unexpected Serious Adverse Reactions |
| TMF | Trial Master File |
| vp | Viral Particles |
| WHO | World Health Organisation |

### 4. Background information and rationale

#### 4.1. Coronavirus disease 2019 current situation

Severe Acute Respiratory Syndrome Coronavirus 2 (SARS-CoV-2) is associated with a new diverse clinical syndrome called Coronavirus disease 2019 (COVID-19). Common symptoms of COVID-19 include fever, cough, shortness of breath and dyspnea that may eventually progress towards acute respiratory distress syndrome and death. About 80% of patients have mild to moderate disease, 14% have severe disease and 6% are critical (namely, they develop respiratory failure, septic shock, and/or multiple organ dysfunction/failure). The severity of clinical presentation is directly dependent on patients’ age. Infections needing hospitalization are exceptional events among children younger than 10 years while the case-fatality rate can exceed 10% among people older than 70 years. [^[1]^,^[2]^]

The COVID-19 pandemic originated in Wuhan, China, in early December 2019. The World Health Organization declared the outbreak a Public Health Emergency of International Concern on January 30^th^ and by March 11^th^ it has been recognized as the cause of the biggest pandemic event over the last century. [2,^[3]^]

The first cases reported in Italy were two Chinese tourists who were identified in Rome on January 29^st^.[^[4]^] Due to the prompt intervention of Regional Infection Control network, neither of these cases did actually produce independent cluster. However, the epidemic sprung in the North of the country since the beginning of February when eleven municipalities were locked down. As for other countries, the Italian government implemented a series of strict collective measures for the control of local transmission. On March 11^th^, the government enforced a law to stop nearly all commercial activities except for supermarkets and pharmacies. [^[5]^] On March 21^st^, the quarantine was strengthened, all non-essential businesses were stopped and people’s movement severely restricted. [^[6]^] These collective measures adopted in the first phase of the epidemic, have contributed decisively to the flattening of the epidemic curve with a reduction in new cases and a consequent lightening of the health care burden. Although these measures made it possible to overcome the emergency phase, in another respect they had deleterious impact on economics and social life, which can be unsustainable in the medium-long term. Therefore, Italy, like many countries in Europe, has been easing collective measures in favor of a progressive recovery of the economic and social life. [^[7]^] However, due to the still low level of immunity of the population, a rapid resumption of sustained transmission in the community may occur. [^[8]^] There are three main strategies for controlling the spreading of new emergent pathogens during a massive event such as the current COVID-19 pandemic.

Firstly, implementing and strengthening of a robust surveillance system that it is capable of promptly identifying and of properly managing incident cases and their contacts in order to intercept local clusters and curb the subsequent epidemic waves. [^[9]^]

Secondly, endorsing research on new drugs with high direct antiviral activity that can be used either to prevent infection in high-risk individuals (pre-exposure prophylaxis), to prevent infection in exposed people (post-exposure prophylaxis) or to care infected people for reducing their capability to transmit the infection and for avoiding clinical progression toward severe diseases. [^[10]^]

Finally develop effective vaccine strategies for either preventing infection in general population or, at least, for preventing severe clinical presentation diseases in frailest subjects. [^[11]^]

Here we propose a program for rapidly scale up a new investigational vaccine based on an innovative simian adenoviral vector. The study design has been tailored on currents needs of Italy and it is aimed at rapidly demonstrating safety and immunogenicity of a single vaccine administration at escalating doses either in healthy adult or in elderly people. The results of the study will be eventually implemented into phase 2/3 clinical trial designed to meet the immediate need of those groups of people at the highest risk of severe COVID-19

#### 4.2. The emergent SARS-CoV-2 virus and the choice of S-protein protein as vaccine antigen

SARS-CoV-2 belongs to the broad family of viruses known as coronaviruses. It is a lipid enveloped virus sizing approximately 100–120 nM, and the inner nucleocapsid houses the single-stranded non-segmented positive-sense RNA genome (ssRNA+). SARS-CoV-2 is the seventh known coronavirus that can be transmitted human-to-human, after 229E, NL63, OC43, HKU1, MERS-CoV, and the original SARS-CoV. As other SARS-like coronavirus, including MERS-CoV and SARS-CoV, SARS-CoV-2 is a member of the subgenus Sarbecovirus (beta-CoV lineage B) and has been associated with severe clinical presentation in human beings. SARS-CoV-2 genome is approximately 30,000 bases in length. SARS-CoV-2 is unique among known betacoronaviruses in its incorporation of a polybasic cleavage site, a characteristic known to increase pathogenicity and transmissibility in other viruses. [^[12]^] It is believed that SARS-COV-2 has zoonotic origins due to close genetic similarity to bat coronaviruses, suggesting it emerged from a bat-borne virus. There is no evidence yet to clarify the animal-human interface, including an animal species or special environmental conditions to link the virus natural reservoir and its introduction to human communities. However, the little genetic diversity among current SARS-CoV-2 strains suggest that the spillover event might have been occurred in late 2019. [^[13]^]

Like other coronaviruses, SARS-CoV-2 has four structural proteins, known as the S (Spike), E (envelope), M (membrane), and N (nucleocapsid) proteins (Figure 1); the N-protein holds the RNA genome, and the S, E, and M proteins together create the viral envelope. Coronavirus S-protein and N-protein are highly immunogenic and constitute important antigenic targets for the development of serologic assays. [^[14]^]

The S-protein has a pivotal central role in viral infection and pathogenesis. S-protein is, in fact, the central element for allowing the virus to attach to and fuse with the membrane of a host cell; specifically, its S1 subunit catalyzes attachment, the S2 subunit fusion. [^[15]^-^[16]^]

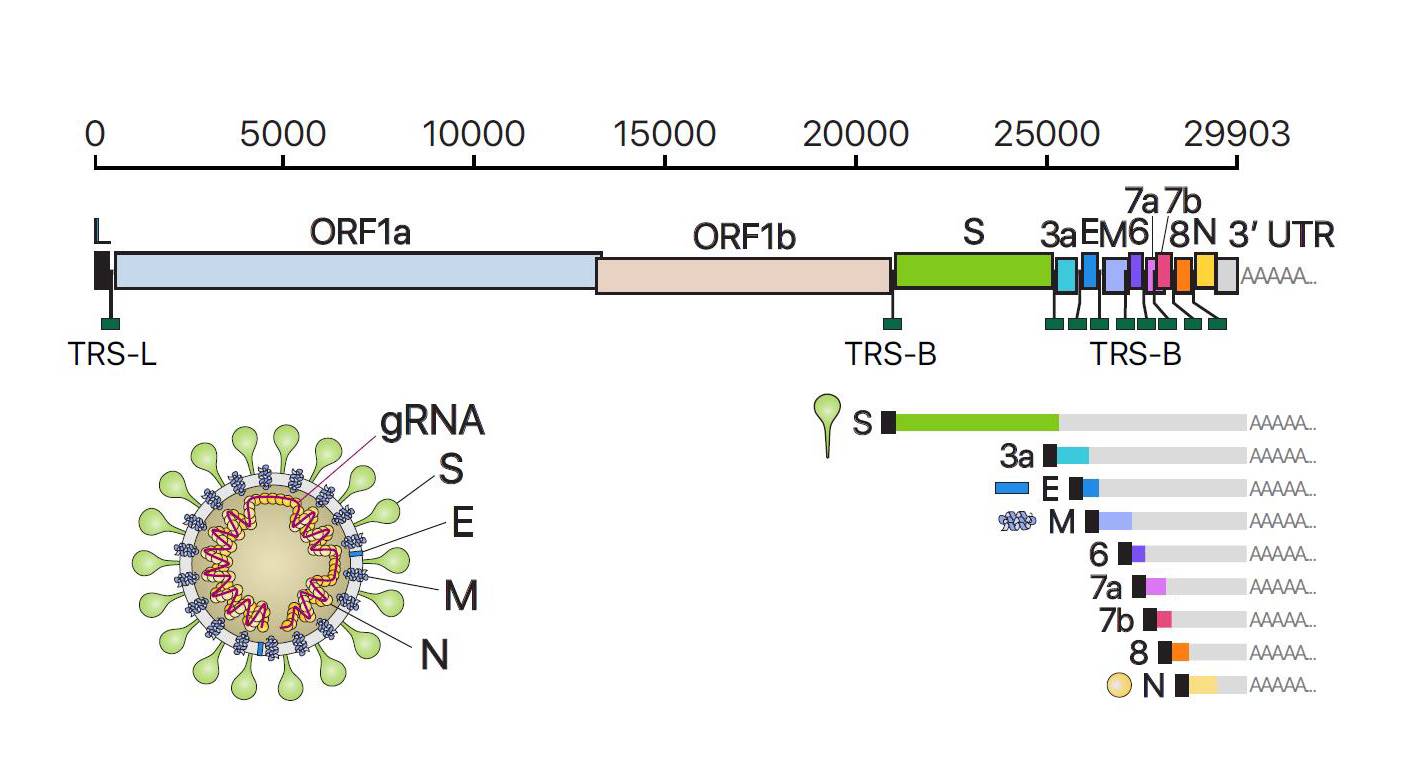

###### *Figure 1* The SARS-CoV-2

Recent evidences suggest that anti-S neutralizing antibodies from sera of COVID-19 convalescent patients can prevent cell infection in vitro by blocking viral binding to ACE2 and the eventual formation of syncytia. [^[17]^-^[18]^]

SARS-CoV-2 cell-adhesion and cell-entry is a complex mechanism. Functional S-protein has a dimeric structure including 2 subunits (S1 and S2). Cleavage of S-protein into the functional dimer is mediated by attachment of the S1 RBD to the ACE2 enzyme which exposes a cleavage site on S2 that is acted on by host cell proteases to initiate the process of cellular entry. Although SARS-CoV also uses the ACE2 receptor, SARS-CoV2 has a distinct furin cleavage site, not found in SARS-CoV, which may explain some of the biological differences. In fact, removal of this motif affects cellular entry: the furin cleavage site expands the versatility of SARS-CoV-2 for cleavage by cellular proteases and potentially the tropism and transmissibility owing to the wide cellular expression of furin proteases especially in the respiratory tract. [^[19]^-^[20]^[]](https://jcp.bmj.com/content/early/2020/05/06/jclinpath-2020-206658.long#ref-12)

The critical role of the S-protein, both in infection and pathogenesis process suggests that it is an attractive target for both for new drugs and vaccinations.

#### 4.3. Progress toward vaccination

As of June 2020, WHO reports 148 vaccines under development for SARS-CoV-2. Of those 148 candidates, 17 has already entered clinical testing while the remaining 131 are at different stage of preclinical testing. Around 10% of the candidate listed are based on “classical” vaccine approaches, such as either inactivated or live attenuated SARS-CoV-2 virus. Subunit protein in adjuvants or VLPs are well represented in the list, representing around 45% of candidates. The remaining 45% of candidate landscape is made of genetic vaccine platforms, with around one half based on either DNA or formulated RNA, and the remaining half using either replication deficient or replication competent viral vectors to deliver SARS-CoV-2 antigens. [^[21]^]

Amongst the most advanced vaccine candidates already in clinical trials, four are based on inactivated virus, three are a subunit vaccine, the other ten are genetic vaccines: two based on DNA, five on RNA and three on non-replicating adenoviral vectors.

The majority of the candidate vaccines are based solely on the SARS-CoV-2 S-protein surface glycoprotein or the corresponding gene, either in full length form or limited to the S1 subunit or to the receptor binding domain (RBD), aiming at focusing the immune response on the critical epitopes to induce neutralizing antibodies. Additional variations include the introduction of two prolines in the S2 region, to stabilize the protein in the prefusion conformation [^[22]^], or the expression of the S-protein either as a soluble protein by deleting C terminus and transmembrane domain.

Genetic platforms represent an innovative approach for implementing effective vaccine for emerging viral infections. A genetic vaccine platform based on VSV viral vector has been successfully used for developing a vaccine against Ebola virus. [^[23]^] Genetic vaccine platforms have several interesting advantages over conventional vaccines based either on whole pathogens (inactivated or live attenuated virus) or on viral components (viral proteins).

Whole pathogens vaccine is potently immunogenic, as witnessed by the fact that most of the licensed vaccines are based on this approach. However, they pose manufacturing issues mainly related to biosafety and containment. Moreover, if live-attenuated pathogens are used there is the risk of reversion to pathogenicity. The issue of reversion is particularly relevant when frail subjects are vaccinated or when novel viruses, whose biology is little known, are used. Furthermore, genetic vaccines can mimic natural interaction between the host and the pathogen much better than either inactivated or component vaccine. Indeed, in genetic vaccine the immunogenic antigens are produced intracellularly by host cell machinery. This results in Th1 polarized phenotype response. This Th1 skewed response pattern is highly desirable in vaccination against viral infection, such as SARS-CoV-2, as it is associated with clinical recovery and prevent potential concerns about vaccine-induced enhanced disease phenomena that are generally associated with Th2 response. [^[24]^-^[25]^]

In contrast genetic vaccine platforms are endowed with intrinsic safety, since they only encode for one or few antigens of the target pathogen and thus a natural infection cannot be generated by the vaccine itself. Moreover, genetic vaccine platforms are very attractive in term of programming public health preparedness and response. The development of a new genetic vaccine vector represents a framework of response against new epidemics rather than the simple vaccine strategy for a unique pathogen in a special setting. In fact, the same genetic vector can be re-designed to carry diverse antigen sequences and thus for targeting multiple pathogens. This approach is pivotal in term of public health decision as the implementation of several safe vaccine platforms may speed up the production of different vaccine tailored on the specific needs of different epidemic settings.

Adenoviral vectors are currently among the most attractive genetic vectors used for the development of vaccines.

#### 4.4. TheGRAd-COV2 vaccine

##### 4.4.1. The simian adenoviral vector vaccine platform

Replication-defective adenoviruses are among the most potent vectors for induction of antibody and T cell responses to encoded antigens. Since their discovery, adenoviruses (Ads) showed features that made them an attractive platform for vaccine design: ease of genetic manipulation, having broad tissue tropism, eliciting a robust immunogenicity and endowed with a favorable safety profile. [^[26]^] In addition, they can be easily propagated in cell culture and are relatively inexpensive to manufacture.

Ads vectors can infect many different cell types, including quiescent, long-lived cells and professional antigen presenting cells. They are not cytolitic, and their genome does not integrate in the host cell genome but remains in episomal form, conferring a safer profile compared to other viral vectors. [^[27]^] Ads genomes have been shown to persist at low levels in the target tissues. For example, upon intramuscular injection Ads genomes can be detected for months in muscles and draining lymph nodes. [^[28]^-^[29]^] These genomes have been shown to maintain a transcriptionally active form for an extended period of time [^[30]^-^[31]^-^[32]^], thus constantly re-exposing the immune system to the encoded vaccine antigen. Indeed, adenoviral vectored-vaccines have shown to induce long-lasting cellular immune responses with a large representation of T effector memory cells in the antigen-specific T cell pool, a subset that can offer immediate protection to invading pathogens. [^[33]^-^[34]^]

Adenoviral vectors can be considered self-adjuvanting vaccines. Indeed, upon infection, the viral proteins and nucleic acids act as pathogen associated molecular patterns (PAMPs) to potently trigger many extra and intracellular innate immunity sensors in the host cell, such as toll-like receptors (TLR) and other pattern recognition receptors (PRRs). The resulting innate immune response in turn can potentiate the adaptive responses to the vaccine-encoded antigens, without any need for chemical/synthetic adjuvants. [^[35]^-^[36]^] A major drawback of Ads vectored vaccine is that their efficacy can be significantly impaired by pre-existing humoral anti-vector immunity. [^[37]^] For example population immunity against Ad5, a human Ads commonly used to produce genetic vectors, range from 60% to 90% in Europe and sub-Saharan Africa, respectively [^[38]^]. The high prevalence of immunity against Ads has endorsed the use of other related viruses, such as simian adenoviruses (SAds) that have the double advantage of having very low level of sero-prevalence among human population and being associated with no human diseases. [^[39]^-^[40]^] Starting from 2007 and as of 2020, six chimpanzee-derived (three species C-ChAd3, PanAd3, ChAd155; three species E-ChAd63, ChAdOx1 and ChAdOx2) and one gorilla-derived (species C GAd20-same family of GRAd23) adenoviral vectors have been evaluated in many different clinical investigations as prophylactic and therapeutic vaccine candidates against infectious diseases or cancer. [^[41]^] Over the past 13 years, candidate vaccines based on these seven different simian vectors have been safely administered to thousands of volunteers in clinical trials up to phase 2b. The safety profile described in many studies was invariantly favorable not only in healthy adults of various geographical origin (Europe, Africa, US), but also in more vulnerable populations like elderly, children and infants, and population with concomitant healthy conditions (intravenous drug users, HCV and HIV infected patients). Of particular relevance are the results of Phase I and II clinical trials with the chimpanzee-derived adenoviral vector ChAd3 encoding Ebola virus glycoprotein (EBOV GP). These studies showed that a single dose of the vaccine induced neutralizing antibody titers comparable to those induced by the protective VSV Ebola vaccine and a better safety profile in humans. [^[42]^-^[43]^]

##### 4.4.2. The GRAd-COV2 vaccine development

The GRAd-COV2 vaccine developer, ReiThera Srl (former Okairos Srl), has pioneered the development, manufacture and clinical testing of ChAd-vectored vaccines against several infectious diseases, now owned by GlaxoSmithKline (GSK).

Independently from GSK, ReiThera has recently developed a novel proprietary, species C SAd vector, GRAd32, from a SAd strain isolated from gorilla stools. Gorilla adenovirus vaccine vectors have been described to be potently immunogenic and shown to be unfrequently recognized by neutralizing antibodies in the human population [^[44]^]. Recently, vaccine candidates based on gorilla adenovectors have been successfully assessed in animal models to protect against malaria and Zika. [^[45]^]

The GRAd32 vector is made replication defective by deleting the genomic region E1, and production is achieved by trans complementation of the E1 gene products in Hek293 cells. To further increase the cloning capacity, the E3 region is deleted from the GRAd32 genome backbone, a region also known to play a role in counteracting host innate immunity. To improve productivity, the E4 region is also deleted and the Ad5ORF6 is inserted to provide optimal interaction with the complementing Ad5 E1 region. A technological improvement in GRAd vector productivity and genetic stability is obtained by silencing transgene expression during production, thanks to constitutive expression of a tetracycline repressor by the ReiThera proprietary production cell line ReiCell35S.

The GRAd-COV2 vaccine vector contains the full length S-protein (Wuhan NCBI No QHD43416) cloned into the E1 region. The protein contains two mutations K986P, V987P which are described to stabilize the trimer in the pre-fusion conformation. [^[46]^] The rationale for the choice of a pre-fusion stabilized S-protein as a vaccine antigen is due to the fact that the wild type trimer has a metastable conformation, and the binding of the RBD to hACE2 receptor likely leads to the shedding of S1 protein from S2 protein, thus promoting S2-mediated virus-host membrane fusion and virus entry. Given the critical role of the RBD in initiating invasion of SARS-CoV-2 into host cells, it is obviously a vulnerable target for neutralizing antibodies. Stabilization of the protein in the pre-fusion conformation should avoid the dynamic of conformational states of the S-protein and provide a long window for the immunogenic epitopes of RBD exposure to specific B cells.

The C-terminus of the S-protein is fused to a peptide tag (HA tag) derived from the human influenza hemagglutinin protein, which is composed of a 9-amino acid sequence, YPYDVPDYA, that is recognized with high affinity by commercially available antibodies.

The HA tag is a commonly used epitope which does not seem to affect the activity or distribution of the expressed protein, and is useful when protein-specific antibodies are not available, as it was the case when the GRAd-COV2 vector was constructed.

#### 4.5. Pre-clinical studies on GRAd-COV2

##### 4.5.1. GRAd32 neutralizing antibodies (nAb)

The presence and levels of antibodies in human sera that are able to cross-neutralize the novel gorilla derived GRAd32 vector compared to neutralization profile of the common human Ad5 was assessed in a panel of 40 human sera, using a standard neutralization assay based on infection of HEK293 cells with Ad5 and GRAd32 vectors encoding secreted alkaline phosphatase reporter gene. [^[47]^]

Figure 2 shows that while neutralizing antibodies to Ad5 are frequently detected at high titers, cross-neutralizing nAb titers to GRAd32 are found and at significantly lower titers (geometric mean 443 vs 48). In previous clinical trials of an Ad5-based candidate vaccine for HIV, nAb titers above 200 were reported to negatively impact on immune response to vaccination. [^[48]^] Indeed, 67% of subjects in the present cohort had titers above 200 for Ad5, while only 15% of sera had cross-neutralizing activity on GRAd32.

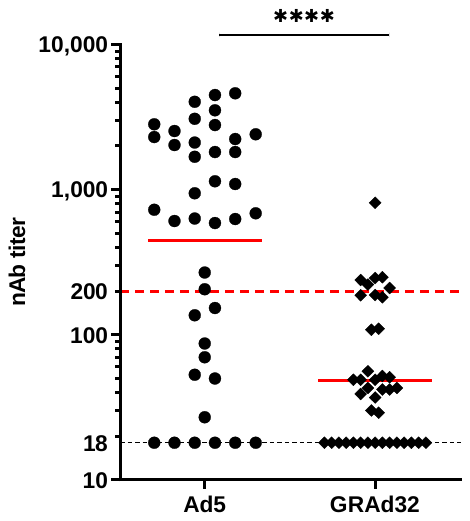

*Figure 2*. Seroprevalence of Ad5 and GRAd32 nAb in a cohort of 40 human sera. Data are expressed as the reciprocal of serum dilution able to neutralize inhibit SEAP expression by 50% compared to the SEAP expression of virus infection alone.

This feature is likely to be relevant for the potency in humans of the GRAd-COV2 vaccine, since a substantial negative impact on SARS-CoV2 humoral and cellular response to vaccination in volunteers with baseline nAb titers >200 was reported recently by CanSino in the phase I trial of their Ad5 based COVID-19 vaccine.[^[49]^] Specifically, seroconversion rate and titers of SARS-CoV-2 neutralizing antibodies and peak T cell response magnitude post vaccination were both clearly affected by Ad5 pre-existing immunity.

##### 4.5.2. GRAd-COV2 Immunogenicity in mice

Immunogenicity and potency studies were conducted in 6 to 8-weeks old female BALB/c mice.

Groups of 8 animals were vaccinated intramuscularly with a single administration of GRAd-COV2 at the dose of 10^9^ or 10^8^ viral particles (vp). Serum samples were taken 14 and 35 days post vaccination, and endpoint titers of total IgG or IgG2a/IgG1 ratio as a measure of Th1/Th2 polarization, were measured by ELISA using as a coated antigen the full length recombinant baculovirus expressed SARS-CoV-2 S-protein (Sino Biological). Neutralization titers in sera at week 5 post immunization were assessed by means of a plaque reduction assay on VERO E6 cells with SARS-CoV-2 virus.

The antigen specific T cell response in mouse splenocytes 5 weeks post immunization was characterized in depth by means of different assays. Magnitude and specificity of cellular responses were measured by IFNγ ELISpot, using as antigenic stimulus 2 pools of overlapping 15mer peptides encompassing the SARS-Cov-2 S-protein sequence: pool S1 and S2 (158 and 157 peptides each respectively, JPT PepMix). To explore the Th1/Th2 balance of the vaccine-induced response, IL4 secretion by antigen-stimulated splenocytes was also assessed by ELISpot. Finally, to characterize the antigen specific T cell subsets (CD8/CD4) and additional Th polarization profile, splenocytes were subjected to IFNγ, IL2, IL4+IL13 and IL17 intracellular staining (ICS) upon antigen restimulation and FACS analysis.

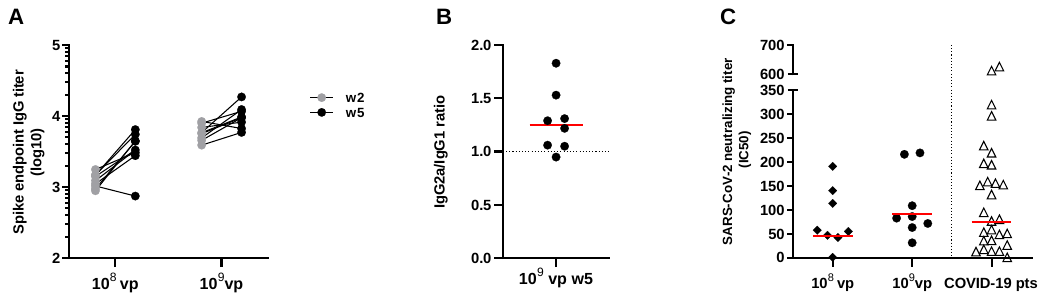

*Figure 3*. Humoral response induced in BALB/c mice at two different time points after vaccination (2 and 5 weeks) with 10^8^ and 10^9^ vp of GRAd-COV2, and comparison with neutralizing response in convalescent COVID-19 patients. A) S-protein binding total IgG titrated on individual animals by ELISA on a recombinant S-protein. Data are expressed as endpoint titer. B) IgG1 and IgG2a titers measured in week 5 sera by ELISA developed in house with appropriate secondary antibodies. Data are shown as the ratio between IgG2a and IgG1. C) SARS-CoV-2 neutralizing antibodies in week 5 mouse sera and in a panel of 28 sera from convalescent COVID-19 patients detected by SARS-CoV-2 (2019-nCoV/Italy-INMI) microneutralization assay on VERO E6 cells. Data are expressed as IC50, or the reciprocal of serum dilution achieving 50% neutralization. Horizontal lines in all graphs represent geometric mean.

A single GRAd-COV2 administration at both the tested doses rapidly induced S-protein binding antibodies in all mice 2 weeks post immunization and IgG titers increased further 5 weeks post immunization. The S-protein specific antibody response was composed of both IgG1 and IgG2a, with IgG2a/IgG1 ratio>1 indicative of Th1 polarization. Importantly, GRAd-COV2 vaccination induces SARS-CoV-2 neutralizing antibodies (Nabs) in a dose-dependent fashion, which were titrated by means of a microneutralization assays. NAbIC50 titers in the serum of GRAd-COV immunized mice were in the range of those observed in a panel of sera from convalescent COVID-19 patients.

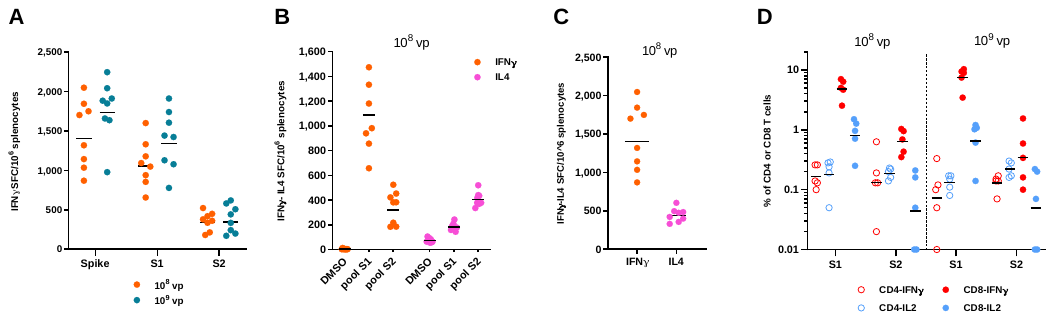

*Figure 4*. T cell response induced in BALB/c mice five weeks after vaccination with 10^8^ and 10^9^ vp of GRAd-COV2. A) IFNγ ELISpot on splenocytes. Data are expressed as IFN-γ Spot Forming Cells (SFC)/10^6^ splenocytes. Individual data points represent total S-protein response in each animal, obtained by summing reactivity to each of the 2 S-protein peptide pools and subtracting 2 times the DMSO background. Responses on S1 and S2 pools are also shown. IFNγ and IL4 ELISpot on splenocytes from mice immunized at 10^8^ vp dose. Data are shown as B) response to individual S1 and S2 pools stimulation compared to mock stimulation (DMSO) or C) as the total response to S-protein. D) IFNγ/IL2 intracellular staining and FACS analysis on splenocytes. Data are expressed as the percentage of either CD8 or CD4 secreting IFNγ or IL2 in response to S-protein peptide pools stimulation, subtracted of mock stimulation (DMSO) background. Horizontal lines in all graphs represent geometric mean.

At both the tested doses, potent T cell responses were detected in spleens of immunized mice five weeks after a single GRAd-COV2 administration (Figure 4A), a hallmark of adenoviral vectors. As also indicated by the IgG2a/IgG1 ratio, the immune response was skewed towards Th1 , as shown by a massive IFNγ secretion in response to S-protein restimulation, compared to a low level of IL4 secretion, mostly S2 targeted (Figure 4B-4C). In this mouse strain, an immunodominant CD8 epitope is located in the first S1 S-protein subunit and is responsible for the majority of IFNγ secretion reaching as high as 7.5% and 5.0% of all CD8, respectively for 10^9^ and 10^8^ dose (Figure 4D). However, additional CD8 responses in S2, as well as a clear Th1 CD4 responses in both S1 and S2, were induced at both vaccine dose tested.

IL2 secretion was also easily detected in response to S-protein stimulation in both CD8 and CD4 subsets. A small proportion (around 10-15%) of S-protein-specific IFNγ-secreting CD8 co-secreted IL2, representing most probably central memory cells precursors, with no antigen specific CD8 producing IL2 only. S-protein-specific CD4 were more heterogeneous, with either IFNγ or IL2 single-producers represented and around 60-70% of S-protein specific CD4 co-producing both cytokines (not shown). S-protein-specific Th2 and Th17 CD4 T cells were not induced, or below the limit of detection of our ICS method.

On the ground of previous studies with simian group C vectors encoding different transgenes, two low vaccine doses (10^7^ and 10^6^vp) were chosen to establish the so-called *immunological breakpoint* (Colloca et al 2012), defined as the minimal dose capable of inducing T cell responses to the encoded transgene in 40% of vaccinated animals. T cell response in splenocytes and in lung infiltrating lymphocytes (at 10^7^ vp dose only) were measured by IFNγ, IL4 and Il17 ELISpot 3 weeks post vaccination.

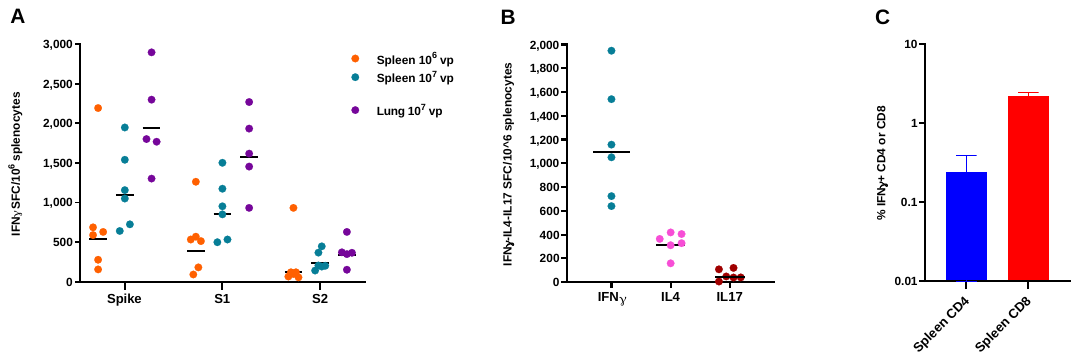

*Figure 5.* T cell response induced in BALB/c mice three weeks after vaccination with 10^7^ and 10^6^ vp of GRAd-COV2. A) IFNγ ELISpot on splenocytes and lung infiltrating lymphocytes. Data are expressed as IFN-γ Spot Forming Cells (SFC)/10^6^ splenocytes. Individual data points represent total S-protein response in each animal, obtained by summing reactivity to each of the 2 S-protein peptide pools and subtracting 2 times the DMSO background. Responses on S1 and S2 pools are also shown. B) IFNγ, IL4 and IL17 ELISpot on splenocytes from mice immunized at 10^7^ vp dose. Data are shown as the total response to S-protein. Horizontal lines in A and B graphs represent geometric mean.C) IFNγ intracellular staining and FACS analysis on splenocytes. Median+IQR is shown. Data are expressed as the percentage of either CD8 or CD4 secreting IFNγ in response to S-protein peptide pools stimulation.

Clear dose dependent T cell responses were detected by IFNγ ELISpot in the spleen of all mice at both low doses tested (10^7^ and 10^6^), indicating that GRAd32 ranks amongst the most potent group C adenoviral vectors.

Importantly, S-protein-specific T cells were found at high levels in the lungs of intramuscularly vaccinated mice three weeks post immunization (Figure 5A).

SARS-CoV-2 specific T cell response targeted epitopes in both S1 and S2 region of the protein, with same recognition hierarchy seen at higher vaccine dose. Overall, the T cell response was skewed towards Th1 also at lower vaccine dose, and while low levels of IL4 secretion were clearly detectable, S-protein-specific Th17 cells were not induced in immunized mice (Figure 5B). As expected, the majority of the IFNγ secreting S-protein-specific T cells were CD8, but lower levels of CD4 Th1 responses were also clearly detectable (Figure 5C).

Taken together the mouse data show that GRAd-COV2 candidate vaccine is highly immunogenic and induces S-protein-binding as well as SARS-CoV-2 neutralizing Ab in serum, as well as potent CD8 responses to the encoded antigen in spleen and lungs, with CD4 responses mostly indicative of Th1 polarization.

##### 4.5.3. GRAd-COV2 efficacy and safety in Rhesus macaques

An efficacy study is being conducted in collaboration with NIH/VRC in Rhesus macaques, a species shown to be susceptible to SARS-CoV-2 infection. NHPs are considered among the key animal models to determine vaccine protective efficacy and safety, in terms of assessment of vaccine-mediated enhanced disease. [^[50]^-^[51]^-^[52]^]. Indeed, upon intranasal/intratracheal SARS-COV-2 challenge the animals develop clinical symptoms and pneumonia, and virus shedding can be detected in nasal/throat swabs and in bronchoalveolar lavage (BAL).

A vaccination and SARS-CoV-2 challenge study in Rhesus macaques is being conducted in collaboration with NIH/VRC, to demonstrate vaccine immunogenicity, efficacy and safety.

The study is composed of 3 groups of 6 Rhesus macaques (macaca mulatta) each, 4-8 years old. Groups 1 and 2 received a single dose of 1x10^11^vp or 2.5x10^10^ vp GRAd-COV2 at day 0; while control Group 3 received one 1x10^11^ vp dose of a GRAd32 vector encoding a SARS-CoV-2 irrelevant antigen, as a mock vaccination.

Vaccine dosages correspond to either the intended clinical trial intermediate dose or to a lower dose for comparison with previously published data with a SARS-Cov-2 vaccine based on a different simian adenoviral vector [34].

Seven weeks post vaccination the animals will be challenged intranasally/intratracheally (1ml and 3ml volumes respectively) with ~106 PFU of SARS-CoV-2 USA/WA1/2020 strain.

Immunogenicity of the vaccine will be assessed before and after challenge in serum, PBMC and BAL cells. Infected animals will be followed for clinical, radiological (chest X-ray) and virological endpoints such as viral load by subgenomic PCR analysis and detection of infectious virus in nasal swabs and BALs, until sacrifice. At necropsy, viral load will be assessed in upper and lower respiratory tract samples, and lung pathology will be assessed by immunohistochemistry.

Study results will be available while the phase I trial is ongoing.

### 5. Objective

#### 5.1. General objective

This is an accelerated vaccine development program to be implemented during a massive global epidemic due to a newly emergent viral pathogen, SARS-CoV-2. The aim of the program is to prove *first in human* safety and immunogenicity of the new vaccine GRAd-COV2 considering both healthy adult population and special groups at risk such as the elderlies. Our approach includes a phase 1A/1B dose escalation trial with six sequential arms in two cohorts.

**Adults-Cohort.** Will enroll 45 healthy adults aged 18-55 to produce evidence on vaccine safety and immunogenicity at different vaccine doses in the general population (*first in human safety*).

**Elderlies-Cohort.** Will enroll 45 people with no major comorbidities aged 65-85 to produce evidence on vaccine safety and immunogenicity at different vaccine doses in a special population.

Our goals are to:

A. Provide preliminary information of vaccine safety and immunogenicity in healthy adults within 4 months since trial approval (Adults-Cohort)
B. Confirm vaccine safety and immunogenicity in a special population (such as older adults) within 6 months (Elderlies-Cohort)
C. Implement an eventual phase 2/3 trial tailored on current Italian epidemic situation at the beginning of 2021.
D. Provide full data about safety and immunogenicity at 6 months after vaccination within one year since the approval of the program.

Main endpoints and expected results for either Adults-Cohort or Elderlies-Cohort are reported in figure 6 and figure 7, respectively.

#### 5.2. Endpoints

##### 5.2.1. Primary endpoints (safety)

The primary endpoint of the study is to assess safety in term of occurrence of solicited and unsolicited adverse events (AE). Occurrence of AE will be monitored by preplanned medical visits and blood tests (active surveillance) and by recording volunteer’s reported AE (passive surveillance). This includes:

A. Occurrence of solicited local AE signs and symptoms for 7 days following the vaccination
B. Occurrence of solicited systemic AE signs and symptoms for 7 days following the vaccination
C. Occurrence of unsolicited AE for 28 days following the vaccination
D. Change from baseline for safety laboratory measures at 2 days, 1 week, 2 weeks, 4 week, 8 weeks, 12 weeks and 24 weeks
E. Occurrence of serious AE (SAE) during the whole study duration

Volunteers will undergo clinical follow up for AE for 24 weeks following completion of the vaccination regimen. SAEs will be collected throughout the study.

##### 5.2.2. Secondary endpoints (immunogenicity)

The secondary endpoint for the study is the assessment of cellular and humoral immune response elicited by the vaccine at different time after the administration of three different doses of vaccine, namely 5x10^10^, 1x10^11^ and 2x10^11^ viral particles, either in adults or in elderlies. This include:

A. CLIA to quantify antibodies to SARS-CoV-2 S-protein (anti-S-Ab) at T0, week 1, 2, 4, 8, 12 and 24 in adults who receive either 5x10^10^, 1x10^11^ and 2x10^11^ viral particles;
B. CLIA to quantify antibodies to SARS-CoV-2 S-protein (anti-S-Ab) at T0, week 1, 2, 4, 8, 12 and 24 in elderlies who receive either 5x10^10^, 1x10^11^ and 2x10^11^ viral particles;
C. CLIA to detect antibodies to SARS-CoV-2 N-protein(anti-N-Ab) at T0, week 1, 2, 4, 8, 12 and 24 in adults who receive either 5x10^10^, 1x10^11^ and 2x10^11^ viral particles;
D. CLIA to detect antibodies to SARS-CoV-2 N-protein(anti-N-Ab) at T0, week 1, 2, 4, 8, 12 and 24 in elderlies who receive either 5x10^10^, 1x10^11^ and 2x10^11^ viral particles;
E. SARS-Cov-2 micro-neutralization assay to quantify neutralizing antibody activity to the virus at week 4 and 24 in adults who receive either 5x10^10^, 1x10^11^ and 2x10^11^ viral particles;
F. SARS-Cov-2 micro-neutralization assay to quantify neutralizing antibody activity to the virus at week 4 and 24 in elderlies who receive either 5x10^10^, 1x10^11^ and 2x10^11^ viral particles;
G. Ex vivo IFNγ ELISpot responses to SARS-CoV-2 S-protein at T0, week 2, 4, 8, 12 and 24 in adults who receive either 5x10^10^, 1x10^11^ and 2x10^11^ viral particles
H. Ex vivo IFNγ ELISpot responses to SARS-CoV-2 S-protein at T0, week 2, 4, 8, 12 and 24 in elderlies who receive either 5x10^10^, 1x10^11^ and 2x10^11^ viral particles

##### 5.2.3. Exploratory endpoints

The exploratory objectives of this study are to provide preliminary evidence on kinetics of immune response for 3 doses (namely 5x10^10^, 1x10^11^ and 2x10^11^ viral particles) of GRAd-COV2 either in adults or in elderlies. Kinetics will be described as the variation of humoral and cellular immune response at time of enrollment and eventually at week 2, 4, 8, 12 and 24 after vaccination in each arm. Several T cells assay will be performed to identify T cell subset mainly involved in the response to vaccination and to evaluate their polyfunctionality and phenotypic profile. Level of neutralizing antibodies (nAb) to GRAd32 will be measured in serum at time of enrollment, 4 and 24 weeks after vaccination to evaluate any potential impact on vaccination efficiency. A correlation between humoral and cellular immune response will be evaluated. Within-the-cohort comparisons will be carried out to assess difference in immune response according to different doses in the same population. Across-the-cohort comparison will be carried out to assess whether elderlies and adults have different pattern of immune response after vaccination at different end points.

##### 5.2.4. Milestones

We expect to start enrollment within 4 weeks after final study approval. Eventual milestones include:

A. DSMB authorizes ARM1 group B and ARM2 Group A
B. DSMB authorizes ARM2 group B and ARM3 Group A
C. DSMB authorizes ARM3 group B
D. DSMB revises 4-week safety of whole Arm-1. DSMB authorize enrollment in ARM4 Group A.
E. DSMB authorizes ARM4 Group B and ARM5 Group A
F. DSMB authorize ARM5 Group B and ARM6 Group A
G. DSMB authorize ARM6 Group B

##### 5.2.5. Deliverables

1. A report on safety and immunogenicity at 4 weeks for Adults-Cohort will be developed; this report will include:
   a. an analysis of safety issue in the three adults-arms;
   b. pivotal considerations for authorizing enrollment in the Elderlies-Cohort;
   c. a preliminary analysis of immunogenicity in the three adults-arms aimed at identifying the most convenient dose to be used in the eventual phase 2/3 trial.
2. A report on safety and immunogenicity at 4 weeks for Elderlies-Cohort will be developed; this report will include pivotal considerations on:
   a. an analysis of safety issues in the three elderlies-arms
   b. a preliminary analysis of immunogenicity in the three elderlies-arms aimed at identifying the most convenient doses to be used in the eventual phase 2/3 trial.
   c. comparison of safety and immunogenicity between adults and elderlies (across the cohort comparison) to define the most convenient dose to be used either in general population or special groups at risk.
3. Within one year since LSLV final report will be produced including:
   a. Final analysis on safety and immunogenicity at 24 weeks after vaccination in Adults-Cohort
   b. Final analysis on safety and immunogenicity at 24 weeks after vaccination in Elderlies-Cohort

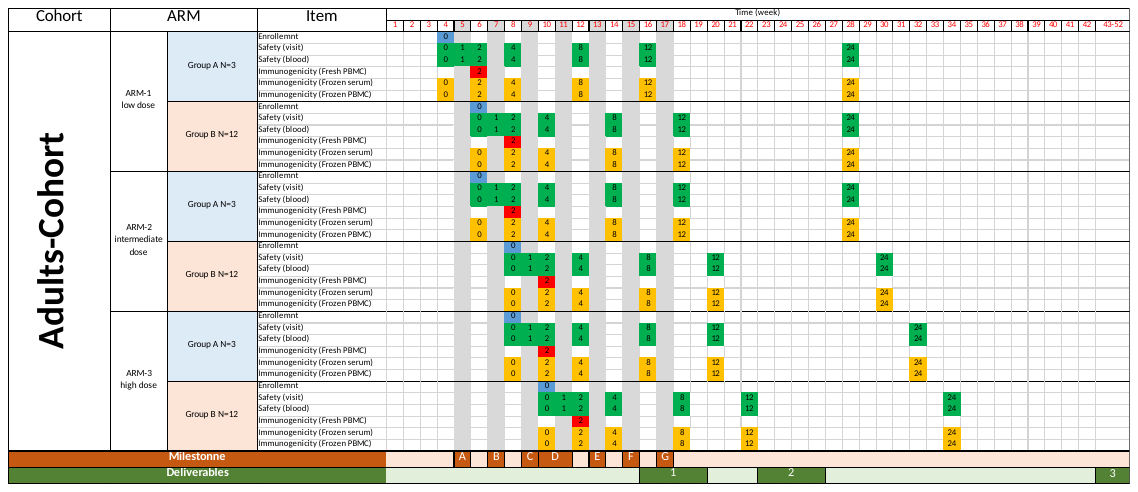

*Figure 6*. Estimated time (weeks) for enrollment and follow-up for Adults-Cohort. Figures within cells represent time (in weeks) after vaccination Grey bars represent timing of authorization for enrollment. Blue cells represent time of volunteers’ enrollment. Emerald green cells represent time for safety assessment. Golden cells represent time of sampling carried out for immunogenicity assay on frozen specimens Red cells represent time of sampling carried out for immunogenicity assay on fresh samples. Orange cells represent milestones (letters represent the specific milestone). Olive green cells represent deliverables (numbers represent the specific deliverable).

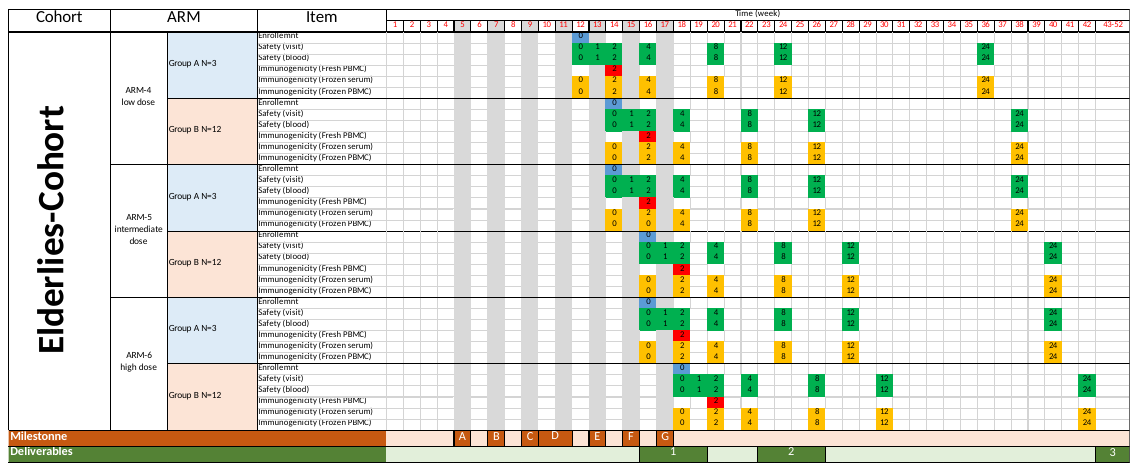

*Figure 7.* Estimated time (weeks) for enrollment and follow-up for Elderlies-Cohort. Figures within cells represent time (in weeks) after vaccination Grey bars represent timing of authorization for enrollment. Blue cells represent time of volunteers’ enrollment. Emerald green cells represent time for safety assessment. Golden cells represent time of sampling carried out for immunogenicity assay on frozen specimens Red cells represent time of sampling carried out for immunogenicity assay on fresh samples. Orange cells represent milestones (letters represent the specific milestone). Olive green cells represent deliverables (numbers represent the specific deliverable).

### 6. Study design

This is *a first-in-human*, open-label, dose escalation, phase 1A/1B clinical trial to assess the safety and immunogenicity of the candidate GRAd-COV2 vaccine in adult volunteers aged 18-55 years and elderly volunteers aged 65-85 years. The vaccine will be administered intramuscularly once in time.

The aim is to enroll 90 volunteers in two cohorts with three arms each (i.e. 15 volunteers per arm in six arms). Each study arm will assess a unique dose of the candidate GRAd-COV2 in a particular population, either adults or elderlies.

- Adults-Cohort (Phase 1A):
  - Arm-1 will investigate safety and immunogenicity of a dose of 5x10^10^ viral particles in adults.
  - Arm-2 will investigate safety and immunogenicity of a dose of 1x10^11^ viral particles in adults.
  - Arm-3 will investigate safety and immunogenicity of a dose of 2x10^11^ viral particles in adults.
- Elderlies-Cohort (Phase 1B)
  - Arm-4 will investigate safety and immunogenicity of a dose of 5x10^10^ viral particles in elderlies.
  - Arm-5 will investigate safety and immunogenicity of a dose of 1x10^11^ viral particles in elderlies.
  - Arm-6 will investigate safety and immunogenicity of a dose of 2x10^11^ viral particles in elderlies.

To minimize the risk of severe adverse events in frail subjects enrollment of Elderlies-Cohort will start after that 4-week safety results in Adults-Cohort will be available. Moreover, as it is a first in human trial, we have scheduled volunteers to be enrolled according to a strict frame of authorization rules. In particular, each arm includes two groups (A and B). Group As are made of 3 volunteers each to be enrolled throughout a week. The occurrence of SAE within 48h since the injection will be reported to data safety monitory board (DSMB) and the enrolment suspended until a direct effect of the vaccination is excluded. Group Bs are made of 12 volunteer each to be enrolled over a week. DSMB provide advice on sequential enrollment across groups and arms; this is:

- enrolment of Arm-1 group B and Arm-2 group A is bound to a favorable 7-day safety profile in Arm-1 group A;
- enrolment of Arm-2 group B and Arm-3 group-A is bound to a favorable 7-day safety profile in Arm-2 group A;
- enrolment of Arm-3 group B is bound to favorable 7-day safety profile in Arm-3 group A;
- enrolment of Arm-4 group A is bound to a favorable 4-week safety profile in the whole Arm-1;
- enrolment of Arm-4 group B and Arm-5 group A is bound to a favorable 7-day safety profile in Arm-4 group A;
- enrolment of Arm-5 group B and Arm-6 group-A is bound to a favorable 7-day safety profile in Arm-5 group A;
- enrolment of Arm-6 group B is bound to a favorable 7-day safety profile in Arm-5 group A.

Figure 8 reports the flow-charts for authorization and enrollment.

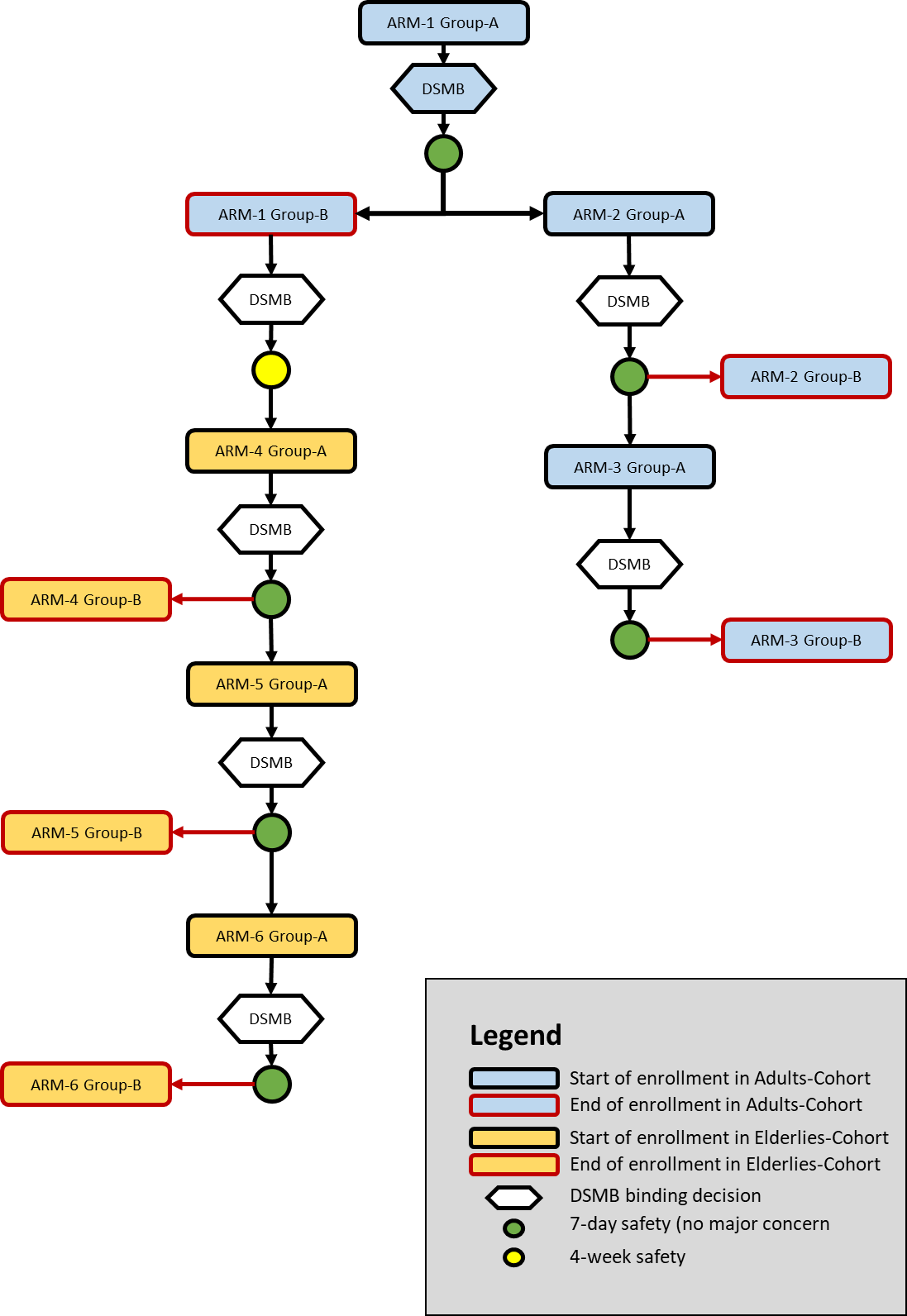

*Figure 8*. Study design and enrollment flow charts according to DSMB binding decisions.

### 7. Recruitment and withdrawal

#### 7.1. Recruitment strategy

Volunteers may be recruited by use of an advertisement +/- registration form formally approved by the ethics committee. All volunteers will sign and date the informed consent form (ICF) before any study-related procedures can be performed. Doctors enrolling the volunteers are responsible for obtaining the ICF before enrolling volunteers (i.e. before making screening test)

The volunteer’s GP (Medico di Medicina generale) will be informed and relevant clinical information registered if it is possible.

#### 7.2. Adults-Cohort

Volunteers aged 18–55 years and full details of the eligibility criteria are described in sections 6.4 and 6.5.

#### 7.3. Elderlies-Cohort

Volunteers aged 65–85 years and full details of the eligibility criteria are described below

#### 7.4. Inclusion criteria

A subject must meet all of the following criteria to be eligible to participate in this study:

1. Provides written informed consent prior to initiation of any study procedures.
2. Be able to understand and agrees to comply with planned study procedures and be available for all study visits.
3. Agrees to the collection of venous blood per protocol.
4. Confirms to have not donated blood three months before the study
5. Agrees to refrain from blood donation during the study and in the three months after the end of the study
6. Body Mass Index 18-29 kg/m^2^, inclusive, at screening.
7. Premenopausal women must have a negative urine pregnancy test the day of vaccination and are routinely using - and willing to use up to six months from vaccine administration - an effective method of birth control resulting in a low failure rate (i.e., hormonal contraception, in combination with a male barrier method). Male volunteers with female partners of childbearing potential are required to use barrier methods for the purposes of contraception whilst taking part in this study or have undergone successful vasectomy at least 6 months prior to entry into the study.
8. Oral temperature ≤37.0 degrees Celsius the day of the administration of the vaccine
9. Pulse no greater than 100 beats per minute.
10. Blood pressure (BP) is 89 to 139 mmHg, inclusive the day of vaccination.
11. Laboratory test screening are carried out no more than 21 days before vaccination and shows no clinically significant alteration.

#### 7.5. Exclusion Criteria

A subject who meets any of the following criteria will be excluded from participation in this study:

1. Positive serology for anti-HIV-Ab
2. Positive HbBsAg
3. Positive anti-HCV-Ab
4. Positive for SARS-CoV-2 (either anti-S-Ab or anti-N-Ab)
5. Acute illness, as determined by the site PI or appropriate sub-investigator, the day of vaccination.
6. Breastfeeding women
7. Autoimmune and hyper-inflammatory condition
8. History of atopy (or any [IgE](https://en.wikipedia.org/wiki/Immunoglobulin_E) associated condition) who had required treatment over the last 6 months;
9. History of hypersensitivity or severe allergic reaction (e.g., anaphylaxis, generalized urticaria, angioedema, other significant reaction) to any previous licensed or unlicensed vaccines;
10. Assumption of any immunomodulatory medication over the last 4 months (including, but not limited to, systemic corticosteroids, allergy injections, immunoglobulin, interferon, immunomodulators, cytotoxic drugs, or other similar or toxic drugs). The use of low dose topical, ophthalmic, inhaled, and intranasal steroid preparations will be permitted.
11. Presence of self-reported or medically documented significant medical condition
12. Presence of self-reported or medically documented significant psychiatric condition
13. Significant cardiovascular disease needing therapy or history of myocarditis or pericarditis or heart surgery. Volunteers in treatment with Sartans or ACE-Inhibitors and good response to therapy may be included.
14. Neurological or neurodevelopmental conditions (e.g., history of migraines in the past 5 years, epilepsy, stroke, seizures in the last 3 years, encephalopathy, focal neurologic deficits, Guillain-Barré syndrome, encephalomyelitis or transverse myelitis).
15. Ongoing malignancy or recent diagnosis of malignancy in the last five years excluding basal cell and squamous cell carcinoma of the skin, which are allowed.
16. Primary or secondary immunodeficiency of any cause.
17. Participated in another interventional study in the last 12 months.
18. Currently enrolled in or plans to participate in another clinical trial with an investigational agent that will be received during the study-reporting period.
19. Administration of immunoglobulins and/or any blood or blood products within the 4 months before the first vaccine administration or at any time during the study.
20. Has any significant disorder of coagulation.
21. Has any chronic liver disease, including fatty liver.
22. Has a history of alcohol abuse or other recreational drug use within 6 months before the first vaccine administration.
23. Has any abnormality or permanent body art (e.g., tattoo) that would interfere with the ability to observe local reactions at the injection site (deltoid region).
24. Received or plans to receive any vaccine (licensed or investigational) within 4 weeks before or after GRAd-COV2 vaccination. Influenza and Pneumococcal vaccination are permitted up to two weeks before study vaccination.
25. Has been reported as a case (confirmed or probable) of COVID-19 form the regional health system
26. Has any clinical conditions that, in the opinion of the site PI or appropriate sub-investigator, precludes study participation, this includes any acute, subacute, intermittent or chronic medical disease or condition that would place the subject at an unacceptable risk of injury, render the subject unable to meet the requirements of the protocol, or may interfere with the evaluation of responses or the subject's successful completion of this trial

#### 7.6. Lost to follow-up and withdrawal

In accordance with the principles of the current revision of the Declaration of Helsinki and any other applicable regulations, a volunteer has the right to withdraw from the study at any time and for any reason, and is not obliged to give his or her reasons for doing so. The Investigator may withdraw a volunteer at any time in the interests of the volunteer’s health and well-being.

In addition, the subject may withdraw/be withdrawn for any of the following reasons:

- Volunteer withdraws consent
- Administrative decision.
- Ineligibility (either arising during the study or retrospective, having been overlooked at screening).
- Significant protocol deviation.
- Volunteer non-compliance with study requirements.
- An adverse event which results in inability to continue to comply with study procedures.
- Loss to follow up.
- Sponsor, IEC/IRB, or regulatory agency terminates the study.

The reason for withdrawal will be recorded in the CRF (if available). If withdrawal is due to an AE, appropriate follow-up visits or medical care will be arranged, with the agreement of the volunteer, until the AE has resolved.

If a volunteer withdraws from the study, blood samples collected before their withdrawal from the trial will be used/stored unless the volunteer specifically requests otherwise. In all cases of volunteer withdrawal, except those of complete consent withdrawal, long-term safety data collection, including some procedures such as safety bloods, will continue as appropriate if subjects have received vaccine.

If subjects fail to appear for a safety follow-up assessment, extensive effort (i.e., three documented contact attempts via phone calls, e-mails, etc., made on separate occasions) will be made to locate or recall them, or at least to determine their health status. These efforts will be documented in the subject’s records and the volunteer declared lost to follow up.

The Sponsor may decide to replace a volunteer withdrawn or lost to follow-up if that is possible within the specified enrollment timeframe.

#### 7.7. Compliance with Dosing Regime

All doses in this vaccine study will be administered under the supervision of a medical investigators by registered healthcare workers and recorded in the CRF. The study medication will be at no time in the possession of the volunteer and compliance will, therefore, not be an issue.

#### 7.8. Pregnancy

Should a volunteer become pregnant during the trial, pregnancy will be reported on the Pregnancy Report form. She will be followed up as other volunteers and in addition will be followed until pregnancy outcome. We will not routinely perform venipuncture in a pregnant volunteer.

Female volunteers are required to use an effective form of contraception during the course of the study (i.e until their last follow up visit). As this is a Phase I, *first-in-human*, study there is no information about the effect of this vaccine on a fetus.

Male volunteers with female partners of childbearing potential are required to use barrier methods for the purposes of contraception from the time of signing the Information Consent Form until the end of the study (approximately 6 months)or have undergone successful vasectomy at least 6 months prior to entry into the study. Furthermore, male volunteers must avoid sperm donation.

Acceptable forms of contraception for female volunteers include:

- Established use of oral, injected or implanted hormonal methods of contraception.
- Placement of an intrauterine device (IUD) or intrauterine system (IUS).
- Total abdominal hysterectomy
- Barrier methods of contraception (condom or occlusive cap with spermicide)
- Male sterilisation, if the vasectomised partner is the sole partner for the subject.
- Same-sex-intercourse only or true abstinence, when this is in line with the preferred and usual lifestyle of the volunteers.

Periodic abstinence (e.g., calendar, ovulation, symptothermal, post-ovulation methods), declaration of abstinence for the duration of exposure to IMP, and withdrawal are not acceptable methods of contraception.

If a volunteer becomes pregnant during the study, she will be discontinued from the study immediately. Volunteers should be instructed to notify the Investigator if it is determined that they have become pregnant during the study. Whenever possible, a pregnancy should be followed to term, any premature terminations reported, and the status of the mother and child should be reported to the CRO or Sponsor after delivery.

The volunteer will be asked to read and sign a separate pregnancy follow up consent form, which would allow the study doctor to ask about the outcome of pregnancy, the birth, and health of the baby.

### 8. Clinical procedures

All volunteers will receive the GRAd-COV2 vaccine and undergo follow-up at specific endpoint for a total of 24 weeks. All follow-up visit will include a medical visit and a blood sampling. Moreover, all volunteers will receive a telephone number to be contacted for receiving assistance should any medical condition occur, potentially associated with the vaccination. Additional visits or procedures may be performed at the discretion of the investigators based on volunteers needs.

#### 8.1. Vaccination procedure

Vaccines will be administered intramuscularly in the deltoid of non-dominant arm according to different doses:

A. Dose 5x10^10^ vp (arm 1 and arm 4)
B. Dose 1x10^11^ vp (arm 2 and arm 5)
C. Dose 2x10^11^ vp (arm 3 and arm 6)

The injection site will be covered with a sterile dressing and the volunteers will stay in observation per 4 hours, in case of immediate adverse events. Investigator will decide whether the volunteer can leave or need additional time in hospital monitoring.

All volunteers will be vaccinated with the same procedure and according to the same schedule of clinical record. A standard electronic case report forms (e-CRF) will be produced and filled in.

If the volunteer has any abnormality or permanent body art (e.g., tattoo) that would interfere with the ability to observe local reactions in the deltoid of non-dominant arm, the investigator may decide to administer the vaccine in the contralateral deltoid.

#### 8.2. Medical visit

Medical visit will be carried out at the endpoints indicated in the schedule of procedures and will always include a check of medical history, adverse events, concomitant medications, physical examination, weight, height (at screening only), pulse, blood pressure, after 10 min rest in sitting position, measurement of blood oxygen saturation (SpO2%) and body temperature.

#### 8.3. Blood test and other analysis for monitoring safety

Biological testing for monitoring safety will be carried out at the endpoints indicated in the schedule of procedures at the laboratory of the clinical centers that enroll volunteers and will include:

A. Full Blood Count
B. Urine analysis
C. Biochemistry: Sodium, Potassium, Creatinine, Albumin, Liver Function Tests (ALT, AST, Bilirubin total and direct), Renal function (Creatinine and BUN), lactate dehydrogenases (LDH), alkaline phosphatases (ALP), C-reactive protein (CRP)
D. Coagulation includes: INR and aPTT ratio
E. Diagnostic serology: HBsAg, HCV antibodies, HIV antibodies (specific consent will be gained prior to testing blood for these blood-borne viruses), SARS-CoV-2

Urine will be tested for pH, Glucose, Protein (or albumin), Haemoglobin, Leukocyte esterase, Ketones, Bilirubin, Urobilinogen and Nitrites at screening. For female volunteers only, urine will be tested for beta-human chorionic gonadotrophin (β-HCG) at screening and pre-dose.

During the study, volunteers presenting with respiratory symptoms consistent with COVID-19 will undergo additional tests (molecular and serology) for SARS-CoV-2, and managed according to regional guidelines. Additional safety blood tests may be performed if clinically relevant at the discretion of the medically qualified investigators.

| Item | Time of the visit since vaccination | | | | | | | | | Type of intervention |
| --- | --- | --- | --- | --- | --- | --- | --- | --- | --- | --- |
|  | Scr | T0 | D2 | W1 | W2 | W4 | W8 | W12 | W24 |  |
| Window (days) | ≤ 28 | 0 | 0 | ±1 | ±1 | ±2 | ±4 | ±7 | ±14 |  |
| Informed consent | x |  |  |  |  |  |  |  |  | Procedures |
| Eligibility | x | x |  |  |  |  |  |  |  |  |
| Vaccination |  | x |  |  |  |  |  |  |  |  |
| AE reporting | x | x | x | x | x | x | x | x | x |  |
| Diary card |  | A |  | B | B | B^§^ |  |  |  |  |
| Checking diary card completeness |  |  | x | x | x | x |  |  |  |  |
| Medical history | x | x |  |  |  |  |  |  |  | Medical visit |
| Adverse Events | x | x | x | x | x | x | x | x | x |  |
| Concomitant Medications | x | x | x | x | x | x | x | x | x |  |
| Physical examination | x | x | x | x | x | x | x | x | x |  |
| Body Temperature | x | x | x | x | x | x | x | x | x |  |
| SpO2 % | x | x | x | x | x | x | x | x | x |  |
| Pulse | x | x | x | x | x | x | x | x | x |  |
| Blood pressure | x | x | x | x | x | x | x | x | x |  |
| ECG | x |  |  |  |  |  |  |  |  |  |
| Urine analysis | x |  |  |  |  |  |  |  |  | Urine |
| Urine B-HCG | x | x |  |  |  |  |  |  |  |  |
| Blood counts | x | x | x | x | x | x | x | x | x | Blood test |
| Biochemistry | x^†^ | x | x | x | x | x | x | x | x |  |
| Coagulation | x |  |  |  |  |  |  |  |  |  |
| HIV-Ab | x |  |  |  |  |  |  |  |  |  |
| HBsAg and HCV | x |  |  |  |  |  |  |  |  |  |
| HLA Typing |  |  |  | x |  |  |  |  |  |  |
| SARS-CoV-2 Serology * | x |  |  |  |  |  |  |  |  | Immuno |
| CLIA anti-S (IgG) |  | x |  | x | x | x | x | x | x |  |
| CLIA anti-N (IgG) |  | x |  | x | x | x | x | x | x |  |
| Micro neutralization |  |  |  |  |  | x |  |  | x |  |
| ELISpot SARS-CoV-2 |  | x |  |  | x | x | x | x | x |  |
| Total blood volume (mL) | 36 | 60 | 10 | 27 | 60 | 60 | 60 | 60 | 60 | 433 |

*Figure 9.* Timing of volunteer follow-up after vaccination. Scr=screening to be done no more than 21 days before vaccination; T0= day of the vaccination; D2= day 2 after vaccination; W1= one week after vaccination; W2= two weeks after vaccination W4= four weeks after vaccination; W8= eight weeks after vaccination; W12= twelve weeks after vaccination; W24= twenty-four weeks after vaccination. A) Diary given to volunteers; B) Diary received form volunteers. ^§^At week 4, diary cards for W3 and W4 will be collected. * This is any serological assay to detect anti-SARS-CoV-2 total IgG which is validated for clinical use in Italy (i.e. validated for diagnosis of SARS-CoV-2 infection). †no CRP testing at screening.

#### 8.4. Immunogenicity analysis

Tests for assessing immunogenicity will be carried out at INMI Lazzaro Spallanzani laboratory at the endpoints indicated in the schedule of procedures. Management of samples and methodology of testing are reported in section 14. Testing for immunogenicity includes:

A. CLIA to quantify antibodies to SARS-CoV-2 S-protein (anti-S-Ab)
B. CLIA to quantify antibodies to SARS-CoV-2 N-protein(anti-N-Ab)
C. SARS-Cov-2 micro-neutralization assay to quantify neutralizing antibody activity to the virus
D. Ex vivo IFN-gamma ELISpot responses to SARS-CoV-2 S-protein

HLA typing will be done on collected blood samples from vaccinated subjects at visit W1 to inform and enable exploratory immunogenicity assessments.

Other exploratory immunological assays including analysis polyfunctional T cell analysis and other antibody assays will be performed at the discretion of the investigators/Sponsor.

#### 8.5. Screening

Before starting the enrolment a set of subjects will undergo medical screening including a medical visit (pulse, blood pressure, SpO_2_%, body temperature, medical history, concomitant medications, physical examination, ECG) blood testing and urine analysis. Testing for screening is reported in Figure 9. Biochemical test valid for enrollment should be carry out no more than 28 days before vaccination.

Volunteers who meet selection criteria but did not receive vaccine within 28 days or did not meet 1 or more selection criteria can be re-screened only one time within 3 months from the previous screening with the approval of the Sponsor. In this case volunteer should be considered as screening failure and the reason documented in the source document and eCRF. For the re-screening volunteer needs to sign a new Informed Consent Form, new Subject ID is assigned and all screening procedures need to be completed again.

#### 8.6. Enrolment visit (Time 0)

Before vaccination, the eligibility of each volunteer will be reviewed. Investigator will check pulse, blood pressure, SpO_2_%, body temperature, adverse events, medical history, physical examination and carry out a blood sampling. Testing for time 0 is reported figure 9.

Diary Cards, a ruler and thermometer will be given to each volunteer during the enrollment visit, with instruction on use, along with the emergency 24 hour telephone number to contact on-call study physicians if needed. Diary Card cover one week of AE reporting.

The volunteer will select each day any signs and symptoms listed in the Diary and their relative intensity based on the instructions described in the Diary.

#### 8.7. Vaccination sequence

For safety reasons enrolment of *first in human* vaccine administration will be carried out on a weekly sequence in the six different arms. Each arm will be divided into 2 groups. Group-As are made of 3 volunteers each and group-Bs are made of 12 volunteers each.

At the beginning of the study clinical center will smoothly screen eligible subjects.

At enrollment on day 1 the first volunteer (arm 1 group-A) will be vaccinated ahead of any other volunteers and the profile of adverse events will be reviewed 48 hours after vaccination. Provided there are no safety concerns, as assessed by the local medical investigators, another two volunteers will be vaccinated at the same dose after at least 48 hours has elapsed following the first volunteer. Thus within one week, three volunteers will be vaccinated.

The following week the three volunteers vaccinated will undergo clinical screening for AE including a medical visit and a laboratory tests up to 7 days after vaccination. DSMB will analyze data on safety for arm 1 group-A and provide advice for the eventual enrolment. Eventual enrollment will include two blocks of volunteers this is:

A. Arm 1 group-B (12 volunteers)
B. Arm 2 group-A (3 volunteers)

Volunteers in arm 2 group-A will be scheduled for vaccination similarly to those vaccinated in arm 1 group-A, 48 hours apart from each other, provided there are no safety concerns, as assessed by the local medical investigators. Volunteers in arm 1 group-B will be smoothly vaccinated throughout the week.

The volunteers vaccinated in arm 2 group-A and arm-1 group-B undergo clinical screening for AE including a medical visit and a laboratory tests up to 7 days after vaccination. DSMB will analyze data on safety for arm 2 group-A and provide advice for the eventual enrolment. Eventual enrollment will include two blocks of volunteers this is:

A. Arm 2 group-B (12 volunteers)
B. Arm 3 group-A (3 volunteers)

Volunteers in arm 3 group-A will be scheduled for vaccination similarly to those vaccinated other group-As, 48 hours apart from each other, provided there are no safety concerns, as assessed by the local medical investigators. Volunteers in arm 2 group-B will be smoothly vaccinated throughout a week.

The volunteers vaccinated in arm 3 group-A and arm 2 group-B will undergo clinical screening for AE including a medical visit and a laboratory tests up to 7 days after vaccination. DSMB will analyze data on safety for arm 2 group-A and provide advice for the eventual enrolment. Eventual enrollment will include one block of volunteers this is:

A. Arm 3 group-B (12 volunteers)

Volunteers in arm 3 group-B will be smoothly vaccinated throughout the week.

The volunteers vaccinated in arm 3 group-B will undergo clinical screening for AE including a medical visit and a laboratory tests up to 7 days after vaccination. DSMB will analyze data and provide a general advice on vaccine safety at the conclusion of enrollment of the Adult-Cohort.

When data on 4-week safety for the whole arm 1 (15 participants) will be available. DSMB will provide advice for the eventual enrolment in the Elderlies-Cohort. The decision will be done only if no less than full data of 12 out of 15 volunteers will be available.

The enrollment of the Elderlies-Cohort will follow the same scheme of the enrollment of Adults-Cohort.

The first volunteer of the Elderly-Cohort (arm 4 group-A) will be vaccinated ahead of all other elderly volunteers. Provided there are no safety concerns, as assessed by the local medical investigators, another two volunteers will be vaccinated at the same dose after at least 48 hours has elapsed following the first volunteer. By the end of a week, three volunteers will be vaccinated.

All the volunteers vaccinated in ARM 4 group-A will undergo clinical screening for AE including a medical visit and a laboratory tests up to 7 days after vaccination. DSMB will analyze data on safety for arm 4 group-A and provide advice for the eventual enrolment. Eventual enrollment will include two blocks of volunteers this is:

A. Arm 4 group-B (12 volunteers)
B. Arm 5 group-A (3 volunteers)

Volunteers in arm 5 group-A will be scheduled for vaccination similarly to those vaccinated group-As, 48 hours apart from each other, provided there are no safety concerns, as assessed by the local medical investigators. Volunteers in arm 4 group B will be smoothly vaccinated throughout the week.

The volunteers vaccinated in arm 5 group-A and those vaccinated in arm 4 group-B will undergo clinical screening for AE including a medical visit and a laboratory tests up to 7 days after vaccination. DSMB will analyze data on safety for arm 4 group-A and provide advice for the eventual enrolment. Eventual enrollment will include two blocks of volunteers this is:

A. Arm 5 group-B (12 volunteers)
B. Arm 6 group-A (3 volunteers)

Volunteers in arm 6 group-A will be scheduled for vaccination similarly to those vaccinated group-As, 48 hours apart from each other, provided there are no safety concerns, as assessed by the local medical investigators. Volunteers in arm 5 group B will be smoothly vaccinated throughout the week.

The volunteers vaccinated in arm 5 group-A and those vaccinated in arm 5 group-B will undergo clinical screening for AE including a medical visit and a laboratory tests up to 7 days after vaccination. DSMB will analyze data on safety for arm 5 group-A and provide binding advice for the eventual enrolment. Eventual enrollment will include one block of volunteers this is:

A. Arm 6 group-B (12 volunteers)

Volunteers in arm 3 group-B will be smoothly vaccinated throughout the week.

The volunteers vaccinated in arm 6 group-B clinical screening for AE including a medical visit and a laboratory tests up to 7 days after vaccination. No more volunteers will be vaccinated.

#### 8.8. Visit (D2, W1, W2, W4, W8, W12 and W24)

Follow-up visits will take place 48h after the vaccination (D2) and eventually at week 1 (W1), 2 (W2), 4 (W4), 8 (W8), 12 (W12) and 24 (W24) after vaccination. Volunteers will be assessed for local and systemic AE, medical history, concomitant medications, pulse, blood pressure, SpO2%, body temperature, physical examination, review and blood tests at these endpoints. Blood will also be taken for exploratory immunology purposes.

Investigators will check the completeness of the diary card during the D2, W1, W2, W4 visits and will collect the completed diaries during W1, W2 and W4.

If a volunteer experience adverse events (laboratory or clinical), which the investigator (physician) determine necessary for further close observation, the volunteer may be admitted to the clinical facility of the enrolling center. Moreover, investigator can decide to perform additional medical examination if needed.

Volunteers who experience symptoms that are associated with COVID-19 including fever ≥ 37.5 °C and/or respiratory symptoms will be treated according to local guideline for suspect cases of COVD-19 until an infection with SARS-CoV-2 is not excluded by a molecular test. A test for detection of SARS-CoV-2 on respiratory sample will be carried out on all volunteers with fever ≥ 37.5 °C persisting for more than 48 hours at any time after vaccination. Volunteers with confirmed COVID-19 will be admitted to hospital until clearance of the infection.

Schedule of attendance is reported in Figure 9.

#### 8.9. Volunteers restrictions

Subjects will be asked to adhere to the following study restrictions along their study participation, which are also listed and explained in the ICF:

A. not to do strenuous physical exercise during 2 days before each visit
B. To be in fasting condition (for at least 8 h) at each visit
C. To avoid alcohol during the 24 hours before each visit

### 9. Investigational vaccine production

#### 9.1. Manufacturing and Presentation

The GRAd-COV2 vaccine consists of the replication-deficient simian adenovirus vector GRAd32, containing a stabilized surface glycoprotein (S-protein protein) antigen of the SARS-CoV2 expressed from the strong CMV IE promoter. The following vaccinations will be given in this study:

1. GRAd-COV2 5 x 10^10^ vp
2. GRAd-COV2 1 x 10^11^ vp
3. GRAd-COV2 2 x 10^11^ vp

GRAd-COV2 is manufactured in A195 Formulation Buffer at a concentration of 2 x 10^11^ vp/mL. The drug product is filled into 3mL glass vials (supplied by Nuova Ompi Stevanato Group) with a 13 mm grey chlorobutyl rubber stopper and a 13 mm aluminium seal (supplied by West Pharmaceutical Services). The filled vials are supplied sterile. The containers and closures are tested for compliance with defined specifications. The vials are made from Ph Eur borosilicate Type 1 glass.

#### 9.2. Supply and Storage

GRAd-COV2 has been formulated and vialed under Good Manufacturing Practice conditions at the ReiThera Srl Facility, Roma, IT. At ReiThera Srl the vaccine will be certified and labelled for the trial by a Qualified Person (QP) before transfer to the clinical site.

The vaccine is stored at nominal -80°C in a locked freezer, at the clinical site. All movements of the study vaccines will be documented in accordance with existing standard operating procedure (SOP). Vaccine accountability, storage, shipment and handling will be in accordance with relevant SOPs and forms.

Clear instructions will be provided to local sites for managing freezer temperature excursions.

#### 9.3. Administration of Investigational Medicinal Products

The dosages of GRAd-COV2 to be used in this study are: 5x10^10^vp (low dose), 1x10^11^vp (intermediate dose) and 2x10^11^vp (high dose). See the SOP for detailed information on the preparation, labeling, storage, and administration of the GRAd-COV2 investigational vaccine. Each volunteer will receive a single injection of a single dose of GRAd-COV2. For administration of the high dose, 1ml of GRAd-COV2 will be injected without dilution. For intermediate and low dose, the vaccine will be diluted in sterile saline solution to reach a final 1ml injection volume. Vaccine dilution will be performed by the site pharmacist on the same day of vaccine administration.

On vaccination day, GRAd-COV2 will be allowed to thaw to room temperature and will be administered within 1 hour of removal from the freezer. The vaccine will be administered intramuscularly into the deltoid of the non-dominant arm (preferably). Qualified medical personnel will wear gloves. All volunteers will be observed in the unit for 4 hours (±10 minutes) after vaccination. During administration of the investigational products, Advanced Life Support drugs and resuscitation equipment will be immediately available for the management of anaphylaxis. Vaccination will be performed and the IMPs handled according to the relevant SOPs.

The study will be performed in accordance with Genetically Modified Organisms (Contained Use) Regulations (2001/18/CE). In order to minimize dissemination of the recombinant vectored vaccine virus into the environment, inoculation sites will be covered with a dressing after immunization. This should absorb any virus that may leak out through the needle track. The dressing will be removed from the injection site after 30 minutes (+15/- 5 minutes) and will be disposed as GMO waste by autoclaving.

#### 9.4. Accountability and Final Disposition for the Investigational Product

A pharmacist at the study site will receive the study products and store them in a designated secure area. Access will be restricted to the pharmacist, nurse trained to prepare vaccine and the site monitor. The pharmacist will be responsible for keeping accurate records of the material received, the amount dispensed, and the amount remaining at the end of the study. During the study, the used and unused vials and accountability log will be monitored according to ICH-GCP. All unused vials should be returned to the Sponsor at end of Study. Used vials will be discarded of as per local site institutional guidelines for disposal of biohazardous material, and disposal documented in appropriate logs.

The on-site investigator will oversee the duties of the pharmacist and will be responsible for ensuring that the pharmacist is accountable for these responsibilities.

### 10. Adverse event assessment and management

Safety will be assessed by the frequency, incidence and nature of adverse events and severe adverse events arising during the study.

#### 10.1. Definitions

**Adverse event (AE).** (AE) is any untoward medical occurrence in a volunteer, which may occur during or after administration of the vaccine and does not necessarily have a causal relationship with the intervention. An AE can therefore be any unfavorable and unintended sign (including an abnormal laboratory finding), symptom or disease temporally associated with the study intervention, whether or not considered related to the study intervention.

**Adverse reaction (AR)**. An AR is any untoward or unintended response to the vaccine. This means that a causal relationship between the vaccine and an AE is at least a reasonable possibility, i.e., the relationship cannot be ruled out. All cases judged by the reporting medical Investigator as having a reasonable suspected causal relationship to the vaccine (i.e. possibly, probably or definitely related to the vaccine) will qualify as adverse reactions.

**Unexpected adverse reaction (UAR).** An UAR is any untoward or unintended response to the vaccine, the nature and severity of which is not consistent with the information about the medicinal product in question set out in the investigator brochure (IB) or summary of product characteristics.

**Serious adverse event (SAE).** A SAE is an AE that results in any of the following outcomes, whether or not it is considered to be related to the study intervention.

A. Death
B. Life-threatening event (i.e., the volunteer was, in the view of the Investigator, at immediate risk of death from the event that occurred).
C. Persistent or significant disability or incapacity (i.e., substantial disruption of one’s ability to carry out normal life functions).
D. Hospitalization, regardless of length of stay, even if it is a precautionary measure for continued observation.
E. Congenital anomaly/birth defect.
F. Medically significant event (examples of such medical events include allergic reaction requiring intensive treatment in an emergency room or clinic, blood dyscrasias, or convulsions regardless eventual need of hospitalization)

**Serious adverse reaction (SAR)**. A SAE (expected or unexpected) that, is in the opinion of the reporting Investigator, believed to be possibly, probably or definitely due to the vaccine.

**Suspect unexpected serious adverse reaction (SUSAR).** A severe adverse reaction, the nature and severity of which is not consistent with the information about the medicinal product in question set out in the investigator brochure (IB) or Summary of Product Characteristics.

**Adverse events of special interest (AESI)** Any grade ≥3 abnormalities on safety blood test according to Common Terminology Criteria for Adverse Events (CTCAE v.5.0 and MedDRA 23.1).

#### 10.2. Solicited Adverse Reactions

Solicited Adverse Reactions are those (foreseeable) AEs following vaccination with GRAd-COV2, and include injection site pain, erythema, swelling, myalgia, headache, fatigue, fever, chills, nausea, vomiting, diarrhea, ulceration, and abdominal pain.

Solicited Adverse Reactions are grade as following:

A. Mild: Short-lived or mild symptoms; medication may be required. No limitation to usual activity
B. Moderate: Mild to moderate limitation in usual activity. Medication may be required but hospitalization is not required.
C. Severe: Considerable limitation in activity. Invasive intervention or hospitalization is required.

We do not expect life threatening or fatal solicited adverse reactions in this study.

#### 10.3. Assessment of causality

For every AE, an assessment of the relationship of the event to the administration of the vaccine will be undertaken by the doctors who have enrolled the volunteers. An intervention-related AE refers to an AE for which there is a probable or definite relationship to administration of a vaccine. An interpretation of the causal relationship of the intervention to the AE in question will be made, based on the type of event; the relationship of the event to the time of vaccine administration; and the known biology of the vaccine. Alternative causes of the AE, such as the natural history of pre-existing medical conditions, concomitant therapy, other risk factors and the temporal relationship of the event to vaccination will be considered and investigated.

Causality assessment will take place during planned safety reviews by DSMB.

#### 10.4. Guidance for assessing relationship of AE with vaccination

A. No Relationship. No temporal relationship to vaccine and alternative etiology (clinical state, environmental or other interventions); and it does not follow known pattern of response to the vaccine.
B. Unlikely. Unlikely temporal relationship to study product and alternative etiology likely (clinical state, environmental or other interventions) and it does not follow known typical or plausible pattern of response to study product.
C. Possible. Reasonable temporal relationship to study product; or event not readily produced by clinical state, environmental or other interventions; or similar pattern of response to that seen with other vaccines
D. Probable. Reasonable temporal relationship to study product; and event not readily produced by clinical state, environment, or other interventions or known pattern of response seen with other vaccines
E. Definite. Reasonable temporal relationship to study product; and event not readily produced by clinical state, environment, or other interventions; and known pattern of response seen with other vaccines.

#### 10.5. Reporting procedure for all AE

Assessment of all local and systemic Adverse Events (AEs) will be performed throughout the study, from the time of signing of the ICF at screening through the final study visit (Week 24 or early discontinuation). All Adverse Events, whether or not attributed to study medication, will be recorded in the CRF.

All SAEs should be immediately reported (latest within 24 hours of knowledge of the event) to Parexel Safety Services by entering the SAE with all the details in the appropriate sections of the eCRF following the eCRF completion guidelines and completing the SAE report form and e-mailing/faxing the documents to Parexel Safety Services, as per details below:

France (Paris) Mailbox:

or

France (Paris) Fax numbers: +33 1 44 90 32 75 or +33 1 44 90 35 34

Upon receipt of the paper SAE form, or upon documenting an SAE via telephone call, Global Pharmacovigilance Processing Group (Parexel) will forward the SAE report source documents to the sponsor Medical Director on the same business day that the SAE form is triaged. This is required to support DSMB activities.

All AEs that result in a volunteer withdrawal from the study will be followed up until a satisfactory resolution occurs or until a non-study related causality is assigned (if the volunteer consents to this). SAEs will be collected throughout the entire trial period (i.e. until W24).

AEs will be reported to DSMB before each scheduled DSMB meeting. In special circumstance the chair of DSMB may decide to involve the ethical committee to analyze AE reports in order to define how to continue the study

#### 10.6. Assessing severity of adverse events

The severity of the adverse events will be assessed according to the scale reported below. In particular AE will be judged according to inspection of injection area, medical visit and review of all biochemical and hematological tests carried out at each endpoint. AEs autonomously reported by volunteers outside medical visit will be immediately evaluated by investigators on a telephone interview. Investigators may decide to admit volunteer to hospital for further investigation if needed.

#### 10.7. Management of volunteers with AE

The clinical center that enroll volunteers will provide care to any volunteer with any AE as appropriate for the clinical conditions.

Each enrolling center will have a medical investigator that will ensure real-time safety oversight. The investigator will review AEs deemed possibly, probably or definitely related to study interventions. The investigator will immediately register AE on the eCRF.

###### Table 1 Adverse event severity scale

| Adverse events | Severity | | |
| --- | --- | --- | --- |
|  | **Mild** | **Moderate** | **Severe** |
| Erythema | 3-50 mm | 51-100 mm | >100 mm |
| Swelling | 3-20 mm | 21-50 mm | > 50 mm |
| Ulceration | - | - | any |
| Fever | 37.5-38.0 | 38.1-39.0 | >39.0 |
| hypothermia | 35.9-35.1 | - | <35 |
| Tachycardia (bpm) | 101-115 | 116-130 | >130 |
| bradycardia (bpm) | 50-54 | 40-49 | ≤40 |
| Systolic hypertension | 140-159 | 160-179 | >179 |
| Diastolic hypertension | 91-99 | 100-109 | >109 |
| Respiratory Rate low | - | 9-11 | <9 |
| Respiratory Rate high | - | 21-24 | >24 |
| SpO2 at FiO2 21% | 94-96 | 92-93 | <92 |
| Consciousness | - | - | any |
| Biochemistry | MedDra 23.1 and CTAE 5.0. Grade 1=mild; grade2=moderate; Grade 3=severe; grade 4= life threatening; grade 5= death | | |
| Blood counts |  |  |  |

#### 10.8. Halting rules and study termination due to safety issue

Safety halting rules have been developed considering the fact that this is *a first-in-human* dose escalation study. Solicited adverse events are those listed as in section 9.2, occurring within the first 7 days after vaccination (day of vaccination and six subsequent days). Unsolicited adverse events are AE other than the solicited AEs occurring within the first 7 days, or any AEs occurring after the first 7 days after vaccination.

Enrolment of volunteers in one arm will be upheld if:

A. Solicited local adverse events (site of injection): 2 or more vaccinations in a group are followed by the same severe solicited local adverse event beginning within 3 days after vaccination (day of vaccination and two subsequent days) and persisting at severe for >48 hrs and these are deemed possibly, probably or definitely related to study interventions.
B. Solicited systemic adverse events: If 2 or more vaccinations in a group are followed by the same severe solicited systemic adverse event beginning within 2 days after vaccination (day of vaccination and one subsequent day) and these are deemed possibly, probably or definitely related to study interventions.
C. Unsolicited adverse events: If 2 or more vaccinations in a group are followed by the same serious unsolicited adverse event (including the same laboratory adverse event) and relation with vaccination cannot be immediately excluded.
D. One volunteer with serious adverse event considered possibly, probably or definitely related to vaccination occurs
E. Life-threatening condition when relation with study intervention cannot be immediately excluded.
F. Death when relation with study intervention cannot be immediately excluded.

If a halting rule has been met, Sponsor will inform the regulatory authority. Moreover, before restarting enrollment the DSMB should exclude that AE has no immediate causal relation with the vaccine or that benefit of vaccination outweigh risk of further AE occurrence. In these latter case Ethical committee should be involved in the decision. Halting rules apply to one dose and the next higher one, so that if halting rule is met for the highest dose, only this arm is upheld. If halting is met for intermediate dose, two arms are hold. If halting rule is met for the lowest dose all the study is stopped pending on the investigation of causal association between AE and vaccination (DSMB advice).

The DSMB will always consider:

A. The relationship of the AE or SAE to the vaccine.
B. The relationship of the AE or SAE to the vaccine dose, or other possible causes of the event.
C. If appropriate, additional screening or laboratory testing for other volunteers to identify those who may develop similar symptoms and alterations to the current Volunteer Information Sheet are discussed.
D. New, relevant safety information from ongoing research programs on the various components of the vaccine.

The day of vaccination volunteers should have no evidence of clinical condition that may resemble AE.

### 11. Statistical plan

#### 11.1. Sample size

This is a descriptive phase I first in human trial to assess safety and immunogenicity of three different vaccine doses. Overall, the trial will include 90 volunteers and no formal sample size calculation is carried out.

#### 11.2. Statistics analysis

This is a descriptive safety study, where volunteers will be vaccinated with different doses of GRAd-COV2 against SARS-CoV-2. Forty-five volunteers per age cohort (for a total of 90 subjects) will be vaccinated in total. This sample size should allow an estimation to be made of the frequency and magnitude of outcome measures, rather than aiming to obtain formal statistical significance for differences between groups.

Safety data will be presented according to frequency, severity and duration of adverse events. The primary analysis for immunogenicity will be to assess the difference in magnitude of SARS-CoV-2 specific T-cell and antibody responses between the groups. We will assess vaccine immunogenicity by comparing the change in these immunological parameters from baseline to different endpoints. At the end of the study, we expect to 6 measures per volunteer for T-cell response and antibody level. We will explore the possibility to describe kinetics of these parameters through non-parametric test for comparison of:

A. *within-the-arm medians* at different times (i.e. baseline vs. either week-2, week-4, week-8, week-12 and week 24) between groups at different follow-up time);
B. *within the Adults-Cohort medians* at specific time (i.e. Arm-1 vs. either Arm-2 or Arm-3 at baseline, week-2, week-4, week-8, week-12 or week-24).
C. *within the Elderlies-Cohort medians* at specific time (i.e. Arm-4 vs. either Arm-5 or Arm-6 at baseline, week-2, week-4, week-8, week-12 or week-24).
D. *Across the cohort medians at specific time* (i.e. Arm-1 vs. Arm-4; Arm-2 vs Arm-5; Arm-3 vs. Arm-6 at baseline, week-2, week-4, week-8, week-12 or week-24)

Explorative kinetics model based on regression techniques for correlated measures will be carried out if normal distribution of data is proven.

### 12. Data Management

#### 12.1. Quality Control and Source Data Verification

All subject data relating to the study will be recorded on electronic case report form (eCRF). The local investigator (physician) is responsible for verifying that data entries are accurate and correct by physically or electronically signing the eCRF. The Investigator shall assure the accuracy, completeness, legibility, and timeliness of the data reported on the eCRFs and in all required reports.

The Investigator must maintain accurate documentation (source data) that supports the information entered in the eCRF. The Investigator must permit study-related monitoring, audits, Ethical committee review, and regulatory agency inspections and provide direct access to source data documents.

Study monitors will perform ongoing source data review of data to confirm that data entered into the eCRF by authorized site personnel are accurate, complete, and verifiable from source documents; that the safety and rights of subjects are being protected and that the study is being conducted in accordance with the currently approved protocol ICH-GCP and current Italian regulation.

Records and documents, including signed ICF, pertaining to the conduct of this study must be retained by the Investigator for 15 years after study completion unless local regulations or institutional policies require a longer retention period. No records may be destroyed during the retention period without the written approval of the Sponsor. No records may be transferred to another location or party without written notification to the Sponsor.

All data generated by the site personnel will be captured electronically at each study center using eCRF. Once the eCRF clinical data have been submitted to the central server at the independent data center, corrections to the data fields will be captured in an audit trail. The reason for change, the name of the person who performed the change, together with the time and date will be logged to provide an audit trail.

If additional corrections are needed, the responsible monitor or data manager will raise a query in the eCRF. The appropriate staff at the study site will answer queries sent to the Investigator. The name of the staff member responding to the query, and time and date stamp will be captured to provide an audit trail. Once all source data verification is complete and all queries are closed, the eCRF page will be ‘frozen’ to prevent further changes.

The specific procedures to be used for data entry and query resolution using the eCRF will be provided to study sites in a training manual. In addition, site personnel will receive training on the eCRF.

#### 12.2. Audit / Inspection

The Agenzia Italiana del Farmaco the National Ethical committee for COVID and the Sponsor’s Clinical Quality Assurance Group may complete audit / inspections any time to assess compliance with the protocol and the principles of Good Clinical Practice and all other relevant regulations.

Clinical centers and principal investigator must permit study-related monitoring, audits, Ethical committee review and regulatory agency inspections and provide direct access to source data documents; they will be carried out giving due consideration to data protection and medical confidentiality. The Investigator always assures the Sponsor and designated representatives of the necessary support.

#### 12.3. Monitoring and Access to Source Data

During study conduct, monitoring will be performed by a Contract Research Organization (CRO). The CRO will conduct periodic monitoring, via telephone contacts and on-site visits (where applicable), to ensure that the protocol and guidelines are being followed.

During the on-site visits the monitors may review source documents to confirm that the data recorded on the eCRFs are accurate, with a focus on ICF and other relevant study data.

It is important that the physician(s) and their relevant personnel are available during the monitoring visits or during the phone contacts.

Checking of the eCRF entries for completeness and clarity, and cross-checking with source documents, will be required to monitor the progress of the study.

#### 12.4. Data collection

Subjects will be identified by a subject number, assigned at the screening visit.

The data will then be processed, evaluated, and stored in pseudonymised form in accordance with applicable data protection regulations.

##### 12.4.1. Data Management

The Clinical Research Organization (CRO) contracted for this study will be responsible for the study data management, which will process the data in accordance with procedures approved by the Sponsor, including data validation, resolution of errors, and quality control of defined immunogenicity analysis and safety variables.

##### 12.4.2. Case Report Form(eCRF)

The eCRFs will be used to capture all the data related to the study. The eCRFs use third party software (Rave®) to capture data via an on-line system on a computer.

Data related to the study will be recorded electronically in a central database over encrypted lines, and all entries and modifications to the data will be logged in an audit trail. Access to the system will only be granted after appropriate and documented training.

The data will be entered in the system from the source data according to the description and timelines specified in the eCRF Completion Guidelines.

Volunteers that fail screening will be noted as a screen failure in the eCRF. Screen failures are volunteers who consent to participate in the clinical study but are not subsequently entered in the study. A minimal set of screen failure information is required to ensure transparent reporting of screen failure volunteers to meet the Consolidated Standards of Reporting Trials (CONSORT) publishing requirements and to respond to queries from regulatory authorities. Minimal information includes demography, screen failure details, eligibility criteria, and any SAEs.

Electronic signatures will be used where signatures are required on pages and/or visits. Automated data entry checks will be implemented where appropriate; other data will be reviewed and evaluated for accuracy by the CRA.

All entries, corrections, and changes must be made by the investigator or a designee.

##### 12.4.3. Database Release

The database will be declared to be complete and accurate by mutual agreement between the CRO and the Sponsor. Once the database release has been approved by both parties, the database will be released in accordance with the CRO’s procedure, as agreed by the Sponsor.

If a site closes before the study has been completed, the investigator will continue to have read-only access to the eCRF until the study has been completed. At database release, all user access to the eCRF will be changed to read-only. Renewed access to the eCRF will be given if corrections or updates to the released Rave® database are required. At the end of the study, the site(s) will be provided with all data related to the site (including eCRF data, queries, and the audit trail) using a secure electronic medium; the secure storage of these data at the site is the responsibility of the investigator. When confirmation of receipt of the data has been received from all site(s), all user access to the eCRF will be revoked. If, for some reason, the data are not readable for the full retention period 15 years or in accordance with national requirements, whichever is longer), the investigator may be requested that the data be re-sent.

#### 12.5. Source Documents

Source documents provide evidence for the existence of the subject and substantiate the integrity of the data collected. Source documents are filed at the Investigator’s site.

Data entered in the eCRF that are transcribed from source documents must be consistent with the source documents or the discrepancies must be explained. The Investigator may need to request previous medical records or transfer records, depending on the study. Also, current medical records must be available.

#### 12.6. Retention of Study Documents at the Site

The investigator must keep the investigator’s set of documents in the Investigator Trial Master File (TMF) for at least 15 years after the Clinical Study Report has been approved or in accordance with national requirements, whichever is longer. The Sponsor will remind the investigator in writing of this obligation when the Clinical Study Report Synopsis is distributed to the site. Documents that belong to a hospital, institution, or private practice must be kept for the maximum period permitted by the institution.

If off-site storage is used, a study-specific binder will remain at the site after the other study specific documents have been shipped for off-site storage. This binder is considered part of the TMF and must be kept in a secure place by the site for the required period. The binder must contain, at a minimum, the following documents: a copy of the Investigator TMF Index, a certified copy of the Subject Identification Code List, and a Retrieval Form. Under no circumstances may the investigator dispose of any study documents before having obtained written consent from the Sponsor. When the required storage period has expired, the documents may be destroyed in accordance with regulations. If it becomes necessary for the Sponsor or a regulatory authority to review any documentation related to this study, the investigator must permit access to such documentation

### 13. Clinical centers

GRAd-COV2 will be tested in a FIH Phase I multi-center clinical trial.

Enrolled Clinical Sites are:

A. Istituto per Malattie Infettive (INMI) L. Spallanzani, Via Portuense, 292, Rome; Coordinator Clinical Site
B. Centro Ricerche Cliniche Verona srl c/o Policlinico G.B. Rossi P.le Scuro, 10 37134 Verona

The study will be conducted in accordance with the current approved protocol, ICH GCP, relevant regulations and standard operating procedures. Approved and relevant SOPs will be used at all clinical and laboratory sites.

### 14. Monitoring Committees

#### 14.1. Data safety and monitory board (DSMB)

DSMB will include 4 voting members and 2 technical supporting members.

The chair of the board and the 3 other voting members will be independent experts with no connection either with sponsor or clinical centers participating to the study. To participate to the board each member should disclose any potential conflict of interest according to Italian legislation.

The recommendations of DSMB are provided according to the time framework reported in section 4.2.4. These recommendations are not binding, however the enrolment of each sequential group of volunteers cannot be carried out without the advice of DMSB.

In case the enrollment is with hold because a halting rule is met, the DSMB will provide advice on how and when the study enrollment can be restarted, or the study should be terminated.

Principal investigator may seek the advice of DSMB in case of the occurrence of special situation that do not immediately meet criteria for halting rules.

Significant variation to the study design that are proposed after the approval of the study should be approved by DSMB and by ethical committee.

Further details on DSMB responsibilities and functioning are reported in the DSMB charter.

#### 14.2. Ethical committee

The Law Decree 17 March 2020 n. 18, in art. 17, indicates the ethics committee of the National Institute for Infectious Diseases Lazzaro Spallanzani (IRCCS) as the only national ethics committee for the evaluation of clinical trials on COVID-19. In this capacity, the committee will express the authorization for this study.

Local ethical committees of participating center will provide local authorization if needed according to Regional legislation.

### 15. Laboratory methods for immunogenicity assessment

#### 15.1. Serology

Specific IgG against S-protein and N-protein will be tested by using commercial CE-IVD chemiluminescence immunoassays. In particular, the anti-S IgG will recognize specifically the S1 and S2 domains of the SARS-CoV-2 virus S-protein protein (LIAISON®SARS-CoV-2 from DiaSorin®); and the anti-N specific IgG will detect recombinant N-protein from SARS-CoV-2 (Abbott Architect SARS-CoV-2 IgG assay, Abbott Diagnostics).

SARS-CoV-2 neutralizing antibodies will be detected by microneutralization assay under BSL3 conditions at INMI laboratories. Briefly, sera will be heat-inactivated, diluted 1:10 in serum-free medium, and titrated in duplicate in two-fold dilutions. Equal volumes of 100 TCID_50_/well SARS-CoV-2 (2019-nCoV/Italy-INMI) and serum dilutions will be mixed and incubated at 37°C for 30 minutes. Virus-serum mixtures will be added to sub-confluent Vero E6 cells and incubated at 37 °C and 5% CO2 for two days. Plates will be then stained with Gram’s crystal violet + 5% formaldehyde 40% m/v, washed and read at 595nm. Neutralizing antibody titers will be calculated as the last serum dilution presenting the 50% of virus growth inhibition with respect to the control virus.

GRAd32 nAb titers will be determined at ReiThera laboratories by means of a standard neutralization assay based on infection of HEK293 cells with GRAd32 vectors encoding secreted alkaline phosphatase (SEAP) reporter gene. [47] Briefly, SEAP-expressing GRAd32 pre-incubated with serial dilutions of heat-inactivated serum from trial volunteers, is added to semi-confluent HEK293 cells. SEAP activity in the cell supernatant is measured after 24 hours using a chemiluminescent substrate (CSPD). The neutralisation titre is defined as the reciprocal of sera dilution required to inhibit SEAP expression by 50% compared to the SEAP expression of virus infection alone.

Exploratory studies may include additional serological tests to characterize vaccine immunogenicity such as titration (by ELISA or other appropriate method) of Spike or RBD binding antibodies, ACE2/RBD inhibition activity or measurements of serum or plasma cytokine and chemokine profiles. Vaccine induced Spike-specific B cells may also be studied by means of ELISpot or cytofluorimetric techniques.

#### 15.2. T cell Immunology

To test the ability of vaccine to induce a T cell response, we will quantify and characterize SARS-CoV-2 specific T cells before and after vaccination by ELISpot and multiparametric flow cytometry. Peripheral Blood Mononuclear Cells (PBMC) will be isolated by density gradient centrifugation, counted and resuspended in culture medium supplemented with 10% heat inactivated FBS.

The first immunogenicity analysis will be performed after 2 weeks from vaccination (T2) using fresh isolated PBMC. Specifically, fresh PBMC isolated from peripheral blood of vaccinated subjects at T2 will be stimulated for 18-24 hours with SARS-CoV-2 peptides mix (designed on the S-protein protein) and the IFN- γ production by single cells will be evaluated by ELISpot assay. Results will be expressed as Spot Forming Cells (SFC) per million of PBMC.

Moreover, the kinetic of T cell induction after vaccination will be evaluated on frozen PBMC using different T cell assays (ELISpot assay, intracellular staining/flow cytometry and Tetramer staining) with the aim to deeply characterize the vaccine-induced T cell response overtime. Specifically, the ELISpot assay and the intracellular staining/flow cytometry will be performed after in vitro stimulation with S-protein-specific peptides and allow defining the functionality/polyfunctionality (including but not limited to IFN-γ, TNF-α, IL-17, IL-2, IL-10, CD107a) of antigen specific T cell clones. The results of the intracellular staining will be expressed as frequency of specific T cells able to exert one (single functional), two (double-functional), three (triple-functional) or four (quadruple-functional) different functions. Moreover, exploratory studies may include but are not limited to evaluation of T cell proliferative capacity, epitope mapping, Tetramer staining and flow cytometry in selected volunteers with the appropriate HLA class I and II alleles, that will allow an ex vivo deep characterization of S-protein-specific T cells to define their phenotype (activated, exhausted, effector, memory) and function.

### 16. Ethical issues

#### 16.1. Potential Risks for volunteers

The potential risk to volunteers is considered as low. The potential risks are those associated with phlebotomy and vaccination. In general, recombinant adenoviral vectors are safe. Similar vaccines encoding different antigens have been given to several thousand volunteers (including children) with a good safety profile.

##### 16.1.1. Phlebotomy

The maximum volume of blood drawn over the study period (approximately 465 mL) should not compromise these otherwise healthy volunteers. There may be minor bruising, local tenderness or pre-syncopal symptoms associated with venipuncture, which will not be documented as AEs if they occur.

##### 16.1.2. Vaccination:

GRAd-COV2 has not been used in humans before and therefore will be initially administered at the lower dose of 5 x 10^10^ vp before progressing to the higher doses of 1x10^11^ and 2x10^11^ in the arms 2,3 ,5 and 6. Potential expected risks from vaccination include local effects such as pain, redness, warmth, swelling, tenderness or itching. Systemic reactions that could potentially occur following immunization with a recombinant adenovirus vaccine include a flu-like illness with feverishness, fatigue, malaise, arthralgia, myalgia and headache.

As with any vaccine, Guillain-Barré syndrome or immune-mediated reactions that can lead to organ damage may occur, but this should be extremely rare. Serious allergic reactions including anaphylaxis could also occur and for this reason volunteers will be vaccinated in a clinical area where Advanced Life Support trained physicians, equipment and drugs are immediately available for the management of any serious adverse reactions (SAR).

##### 16.1.3. Vaccine induced Disease Enhancement

A potential risk exists that the immunity elicited by an investigational candidate vaccine for COVID-19 may provoke an enhancement of the disease upon infection with SARS-CoV-2. This phenomenon has been described only in animal models when testing vaccines against other coronaviruses, and it was shown to be due to vaccine-induced “suboptimal” immune responses, such as poorly neutralizing antibodies and Th2 skewed immune response promoting inflammatory and allergic immune environment upon pathogen encounter.

The vaccine-mediated disease enhancement has not been observed so far in animal models of SARS-CoV-2 infection or vaccination. In addition, the GRAd-COV2 vaccine has been designed to minimize this risk: first, the choice of a prefusion-stabilized SARS-Cov-2 S-protein protein is meant to maximize the induction of properly neutralizing antibodies, and second, the adenoviral vectored vaccine is a potent inducers of Th1 skewed immune responses that are known to prevent or blunt inflammatory and allergic immune responses.

Nevertheless, as recommended by the International Coalition of Medicines Regulatory Authorities (ICMRA) in a meeting held on March 18 2020 (Global regulatory workshop on COVID-19 vaccine development, http://www.icmra.info/drupal/news/March2020/summary), this potential risk has been clearly stated in the ICF.

During the study follow up, as an additional safety measure, extra-visits are foreseen in case any subject develops febrile or respiratory symptoms. If the investigator will consider these symptoms compatible with COVID-19, the subject will be assessed for the presence of SARS-CoV-2 infection in respiratory tract samples. This measure is meant to ensure that potential exposure to SARS-CoV-2 in vaccinated subjects will be properly monitored and scored to assess the occurrence of any vaccine-mediated disease enhancement.

#### 16.2.  Known Potential Benefits

Volunteers will not benefit directly from participation in this study. However, it is hoped that the information gained from this study will contribute to the development of a safe and effective SARS-CoV-2 vaccine regime. The only benefits for volunteers would be information about their general health status.

#### 16.3. Declaration of Helsinki

The Investigators will ensure that this study is conducted according to the principles of the current revision of the Declaration of Helsinki.

#### 16.4. Guidelines for Good Clinical Practice

The Investigators will ensure that this study is conducted in full conformity with the Good Clinical Practice E2R6 .

#### 16.5. Approvals

The protocol, ICF, volunteer information sheet and any proposed advertising material will be submitted to an appropriate Ethics Committee and Italian regulatory authorities (ISS and AIFA).

The Investigator will submit and, where necessary, obtain approval from the above parties for all substantial amendments to the original approved documents.

No substantial amendments to this protocol will be made without consultation with, and agreement of, the Sponsor and Ethics Committee. Any substantial amendments to the trial that appear necessary during the course of the trial must be discussed by the Investigator and Sponsor concurrently. If agreement is reached concerning the need for an amendment, it will be produced in writing by the Principal Investigator and will be made a formal part of the protocol following ethical and regulatory approval.

#### 16.6.  Informed consent and volunteer confidentiality

All volunteers will sign and date the informed consent form before any study specific procedures are performed. At the screening visit, the volunteer will be fully informed of all aspects of the trial, the potential risks and the time to be spent for attending follow-up visit and the restriction to personal activities. The following general principles will be emphasized:

A. Participation in the study is entirely voluntary
B. Refusal to participate involves no penalty or loss of medical benefits
C. The volunteer may withdraw from the study at any time
D. The volunteers free to ask questions at any time to allow him or her to understand the purpose of the study and the procedures involved
E. The study involves research of an investigational vaccine
F. The study involves genetic analysis for characterization of HLA type
G. There is no direct benefit from participating
H. The volunteer’s GP will be contacted to corroborate their medical history
I. The volunteer’s blood samples taken as part of the study will be stored indefinitely and samples may be sent outside of the Italy and Europe to laboratories in collaboration with the INMI Lazzaro Spallanzani. These will be anonymized.
J. The aims of the study and all tests to be carried out will be explained.

The volunteer will be given the opportunity to ask other questions about the trial, and will then have time to consider whether or not to participate. If they do decide to participate, they will sign and date two copies of the consent form (paper document), one for them to take away and keep, and one to be stored at clinical center.

### 17. Financing and Insurance

#### 17.1. Financing

The clinical trial is funded by ReiThera SRL and IRCCS INMI Spallanzani. IRCCS INMI Spallanzani owns the property of the IMP lot #RL20-0024.

#### 17.2. Insurance

Indemnity and/or compensation for harm arising specifically from an accidental injury and occurring as a consequence of the research volunteer participation in the trial for which ReiThera is the research Sponsor will be covered by a no-fault compensation policy with HDI Global SE – DPT. SPECIAL LINES.

#### 17.3. Volunteers compensation

Volunteers will be compensated for their time and for the inconvenience caused by procedures. Volunteers you will receive an indemnity of €700/00 (seven hundred/00 euros). This amount is inclusive of all expenses that volunteer may have to incur and it is in compliance with art. 1 c. 5 of Legislative Decree 211/2003. Payment of this amount will be made within 60 days of the end of the study.

### 18. Expected results and publication policy

Publication of the results of the trial will be agreed between ReiThera SRL and IRCCS INMI Spallanzani. If one of the parties is interested in proceeding with a scientific publication that covers this clinical trial as a whole or in part, it must give written communication, also electronically, to the other party involved. In order to ensure that any patent protection opportunities or other interests, including national ones, are not affected. Any impediments to publication must also be communicated electronically within 30 days of receipt of the communication of publication of the interested party. If a party deems it inappropriate to publish, it will still try to provide written changes and / or additions to the text that could allow its publication, albeit in a reduced version. All publications covering this clinical trial as a whole or in part must report the affiliation of all the parties involved and the recognition of the authors involved. The Authors’ list will be defined on the basis of the principles contained in the document Uniform Requirements for Manuscripts submitted by Biomedical Journals.

### 19. Reference

1. European Centre for Disease Prevention and Control. Outbreak of novel coronavirus disease 2019 (COVID-19): increased transmission globally – fifth update, 2 March 2020. ECDC: Stockholm; 2020
2. Jin Y, Yang H, Ji W, Wu W, Chen S, Zhang W, Duan G. Virology, Epidemiology, Pathogenesis, and Control of COVID-19. Viruses. 2020 Mar 27;12(4):372. doi: 10.3390/v12040372. PMID: 32230900; PMCID: PMC7232198.
3. https://www.who.int/emergencies/diseases/novel-coronav
4. Albarello F, Pianura E, Di Stefano F, et al. 2019-novel Coronavirus severe adult respiratory distress syndrome in two cases in Italy: An uncommon radiological presentation. Int J Infect Dis. 2020;93:192‐197. doi:10.1016/j.ijid.2020.02.043
5. Lazzerini M, Putoto G. COVID-19 in Italy: momentous decisions and many uncertainties. Lancet Glob Health. 2020;8(5):e641‐e642. doi:10.1016/S2214-109X(20)30110-8
6. http://www.salute.gov.it/portale/nuovocoronavirus/dettaglioNotizieNuovoCoronavirus.jsp?lingua=italiano&menu=notizie&p=dalministero&id=4829
7. European Centre for Disease Prevention and Control. Projected baselines of COVID-19 in the EU/EEA and the UK for assessing the impact of de-escalation of measures – 26 May 2020. ECDC: Stockholm; 2020
8. Clemente-Suárez VJ, Hormeño-Holgado A, Jiménez M, Benitez-Agudelo JC, Navarro-Jiménez E, Perez-Palencia N, Maestre-Serrano R, Laborde-Cárdenas CC, Tornero-Aguilera JF. Dynamics of Population Immunity Due to the Herd Effect in the COVID-19 Pandemic. Vaccines (Basel). 2020 May 19;8(2):E236. doi: 10.3390/vaccines8020236. PMID: 32438622.
9. McMichael TM, Currie DW, Clark S, Pogosjans S, Kay M, Schwartz NG, Lewis J, Baer A, Kawakami V, Lukoff MD, Ferro J, Brostrom-Smith C, Rea TD, Sayre MR, Riedo FX, Russell D, Hiatt B, Montgomery P, Rao AK, Chow EJ, Tobolowsky F, Hughes MJ, Bardossy AC, Oakley LP, Jacobs JR, Stone ND, Reddy SC, Jernigan JA, Honein MA, Clark TA, Duchin JS; Public Health–Seattle and King County, EvergreenHealth, and CDC COVID-19 Investigation Team. Epidemiology of Covid-19 in a Long-Term Care Facility in King County, Washington. N Engl J Med. 2020 May 21;382(21):2005-2011. doi: 10.1056/NEJMoa2005412. Epub 2020 Mar 27. PMID: 32220208; PMCID: PMC7121761.
10. Mitjà O, Clotet B. Use of antiviral drugs to reduce COVID-19 transmission. Lancet Glob Health. 2020 May;8(5):e639-e640. doi: 10.1016/S2214-109X(20)30114-5. Epub 2020 Mar 19. PMID: 32199468; PMCID: PMC7104000.
11. Thanh Le T, Andreadakis Z, Kumar A, Gómez Román R, Tollefsen S, Saville M, Mayhew S. The COVID-19 vaccine development landscape. Nat Rev Drug Discov. 2020 May;19(5):305-306. doi: 10.1038/d41573-020-00073-5. PMID: 32273591.
12. Coronaviridae Study Group of the International Committee on Taxonomy of Viruses. The species Severe acute respiratory syndrome-related coronavirus: classifying 2019-nCoV and naming it SARS-CoV-2. Nat Microbiol. 2020;5(4):536‐544. doi:10.1038/s41564-020-0695-z
13. Andersen KG, Rambaut A, Lipkin WI, Holmes EC, Garry RF. The proximal origin of SARS-CoV-2. Nat Med. 2020;26(4):450‐452. doi:10.1038/s41591-020-0820-9
14. Cui J, Li F, Shi ZL. Origin and evolution of pathogenic coronaviruses. Nat Rev Microbiol. 2019;17(3):181‐192. doi:10.1038/s41579-018-0118-9
15. Wrapp D, Wang N, Corbett KS, Goldsmith JA, Hsieh CL, Abiona O, Graham BS, McLellan JS. Cryo-EM Wrapp D, Wang N, Corbett KS, Goldsmith JA, Hsieh CL, Abiona O, Graham BS, McLellan JS. Cryo-EM structure of the 2019-nCoV S-protein in the prefusion conformation. Science. 2020 Mar 13;367(6483):1260-1263. doi: 10.1126/science.abb2507. Epub 2020 Feb 19. PMID: 32075877; PMCID: PMC7164637.
16. Shang J, Ye G, Shi K, et al. Structural basis of receptor recognition by SARS-CoV-2. Nature. 2020;581(7807):221‐224. doi:10.1038/s41586-020-2179-y
17. Yan R, Zhang Y, Li Y, Xia L, Guo Y, Zhou Q. Structural basis for the recognition of SARS-CoV-2 by full-length human ACE2. Science. 2020;367(6485):1444‐1448. doi:10.1126/science.abb2762
18. Belouzard S, Chu VC, Whittaker GR. Activation of the SARS coronavirus S-protein protein via sequential proteolytic cleavage at two distinct sites. Proc Natl Acad Sci U S A. 2009;106(14):5871‐5876. doi:10.1073/pnas.0809524106
19. Walls AC, Park YJ, Tortorici MA, Wall A, McGuire AT, Veesler D. Structure, Function, and Antigenicity of the SARS-CoV-2 S-protein Glycoprotein. Cell. 2020 Apr 16;181(2):281-292.e6. doi: 10.1016/j.cell.2020.02.058. Epub 2020 Mar 9. PMID: 32155444; PMCID: PMC7102599.
20. Hoffmann M, Kleine-Weber H, Schroeder S, Krüger N, Herrler T, Erichsen S, Schiergens TS, Herrler G, Wu NH, Nitsche A, Müller MA, Drosten C, Pöhlmann S. SARS-CoV-2 Cell Entry Depends on ACE2 and TMPRSS2 and Is Blocked by a Clinically Proven Protease Inhibitor. Cell. 2020 Apr 16;181(2):271-280.e8. doi: 10.1016/j.cell.2020.02.052. Epub 2020 Mar 5. PMID: 32142651; PMCID: PMC7102627.
21. World Health Organization Draft landscape of COVID-19 candidate vaccines <https://www.who.int/who-documents-detail/draft-landscape-of-covid-19-candidate-vaccines>).
22. Pallesen J, Wang N, Corbett KS, Wrapp D, Kirchdoerfer RN, Turner HL, Cottrell CA, Becker MM, Wang L, Shi W, Kong WP, Andres EL, Kettenbach AN, Denison MR, Chappell JD, Graham BS, Ward AB, McLellan JS. Immunogenicity and structures of a rationally designed prefusion MERS-CoV S-protein antigen. Proc Natl Acad Sci U S A. 2017 Aug 29;114(35):E7348-E7357. doi: 10.1073/pnas.1707304114. Epub 2017 Aug 14. PMID: 28807998; PMCID: PMC5584442.
23. Henao-Restrepo AM, Camacho A, Longini IM, Watson CH, Edmunds WJ, Egger M, Carroll MW, Dean NE, Diatta I, Doumbia M, Draguez B, Duraffour S, Enwere G, Grais R, Gunther S, Gsell PS, Hossmann S, Watle SV, Kondé MK, Kéïta S, Kone S, Kuisma E, Levine MM, Mandal S, Mauget T, Norheim G, Riveros X, Soumah A, Trelle S, Vicari AS, Røttingen JA, Kieny MP. Efficacy and effectiveness of an rVSV-vectored vaccine in preventing Ebola virus disease: final results from the Guinea ring vaccination, open-label, cluster-randomised trial (Ebola Ça Suffit!). Lancet. 2017 Feb 4;389(10068):505-518. doi: 10.1016/S0140-6736(16)32621-6. Epub 2016 Dec 23. Erratum in: Lancet. 2017 Feb 4;389(10068):504. Erratum in: Lancet. 2017 Feb 4;389(10068):504. PMID: 28017403; PMCID: PMC5364328.
24. Padron-Regalado E. Vaccines for SARS-CoV-2: Lessons from Other Coronavirus Strains. Version 2. Infect Dis Ther. 2020 Apr 23;9(2):1-20. doi: 10.1007/s40121-020-00300-x. Epub ahead of print. PMID: 32328406; PMCID: PMC7177048.
25. van Doremalen N, Lambe T, Spencer A, Belij-Rammerstorfer S, Purushotham JN, Port JR, Avanzato V, Bushmaker T, Flaxman A, Ulaszewska M, Feldmann F, Allen ER, Sharpe H, Schulz J, Holbrook M, Okumura A, Meade-White K, Pérez-Pérez L, Bissett C, Gilbride C, Williamson BN, Rosenke R, Long D, Ishwarbhai A, Kailath R, Rose L, Morris S, Powers C, Lovaglio J, Hanley PW, Scott D, Saturday G, de Wit E, Gilbert SC, Munster VJ. ChAdOx1 nCoV-19 vaccination prevents SARS-CoV-2 pneumonia in rhesus macaques. bioRxiv [Preprint]. 2020 May 13:2020.05.13.093195. doi: 10.1101/2020.05.13.093195. PMID: 32511340; PMCID: PMC7241103.
26. Tatsis N, Ertl HC. Adenoviruses as vaccine vectors. Mol Ther. 2004 Oct;10(4):616-29. doi: 10.1016/j.ymthe.2004.07.013. PMID: 15451446; PMCID: PMC7106330.
27. Rauschhuber C, Noske N, Ehrhardt A. New insights into stability of recombinant adenovirus vector genomes in mammalian cells. Eur J Cell Biol. 2012 Jan;91(1):2-9. doi: 10.1016/j.ejcb.2011.01.006. Epub 2011 Mar 25. PMID: 21440326.
28. Sheets RL, Stein J, Bailer RT, Koup RA, Andrews C, Nason M, He B, Koo E, Trotter H, Duffy C, Manetz TS, Gomez P. Biodistribution and toxicological safety of adenovirus type 5 and type 35 vectored vaccines against human immunodeficiency virus-1 (HIV-1), Ebola, or Marburg are similar despite differing adenovirus serotype vector, manufacturer's construct, or gene inserts. J Immunotoxicol. 2008 Jul;5(3):315-35. doi: 10.1080/15376510802312464. PMID: 18830892; PMCID: PMC2777703.
29. Planty C, Chevalier G, Duclos MÈ, Chalmey C, Thirion-Delalande C, Sobry C, Steff AM, Destexhe E. Nonclinical safety assessment of repeated administration and biodistribution of ChAd3-EBO-Z Ebola candidate vaccine. J Appl Toxicol. 2020 Jun;40(6):748-762. doi: 10.1002/jat.3941. Epub 2020 Jan 21. PMID: 31965598.
30. Bassett JD, Swift SL, Bramson JL. Optimizing vaccine-induced CD8(+) T-cell immunity: focus on recombinant adenovirus vectors. Expert Rev Vaccines. 2011 Sep;10(9):1307-19. doi: 10.1586/erv.11.88. PMID: 21919620.
31. Tatsis N, Fitzgerald JC, Reyes-Sandoval A, Harris-McCoy KC, Hensley SE, Zhou D, Lin SW, Bian A, Xiang ZQ, Iparraguirre A, Lopez-Camacho C, Wherry EJ, Ertl HC. Adenoviral vectors persist in vivo and maintain activated CD8+ T cells: implications for their use as vaccines. Blood. 2007 Sep 15;110(6):1916-23. doi: 10.1182/blood-2007-02-062117. Epub 2007 May 17. PMID: 17510320; PMCID: PMC1976365.
32. Quinn KM, Zak DE, Costa A, Yamamoto A, Kastenmuller K, Hill BJ, Lynn GM, Darrah PA, Lindsay RW, Wang L, Cheng C, Nicosia A, Folgori A, Colloca S, Cortese R, Gostick E, Price DA, Gall JG, Roederer M, Aderem A, Seder RA. Antigen expression determines adenoviral vaccine potency independent of IFN and STING signaling. J Clin Invest. 2015 Mar 2;125(3):1129-46. doi: 10.1172/JCI78280. Epub 2015 Feb 2. PMID: 25642773; PMCID: PMC4362254.
33. Swadling L, Capone S, Antrobus RD, Brown A, Richardson R, Newell EW, Halliday J, Kelly C, Bowen D, Fergusson J, Kurioka A, Ammendola V, Del Sorbo M, Grazioli F, Esposito ML, Siani L, Traboni C, Hill A, Colloca S, Davis M, Nicosia A, Cortese R, Folgori A, Klenerman P, Barnes E. A human vaccine strategy based on chimpanzee adenoviral and MVA vectors that primes, boosts, and sustains functional HCV-specific T cell memory. Sci Transl Med. 2014 Nov 5;6(261):261ra153. doi: 10.1126/scitranslmed.3009185. PMID: 25378645; PMCID: PMC4669853.
34. Yang TC, Millar JB, Grinshtein N, Bassett J, Finn J, Bramson JL. T-cell immunity generated by recombinant adenovirus vaccines. Expert Rev Vaccines. 2007 Jun;6(3):347-56. doi: 10.1586/14760584.6.3.347. PMID: 17542750.
35. Aldhamen YA, Seregin SS, Amalfitano A. Immune recognition of gene transfer vectors: focus on adenovirus as a paradigm. Front Immunol. 2011 Sep 6;2:40. doi: 10.3389/fimmu.2011.00040. PMID: 22566830; PMCID: PMC3342374.
36. Rhee EG, Blattman JN, Kasturi SP, Kelley RP, Kaufman DR, Lynch DM, La Porte A, Simmons NL, Clark SL, Pulendran B, Greenberg PD, Barouch DH. Multiple innate immune pathways contribute to the immunogenicity of recombinant adenovirus vaccine vectors. J Virol. 2011 Jan;85(1):315-23. doi: 10.1128/JVI.01597-10. Epub 2010 Oct 20. PMID: 20962088; PMCID: PMC3014160.
37. Zhu FC, Li YH, Guan XH, Hou LH, Wang WJ, Li JX, Wu SP, Wang BS, Wang Z, Wang L, Jia SY, Jiang HD, Wang L, Jiang T, Hu Y, Gou JB, Xu SB, Xu JJ, Wang XW, Wang W, Chen W. Safety, tolerability, and immunogenicity of a recombinant adenovirus type-5 vectored COVID-19 vaccine: a dose-escalation, open-label, non-randomised, first-in-human trial. Lancet. 2020 Jun 13;395(10240):1845-1854. doi: 10.1016/S0140-6736(20)31208-3. Epub 2020 May 22. PMID: 32450106; PMCID: PMC7255193.
38. Abbink P, Lemckert AA, Ewald BA, Lynch DM, Denholtz M, Smits S, Holterman L, Damen I, Vogels R, Thorner AR, O'Brien KL, Carville A, Mansfield KG, Goudsmit J, Havenga MJ, Barouch DH. Comparative seroprevalence and immunogenicity of six rare serotype recombinant adenovirus vaccine vectors from subgroups B and D. J Virol. 2007 May;81(9):4654-63. doi: 10.1128/JVI.02696-06. Epub 2007 Feb 28. PMID: 17329340; PMCID: PMC1900173.
39. Colloca S, Barnes E, Folgori A, Ammendola V, Capone S, Cirillo A, Siani L, Naddeo M, Grazioli F, Esposito ML, Ambrosio M, Sparacino A, Bartiromo M, Meola A, Smith K, Kurioka A, O'Hara GA, Ewer KJ, Anagnostou N, Bliss C, Hill AV, Traboni C, Klenerman P, Cortese R, Nicosia A. Vaccine vectors derived from a large collection of simian adenoviruses induce potent cellular immunity across multiple species. Sci Transl Med. 2012 Jan 4;4(115):115ra2. doi: 10.1126/scitranslmed.3002925. PMID: 22218691; PMCID: PMC3627206.
40. Quinn KM, Da Costa A, Yamamoto A, Berry D, Lindsay RW, Darrah PA, Wang L, Cheng C, Kong WP, Gall JG, Nicosia A, Folgori A, Colloca S, Cortese R, Gostick E, Price DA, Gomez CE, Esteban M, Wyatt LS, Moss B, Morgan C, Roederer M, Bailer RT, Nabel GJ, Koup RA, Seder RA. Comparative analysis of the magnitude, quality, phenotype, and protective capacity of simian immunodeficiency virus gag-specific CD8+ T cells following human-, simian-, and chimpanzee-derived recombinant adenoviral vector immunization. J Immunol. 2013 Mar 15;190(6):2720-35. doi: 10.4049/jimmunol.1202861. Epub 2013 Feb 6. PMID: 23390298; PMCID: PMC3594325.
41. Vitelli A, Folgori A, Scarselli E, Colloca S, Capone S, Nicosia A. Chimpanzee adenoviral vectors as vaccines - challenges to move the technology into the fast lane. Expert Rev Vaccines. 2017 Dec;16(12):1241-1252. doi: 10.1080/14760584.2017.1394842. Epub 2017 Oct 30. PMID: 29047309.
42. Tapia MD, Sow SO, Lyke KE, Haidara FC, Diallo F, Doumbia M, Traore A, Coulibaly F, Kodio M, Onwuchekwa U, Sztein MB, Wahid R, Campbell JD, Kieny MP, Moorthy V, Imoukhuede EB, Rampling T, Roman F, De Ryck I, Bellamy AR, Dally L, Mbaya OT, Ploquin A, Zhou Y, Stanley DA, Bailer R, Koup RA, Roederer M, Ledgerwood J, Hill AVS, Ballou WR, Sullivan N, Graham B, Levine MM. Use of ChAd3-EBO-Z Ebola virus vaccine in Malian and US adults, and boosting of Malian adults with MVA-BN-Filo: a phase 1, single-blind, randomised trial, a phase 1b, open-label and double-blind, dose-escalation trial, and a nested, randomised, double-blind, placebo-controlled trial. Lancet Infect Dis. 2016 Jan;16(1):31-42. doi: 10.1016/S1473-3099(15)00362-X. Epub 2015 Nov 4. PMID: 26546548; PMCID: PMC4700389
43. Henao-Restrepo AM, Camacho A, Longini IM, Watson CH, Edmunds WJ, Egger M, Carroll MW, Dean NE, Diatta I, Doumbia M, Draguez B, Duraffour S, Enwere G, Grais R, Gunther S, Gsell PS, Hossmann S, Watle SV, Kondé MK, Kéïta S, Kone S, Kuisma E, Levine MM, Mandal S, Mauget T, Norheim G, Riveros X, Soumah A, Trelle S, Vicari AS, Røttingen JA, Kieny MP. Efficacy and effectiveness of an rVSV-vectored vaccine in preventing Ebola virus disease: final results from the Guinea ring vaccination, open-label, cluster-randomised trial (Ebola Ça Suffit!). Lancet. 2017 Feb 4;389(10068):505-518. doi: 10.1016/S0140-6736(16)32621-6. Epub 2016 Dec 23. Erratum in: Lancet. 2017 Feb 4;389(10068):504. Erratum in: Lancet. 2017 Feb 4;389(10068):504. PMID: 28017403; PMCID: PMC5364328.
44. Limbach K, Stefaniak M, Chen P, Patterson NB, Liao G, Weng S, Krepkiy S, Ekberg G, Torano H, Ettyreddy D, Gowda K, Sonawane S, Belmonte A, Abot E, Sedegah M, Hollingdale MR, Moormann A, Vulule J, Villasante E, Richie TL, Brough DE, Bruder JT. New gorilla adenovirus vaccine vectors induce potent immune responses and protection in a mouse malaria model. Malar J. 2017 Jul 3;16(1):263. doi: 10.1186/s12936-017-1911-z. PMID: 28673287; PMCID: PMC5496260.
45. Hassan AO, Dmitriev IP, Kashentseva EA, Zhao H, Brough DE, Fremont DH, Curiel DT, Diamond MS. A Gorilla Adenovirus-Based Vaccine against Zika Virus Induces Durable Immunity and Confers Protection in Pregnancy. Cell Rep. 2019 Sep 3;28(10):2634-2646.e4. doi: 10.1016/j.celrep.2019.08.005. PMID: 31484074; PMCID: PMC6750284.
46. Wrapp D, Wang N, Corbett KS, Goldsmith JA, Hsieh CL, Abiona O, Graham BS, McLellan JS. Cryo-EM structure of the 2019-nCoV S-protein in the prefusion conformation. Science. 2020 Mar 13;367(6483):1260-1263. doi: 10.1126/science.abb2507. Epub 2020 Feb 19. PMID: 32075877; PMCID: PMC7164637.
47. Aste-Amézaga M, Bett AJ, Wang F, Casimiro DR, Antonello JM, Patel DK, Dell EC, Franlin LL, Dougherty NM, Bennett PS, Perry HC, Davies ME, Shiver JW, Keller PM, Yeager MD. Quantitative adenovirus neutralization assays based on the secreted alkaline phosphatase reporter gene: application in epidemiologic studies and in the design of adenovector vaccines. Hum Gene Ther. 2004 Mar;15(3):293-304. doi: 10.1089/104303404322886147. PMID: 15018738.
48. Buchbinder SP, Mehrotra DV, Duerr A, Fitzgerald DW, Mogg R, Li D, Gilbert PB, Lama JR, Marmor M, Del Rio C, McElrath MJ, Casimiro DR, Gottesdiener KM, Chodakewitz JA, Corey L, Robertson MN; Step Study Protocol Team. Efficacy assessment of a cell-mediated immunity HIV-1 vaccine (the Step Study): a double-blind, randomised, placebo-controlled, test-of-concept trial. Lancet. 2008 Nov 29;372(9653):1881-1893. doi: 10.1016/S0140-6736(08)61591-3. Epub 2008 Nov 13. PMID: 19012954; PMCID: PMC2721012.
49. Zhu FC, Li YH, Guan XH, Hou LH, Wang WJ, Li JX, Wu SP, Wang BS, Wang Z, Wang L, Jia SY, Jiang HD, Wang L, Jiang T, Hu Y, Gou JB, Xu SB, Xu JJ, Wang XW, Wang W, Chen W. Safety, tolerability, and immunogenicity of a recombinant adenovirus type-5 vectored COVID-19 vaccine: a dose-escalation, open-label, non-randomised, first-in-human trial. Lancet. 2020 Jun 13;395(10240):1845-1854. doi: 10.1016/S0140-6736(20)31208-3. Epub 2020 May 22. PMID: 32450106; PMCID: PMC7255193.
50. Munster VJ, Feldmann F, Williamson BN, van Doremalen N, Pérez-Pérez L, Schulz J, Meade-White K, Okumura A, Callison J, Brumbaugh B, Avanzato VA, Rosenke R, Hanley PW, Saturday G, Scott D, Fischer ER, de Wit E. Respiratory disease in rhesus macaques inoculated with SARS-CoV-2. Nature. 2020 May 12. doi: 10.1038/s41586-020-2324-7. Epub ahead of print. PMID: 32396922.
51. Yu P, Qi F, Xu Y, Li F, Liu P, Liu J, Bao L, Deng W, Gao H, Xiang Z, Xiao C, Lv Q, Gong S, Liu J, Song Z, Qu Y, Xue J, Wei Q, Liu M, Wang G, Wang S, Yu H, Liu X, Huang B, Wang W, Zhao L, Wang H, Ye F, Zhou W, Zhen W, Han J, Wu G, Jin Q, Wang J, Tan W, Qin C. Age-related rhesus macaque models of COVID-19. Animal Model Exp Med. 2020 Mar 30;3(1):93-97. doi: 10.1002/ame2.12108. PMID: 32318665; PMCID: PMC7167234.
52. Chandrashekar A, Liu J, Martinot AJ, McMahan K, Mercado NB, Peter L, Tostanoski LH, Yu J, Maliga Z, Nekorchuk M, Busman-Sahay K, Terry M, Wrijil LM, Ducat S, Martinez DR, Atyeo C, Fischinger S, Burke JS, Slein MD, Pessaint L, Van Ry A, Greenhouse J, Taylor T, Blade K, Cook A, Finneyfrock B, Brown R, Teow E, Velasco J, Zahn R, Wegmann F, Abbink P, Bondzie EA, Dagotto G, Gebre MS, He X, Jacob-Dolan C, Kordana N, Li Z, Lifton MA, Mahrokhian SH, Maxfield LF, Nityanandam R, Nkolola JP, Schmidt AG, Miller AD, Baric RS, Alter G, Sorger PK, Estes JD, Andersen H, Lewis MG, Barouch DH. SARS-CoV-2 infection protects against rechallenge in rhesus macaques. Science. 2020 May 20:eabc4776. doi: 10.1126/science.abc4776. Epub ahead of print. PMID: 32434946; PMCID: PMC7243369.
